# How bursty infectiousness shapes epidemic dynamics

**DOI:** 10.64898/2026.07.15.26358199

**Authors:** Stephen M. Kissler

## Abstract

An epidemic’s expected course is determined by the magnitude and timing of a typical person’s infectiousness — captured, in turn, by the basic reproduction number and the generation-time distribution. These fundamental, population-average quantities can mask individual-level variation that shapes how an epidemic actually unfolds: for example, individual variation in the magnitude of infectiousness (overdispersion) creates superspreading, a key feature of the SARS-CoV-1 and SARS-CoV-2 epidemics. However, the impact of individual variation in infectiousness timing is less well understood. Here, we demonstrate that individual infectiousness timing varies substantially and to different degrees across pathogens. For some common pathogens, including influenza, measles, and SARS-CoV-2, infectiousness is “bursty”, or highly concentrated and variably-timed across individuals: for example, the window of appreciable infectiousness for SARS-CoV-2 may last for roughly a day, *vs*. the 9–12 days usually quoted. We show that bursty infectiousness creates superspreading without inherent superspreaders, makes epidemic timing more variable, amplifies the time-sensitivity of common interventions, and complicates inference of key epidemiological parameters. Together with the reproduction number, the generation-time distribution, and overdispersion, burstiness completes a family of basic parameters that govern how epidemics unfold.

---

Epidemic trajectories are profoundly shaped by random, individual-level effects. Super-spreading makes epidemics less likely to establish, but more explosive when they do; and chance events during an outbreak’s cryptic early stages can durably shift its overall timing ^1,2^. Accounting for such effects is vital for public health preparedness: while an epidemic’s expected course can be predicted using aggregate quantities, its stochastic behavior — including best- and worst-case scenarios, and the outcomes of targeted interventions — hinges upon individual variation in transmission capacity and timing.

This individual variation is anchored by two fundamental, population-level quantities: the basic reproduction number (*R*_0_), which captures how many people a typical infector will infect, and the generation interval (GI) distribution (*g*(*τ*)), which captures when those infections will occur (**Fig. 1A**) ^3^. Together, these two quantities are sufficient for describing an epidemic’s full expected path ^4^. The fact that individuals may vary in their transmission potential, relative to *R*_0_, has been recognized since the earliest contagion models ^5^; however, it was not until the formalization of the individual reproduction number, with the overdispersion parameter *k* to describe its variance, that the full impact of this variation was appreciated ^1^.

**Figure 1.**
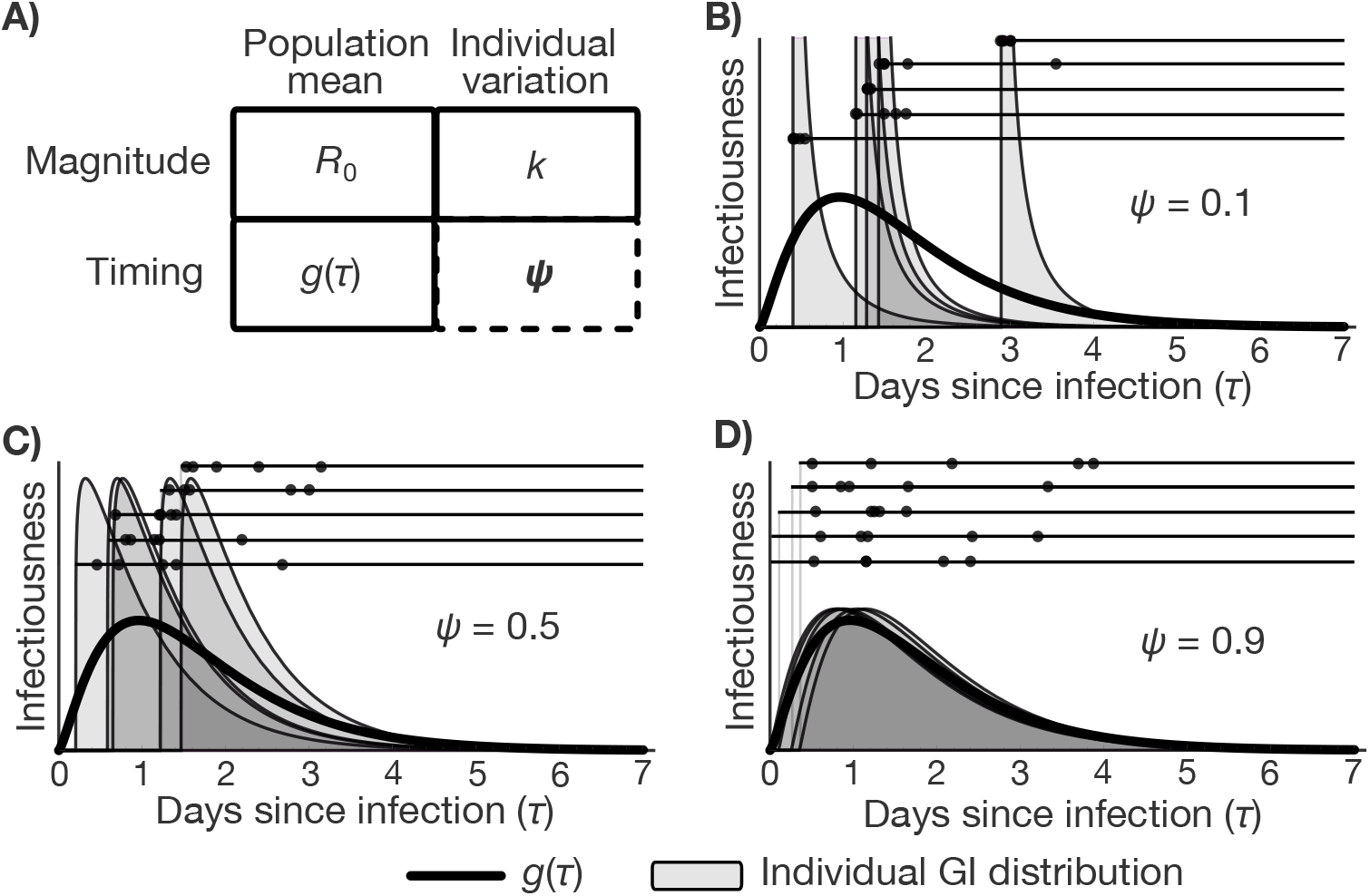
The individual GI distribution and the burstiness parameter *ψ*. (A) The burstiness parameter *ψ* completes a four-member family of parameters describing aggregate disease transmission properties (*R*_0_, *g*(*τ*)) and individual-level variation around those averages (*k, ψ*), together capturing the magnitude and timing of infectiousness. (B–D) A population-level generation interval distribution *g*(*τ*) (thick black curve) can arise from various individual GI distributions (shaded regions), with widths governed by *ψ*. Bursty infectiousness (panel B; *ψ* = 0.1) concentrates each infector’s transmissions near a single moment, with those moments distributed widely in time across infectors; intermediate *ψ* (panel C; *ψ* = 0.5) gives intermediate individual GI distribution spread; and more sustained infectiousness (panel D; *ψ* = 0.9) distributes transmissions from a single infector according to *g*(*τ*). The population average of the individual GI distributions yields the same *g*(*τ*) in each case. Points depict five simulated secondary infection times from five example infectors (corresponding to the illustrated individual GI distributions), demonstrating the tighter clustering of secondary infection times under bursty infectiousness.

The impact of the analogous variation in an individual’s timing and concentration of infectiousness, relative to *g*(*τ*), is less well understood. The reason for this is partly structural: existing disease transmission models can nominally capture heterogeneous infectiousness windows, but cannot precisely tune the time-course of individual infectiousness while holding *g*(*τ*) fixed. Nevertheless, evidence from influenza and SARS-CoV-2 suggests that the window during which a person is substantially infectious may span less than a day ^6^, even though the infectiousness periods for these viruses are typically quoted as lasting 6–8 days for influenza and 9–12 days for SARS-CoV-2^7,8^. These longer spans reflect the population-averaged *g*(*τ*), but *g*(*τ*)’s individual-level counterpart, the individual GI distribution ^9,10^, need not resemble it: *g*(*τ*) can be broad simply because different individuals’ infectiousness windows fall at different times, even if each person is only briefly infectious (**Fig. 1**). If individual GI distributions are in reality much narrower than *g*(*τ*) — *i*.*e*., “bursty” — this could impact everything from how individuals gauge their own transmission risk to how epidemics unfold overall.

Here, we systematically examine the impact of bursty infectiousness on epidemic dynamics. We introduce the burstiness parameter, *ψ* ∈ [0, 1], which governs the width of the individual GI distribution — specifically, the fraction of *g*(*τ*)’s variance attributable to within- *vs*. between-infector variation in transmission timing (**Fig. 1**; **Methods**). At the *ψ* = 0 extreme (maximum burstiness), each infector transmits at a single moment drawn from *g*(*τ*); at *ψ* = 1, their infections are distributed according to *g*(*τ*) itself. We assess the impact of *ψ* using a tractable, tunable model for the individual GI distribution and trace the consequences of bursty infectiousness across epidemic dynamics, superspreading, control, and inference. Critically, varying *ψ* does not affect *g*(*τ*) or *R*_0_; in this respect, *ψ* is the time-axis sibling of the overdispersion parameter *k*, providing a metric of individual-level variation that is hidden by population-level averages yet shapes epidemic dynamics ^1^.

## The Gamma burst model

To enable analytic study of *ψ*, we introduce the Gamma burst model (**Fig. 1B–D**; **Methods**). The model assumes the population-level *g*(*τ*) is Gamma-distributed with shape *α* and rate *β*. At the individual level, infectiousness timing follows a Gamma(*ψα, β*) distribution — the “burst” — following a latent period of length *l* ~ Gamma((1 − *ψ*)*α, β*). Because both components share the same rate *β*, their sum recovers Gamma(*α, β*) = *g*(*τ*) for any *ψ*. This decomposition mirrors within-host viral dynamics: stochastic viral replication early in infection creates variation in the onset of detectable virus (the latent period), while subsequent shedding kinetics are more consistent across individuals (the burst) ^11^. The qualitative consequences of *ψ* that we describe below are robust to model choice: we introduce and consider two additional burst models — an alternative Gamma model and a Log-normal model — in the **Methods**.

## Empirical estimates of *ψ*

Data from transmission clusters suggest that burstiness varies substantially across pathogens. We estimated *ψ* using published transmission trees with serial intervals for ten pathogens ^12^: measles, influenza, COVID-19, hepatitis A, pneumonic plague, Ebola, MERS, smallpox, Nipah virus, and norovirus. To translate from serial to generation intervals, we marginalized over each pathogen’s incubation period using literature values, then estimated *ψ* from a uniform prior (**Methods**). The resulting estimates spanned a wide range (**Fig. 2A–D**). Measles, influenza, and COVID-19 showed the strongest evidence of burstiness (95% credible interval for *ψ*: [0, 0.12], [0, 0.17], and [0, 0.12], respectively, translating into infectiousness bursts with standard deviations of [0, 1.3], [0, 0.9], and [0, 1.6] days). Nipah virus and norovirus showed more sustained infectiousness (*ψ*: [0.23, 0.99] and [0.43, 0.99]; burst standard deviations: [1.4, 3.0] and [1.3, 2.0] days).

**Figure 2.**
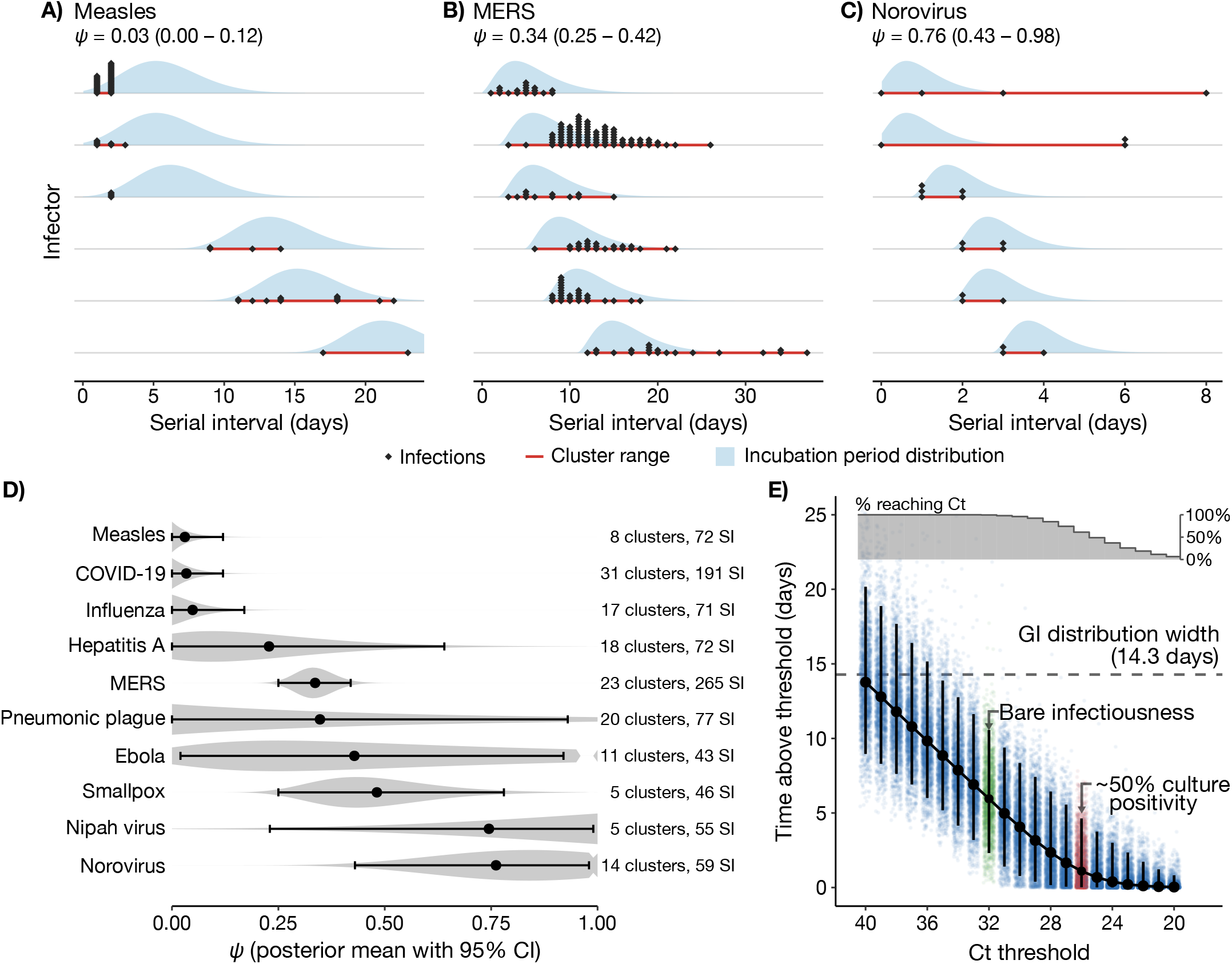
Empirical estimates of *ψ*. (A–C) Serial intervals (time between symptom onsets of secondary *vs*. index cases; points) from the six largest transmission clusters in the OutbreakTrees database ^12^ for measles (A), MERS (B), and norovirus (C). Each row represents one infector, ordered by earliest secondary onset; red bars span the within-cluster range of serial intervals. Blue shading shows the incubation period distribution ^32–34^, positioned so its lower tail aligns with each cluster’s earliest case (vertical scale arbitrary); it depicts the serial-interval spread expected if all of an infector’s secondary infections occurred at a single instant (*ψ* = 0), when onset timing reflects only each secondary case’s incubation period. Serial intervals confined to the shading and non-overlapping across clusters indicate low *ψ* (*e*.*g*., measles); intervals wider than it and overlapping across clusters indicate higher *ψ* (*e*.*g*., norovirus). Posterior means and 95% credible intervals for *ψ* are listed above each panel. (D) Posterior means (points) and 95% credible intervals (whiskers) for *ψ* across ten pathogens, estimated using the Gamma burst model (**Methods**); shaded regions show the posterior density. Estimates span near 0 (most bursty) to near 1 (most sustained). (E) Time spent with viral load above RT-qPCR cycle threshold (Ct) values across 1,991 SARS-CoV-2 infections (colored points); black points and whiskers show the population median and 5%–95% quantiles. Ct < 32 (green) marks potential infectiousness; Ct < 26 (red) marks approximately 50% culture positivity, suggesting moderate infectiousness. Increasingly fewer infections reach the lower Ct thresholds (higher viral load; grey step plot, top). At every threshold consistent with infectiousness, the time above threshold is substantially shorter than the SARS-CoV-2 generation-interval width (dashed line), consistent with bursty infectiousness.

These estimates should be regarded with some caution. Multiple biases can distort *ψ* in either direction: variable case-detection lags can disperse observed infection times, biasing *ψ* upward; while preferential reporting of superspreading events can bias *ψ* downward. Nevertheless, in simulated data matched to the empirical sample, we recovered *ψ* reliably for six of the ten pathogens (measles, COVID-19, influenza, MERS, smallpox, and norovirus), which includes the pathogens with the greatest evidence of burstiness; and the inferences were robust to imperfect ascertainment and incubation period misspecification (**Methods**; **Extended Data Fig. 2**). Furthermore, viral kinetics data from 1,991 SARS-CoV-2 infections indicate that the median amount of time spent above a viral load of 10^6.5^ genome copies per mL (cycle threshold [Ct] = 26) — a threshold roughly consistent with a 50% chance of viral culture positivity (**Methods**) — was just 1.1 days [5%–95% quantiles: 0.36–2.4 days], far shorter than the full GI distribution’s width ^13^ of 14.3 days (difference between 95% and 5% quantiles of *g*(*τ*)) (**Fig. 2E**). This, combined with independent evidence of narrow infectiousness windows for influenza and SARS-CoV-2^6^, supports the conclusion that *ψ* varies across pathogens and that bursty infectiousness is a genuine epidemiological phenomenon.

Moving forward, we illustrate the consequences of *ψ* using three benchmark pathogens spanning a range of reproduction numbers and generation interval shapes (**Fig. 1D**): influenza (*R*_0_ = 2, 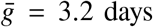, SD[*g*] = 2.1 days), SARS-CoV-2 omicron (*R*_0_ = 6, 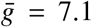, SD[*g*] = 4.6 days), and measles (*R*_0_ = 12, 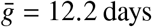, SD[*g*] = 3.6 days) ^13–18^, where 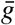 is the mean generation time and SD[*g*] is the standard deviation of the GI distribution — though the methods and findings are general.

## Coincidence superspreading

Superspreading happens when a person infects many more secondary cases than *R*_0_. It is typically assumed to arise from persistent individual-level traits: either a person has more than the average number of contacts, or sheds more than the typical amount of pathogen. The mechanism matters: contact-driven and infectiousness-driven superspreading produce qualitatively different epidemic dynamics and require different interventions ^19^.

A third type of superspreading can arise purely by chance, when an infectious burst happens to align with a brief period of high contacts. This “coincidence superspreading” is fundamentally different: it does not arise from any persistent individual trait and cannot be targeted by individual-level interventions. It thus represents a floor on overdispersion (the variability in individual transmission potential) ^1^ that persists even after contact-driven and infectiousness-driven heterogeneity has been accounted for. In this way, coincidence superspreading is the mechanistic link between overdispersion *k* and burstiness *ψ*.

Bursty infectiousness amplifies coincidence superspreading by capitalizing on brief fluctuations in contact rates. This can be illustrated using a simple periodic contact model, *c*(*t*) = 1 − *ζ* cos(2*πt*/*T*_*c*_), with amplitude *ζ* ∈ [0, 1] and period *T*_*c*_ > 0 relative to a baseline contact rate of 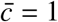 (**Fig. 3A–C, Methods**). This model captures contact variation over time but not across individuals: each person is identical except for when their infectiousness window falls, so any overdispersion that emerges reflects coincidence superspreading alone. Under the Gamma burst model, and assuming infections occur at uniform-random times relative to the contact process, the realized overdispersion (OD) in individual infectiousness can be expressed in terms of *k*:

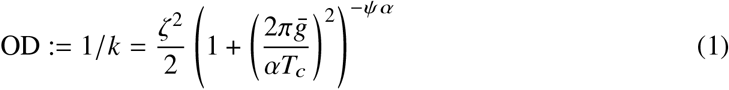

**Figure 3.**
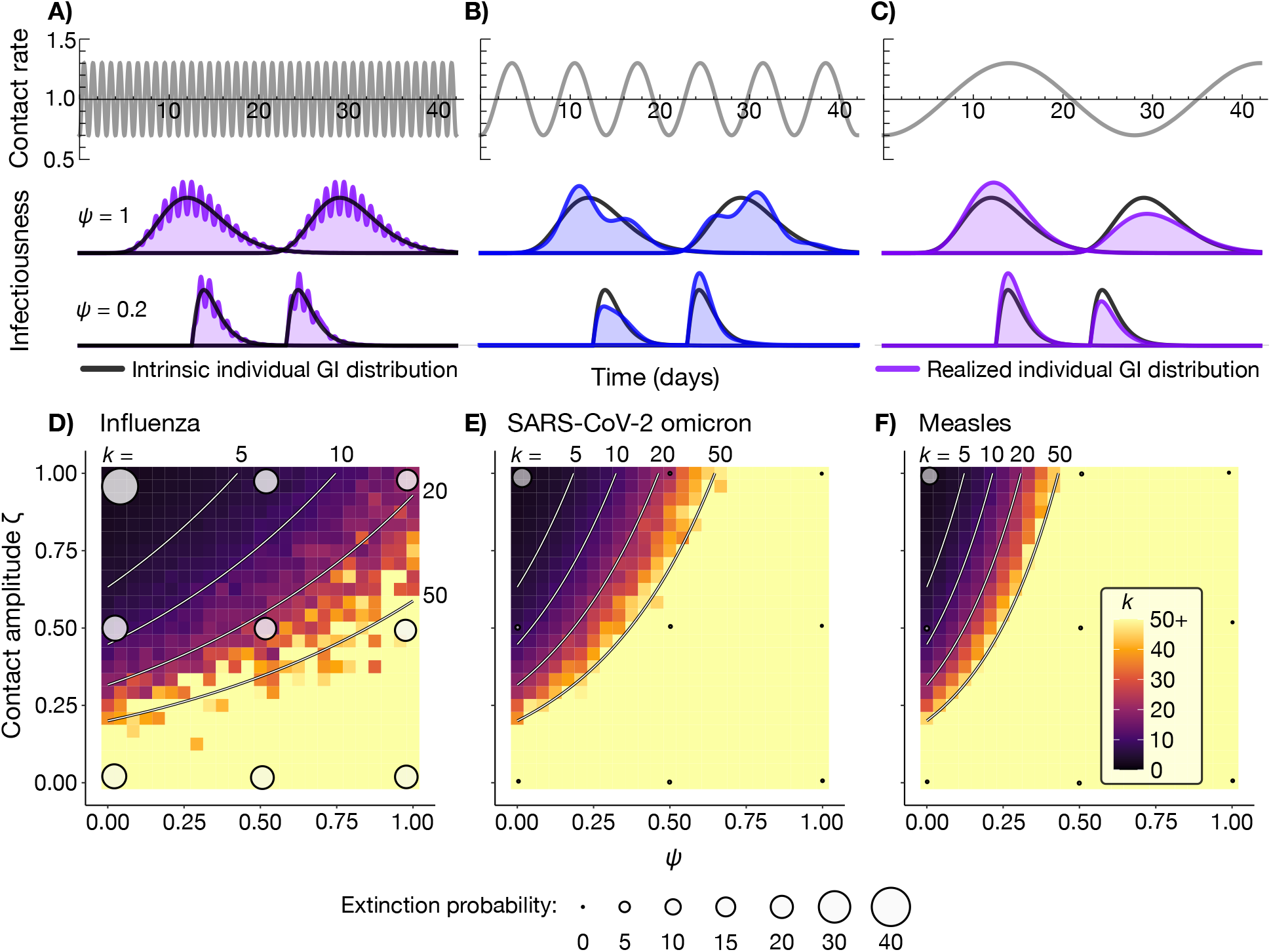
Coincidence superspreading and overdispersion. (A–C) Contact rates (top row) with intrinsic (black curves) and realized (blue shaded curves) individual GI distributions under sustained (*ψ* = 1, middle row) and bursty (*ψ* = 0.2, bottom row) infectiousness, with daily-(A), weekly-(B), and monthly-varying (C) contact patterns. GI distributions are based on measles parameters (*α* = 11.36, *β* = 0.931^17^). With rapid contact variation (A), both GI shapes average over the fluctuations, producing little overdispersion (difference in shaded areas). With weekly variation (B), sustained GI distributions still average over the variation, but bursty distributions are sensitive to timing. With monthly variation (C), the infectious period is short relative to the contact cycle, and both shapes are similarly sensitive to where infectiousness falls within the cycle. (D–F) Overdispersion parameter *k* as a function of *ψ* and contact amplitude *ζ* for influenza (D), SARS-CoV-2 omicron (E), and measles (F); lower *k* indicates greater overdispersion in secondary infection counts. Overdispersion is greatest when infectiousness is bursty (low *ψ*) and contacts are highly variable (high *ζ*). Circles show epidemic extinction probability at selected (*ψ, ζ*) combinations from simulations (**Methods**); circle area is proportional to extinction probability, which is highest where overdispersion is high.

Here, *α* is the shape parameter of *g*(*τ*) and 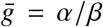 is the mean generation time. Small *k* corresponds to high overdispersion, while *k* → ∞ denotes a Poisson distribution of secondary cases (no overdispersion) ^1^.

Overdispersion is maximized by bursty infectiousness (*ψ* → 0): at the *ψ* = 0 extreme, the full variance of the contact process (*ζ* ^2^/2) is translated directly into overdispersion, while more sustained individual GI distributions (*ψ* → 1) average over the contact process. The sensitivity of OD to *ψ* depends on the ratio of the mean generation time 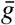 to the contact variation period *T*_*c*_, with the greatest impact when these two timescales are comparable 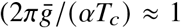; **Fig. 3B**). Faster contact variation gets averaged over by all but the narrowest individual GI distributions (**Fig. 3A**); slower variation produces overdispersion regardless of *ψ* (**Fig. 3C**; **Methods**). For the benchmark pathogens, where 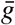 ranges from roughly 3–12 days, weekly contact variation falls within this sensitive zone. Empirical contact surveys confirm contacts vary on this timescale: the POLYMOD and BBC Pandemic studies ^20,21^ document 20%–40% more contacts on weekdays than weekends, corresponding to *ζ* ≈ 0.1–0.2 at *T*_*c*_ = 7 days.

Simulations confirm these predictions: for the benchmark pathogens, overdispersion is maximized by low *ψ* (high burstiness) and high *ζ* (large variation in contact levels), with substantial variation across *ψ*-values when *T*_*c*_ = 7 days (**Fig. 3D–F**). This has direct consequences for epidemic establishment, with bursty infectiousness translating into higher extinction probabilities. The results are robust to the choice of contact process: a stochastic contact-level-switching model yields the same qualitative link between burstiness, contact heterogeneity, and overdispersion (**Methods, Extended Data Figs. 3–4**). Results are also consistent across the alternative burst models (**Extended Data Fig. 4**).

## The epidemic latent period

Like infections, epidemics have a “latent period” — the span between the introduction of the first case and when the outbreak becomes established. Early-outbreak stochasticity shapes this latent period’s length, creating variability in epidemic timing even among identically parameterized outbreaks ^2^. Bursty infectiousness amplifies this variability, producing outbreaks that are more variable in timing and, on average, slower to establish.

To quantify this, we approximated early-epidemic exponential growth as a branching process, *Z* (*t*) ≈ *W e*^*rt*^, where *W* is a random initial condition capturing early-outbreak stochasticity — equivalently expressible as a time shift *ς*, so that *Z* (*t*) ≈ *e*^*r*(*t*+*ς*) 2^. An early-outbreak transmission burst that happens to fall at an extreme of *g*(*τ*) can have a major impact on the epidemic’s effective head start, encoded by variability in *W* and *ς*. Specifically, bursty infectiousness makes the effective initial condition more variable (inflates Var[*W*]), which makes epidemic timing more variable (increases Var[*ς*]) and shifts the typical epidemic slightly later (decreases *E* [*ς*]). The effect is strongest at high *R*_0_ (**Methods, Extended Data Fig. 5**).

Simulations confirm these predictions across the benchmark pathogens (**Fig. 4, Extended Data Fig. 5**). For SARS-CoV-2 omicron and measles, time-to-establishment depended strongly on *ψ*; for influenza, with its lower *R*_0_, differences were negligible. For measles, 96% of simulated outbreaks reached 5% prevalence by week 5 when infectiousness was sustained (*ψ* = 1), versus only 75% when maximally bursty (*ψ* = 0) — consistent with our prediction that the effect grows with *R*_0_. Findings were comparable across burst models (**Extended Data Fig. 5G**).

**Figure 4.**
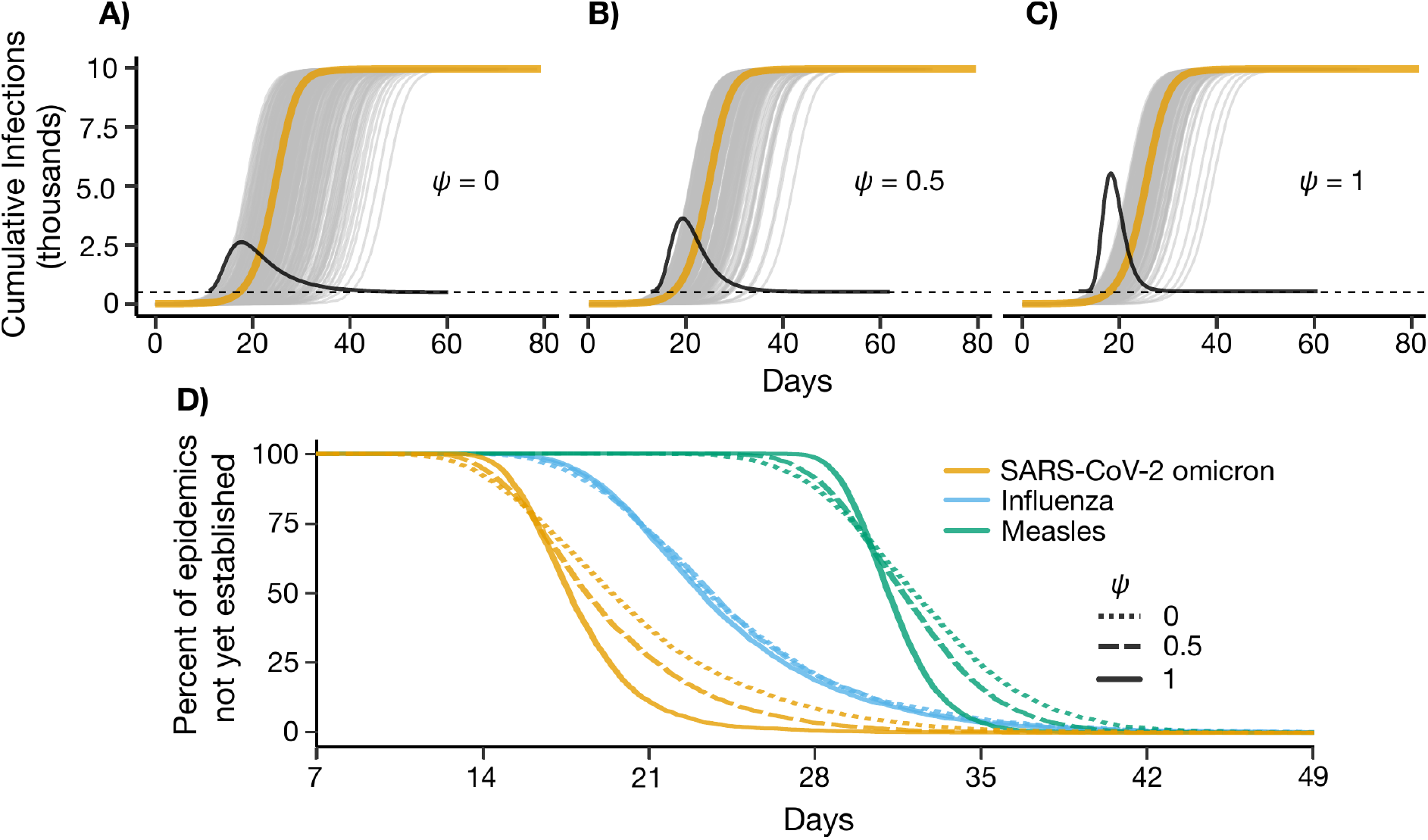
The epidemic latent period. (A–C) Cumulative infections over time for 250 simulated SARS-CoV-2 omicron epidemics in a population of 10,000 with one index case, for *ψ* = 0 (A), *ψ* = 0.5 (B), and *ψ* = 1 (C). Grey curves show individual simulation trajectories; orange curves show the deterministic mean-field trajectory. Black curves show the theoretical distribution of the time-shift parameter *ς*, which captures how early-outbreak stochasticity delays establishment (defined as 500 cumulative infections; dashed line). Under bursty infectiousness (low *ψ*), the grey curves are more widely dispersed, and establishment is more variable and later on average. (D) Survival curves for time to establishment across 5,000 simulations per pathogen-*ψ* combination (same setup as A–C), for the three benchmark pathogens (influenza: blue; SARS-CoV-2 omicron: orange; measles: green) and for *ψ* ∈ {0, 0.5, 1} (*ψ* = 0, dotted; *ψ* = 0.5, dashed; *ψ* = 1, solid). The effect of *ψ* on establishment timing grows with *R*_0_: curves are nearly identical across *ψ*-values for influenza (*R*_0_ = 2) but diverge substantially for SARS-CoV-2 omicron (*R*_0_ = 6) and measles (*R*_0_ = 12).

## Intervention sensitivity

Detect-and-isolate (D&I) interventions aim to identify and isolate infectious or pre-infectious individuals, limiting onward transmission. Their efficacy depends on the timing of detection relative to infectiousness ^22^. When detectability is linked to infectiousness timing — as is realistic for symptom- or test-based detection, where pathogen load determines both ^23^ — bursty infectiousness can cause D&I effectiveness to change dramatically with small shifts in detection time.

To quantify this, we calculated testing effectiveness (TE), defined as the expected fraction of transmission prevented by a D&I intervention ^24^, for symptom-based and test-based detection as a function of *ψ* (**Methods**). For both detection mechanisms, TE declined as detection shifted later relative to peak infectiousness, with the sharpest drops corresponding to the most bursty individual GI distributions (**Fig. 5, Extended Data Fig. 6A–F**). For SARS-CoV-2 omicron, shifting the typical symptom onset date from one day before to one day after peak infectiousness decreased TE from 97% to 2% when infectiousness was maximally bursty (*ψ* = 0), *vs*. only from 81% to 60% when infectiousness was sustained (*ψ* = 1). Similar results held for the other benchmark pathogens and for testing-based interventions, across all burst models (**Fig. 5, Extended Data Fig. 6**).

**Figure 5.**
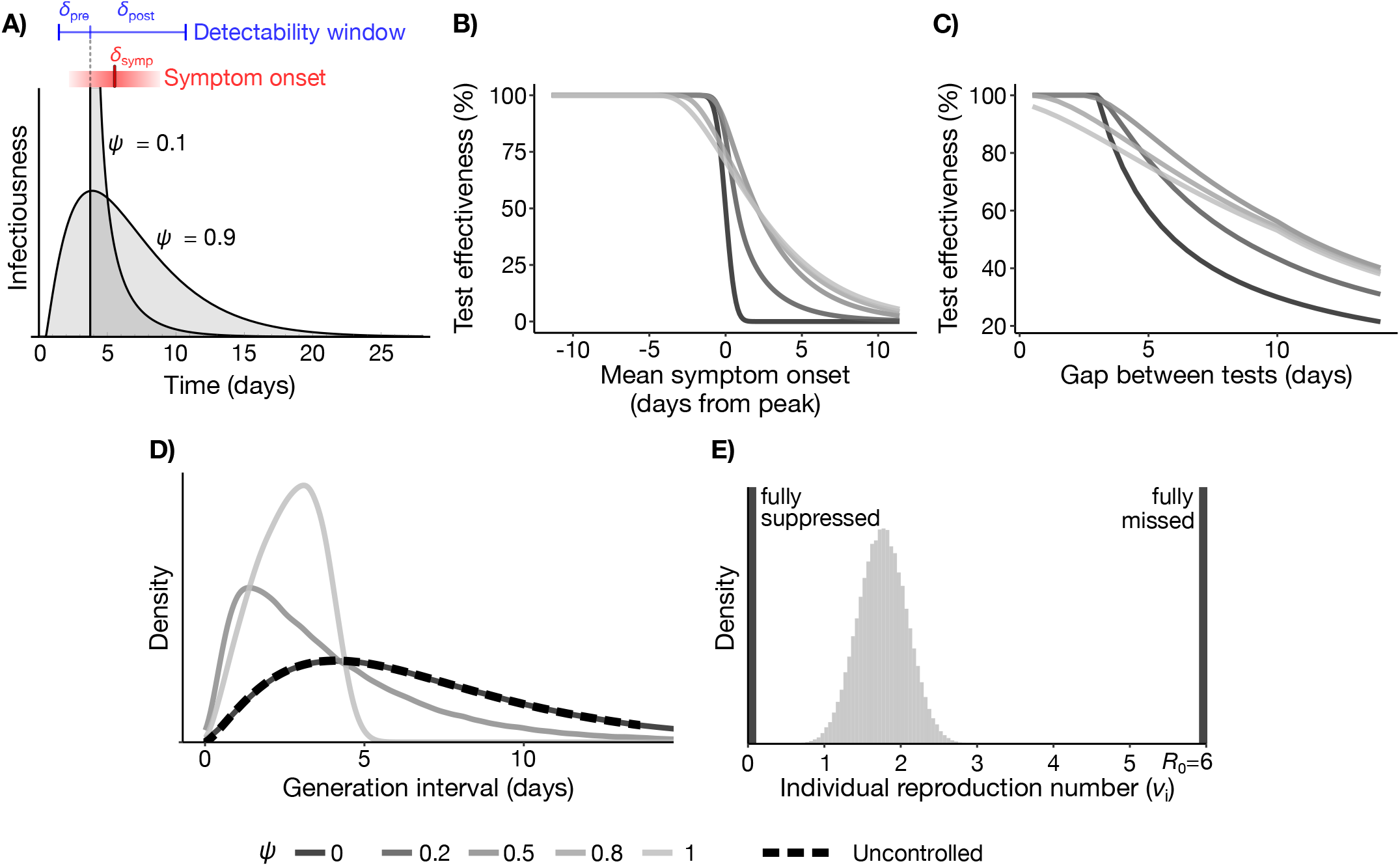
Detect-and-isolate interventions. (A) Illustration of symptom-based detection (red), where symptoms arise at a Normal(*δ*_symp_, *σ*_symp_)-distributed time after peak infectiousness; and screening-based detection (blue), where a detectability window spans *δ*_pre_–*δ*_post_ days before/after peak infectiousness and tests are administered at intervals Δ_test_. Both mechanisms have different impacts when infectiousness (grey) is bursty (*ψ* = 0.1) *vs*. sustained (*ψ* = 0.9): as *ψ* decreases, the infectiousness window is either fully suppressed or fully missed. (B, C) Test effectiveness for SARS-CoV-2 omicron across *ψ* ∈ {0, 0.2, 0.5, 0.8, 1} (dark = low *ψ*), as a function of (B) expected symptom onset *δ*_symp_ relative to peak (with *σ*_symp_ = 0.5) and (C) screening interval Δ_test_ (with [−*δ*_pre_, *δ*_post]_ = [−3, 7] days and an Exp(1)-distributed turnaround). Bursty infectiousness produces sharper TE declines. (D) Realized generation interval distributions under symptom-based D&I (*δ*_symp_ = 0, *σ*_symp_ = 0.5) for *ψ* = {0, 0.5, 1} (solid lines) *vs*. uncontrolled (dashed). For sustained infectiousness (*ψ* = 1), the population GI distribution is shortened; for bursty infectiousness (*ψ* = 0), it is unchanged. (E) Distribution of individual reproduction numbers (*ν*_*i*_) under the same symptom-based D&I intervention (*δ*_symp_ = 0, *σ*_symp_ = 0.5), for sustained (*ψ* = 1; light) and bursty (*ψ* = 0; dark) infectiousness. Bursty infectiousness inflates variation in *ν*_*i*_, since each infectiousness window is either fully suppressed (blocking all transmission) or fully missed (transmission proceeds unimpeded).

Beyond TE, D&I interventions produce two additional *ψ*-dependent effects. First, D&I preferentially removes late transmissions, truncating the effective generation interval distribution and shortening the mean generation time. This effect vanishes as *ψ* → 0, when detection becomes all-or-nothing and surviving transmissions proceed unimpeded (**Fig. 5D**). Consequently, D&I-controlled epidemics grow more quickly than TE alone predicts, especially when infectiousness is sustained: for SARS-CoV-2 omicron at *ψ* = 1, the estimated growth rate under D&I was 70% higher than the TE-only prediction (**Extended Data Fig. 6G–I**). Second, D&I interventions introduce overdispersion in secondary case counts when potential infectors are isolated at different points along their infectiousness period. Bursty infectiousness can translate slight detection-time variation into substantial overdispersion (**Fig. 5E**). This can increase the epidemic extinction probability beyond what the TE reduction alone would predict ^1^.

Unlike D&I interventions, gathering size restrictions reduce expected transmission by a factor that is independent of *ψ* (**Supplementary Information**) — but like D&I, they introduce additional *ψ*-dependent effects on overdispersion. By capping the size of large gatherings, they curtail the high-contact events that bursty infectiousness disproportionately exploits. Thus, gathering size restrictions limit the effects of coincidence superspreading, narrowing the overdispersion gap between bursty and sustained individual GI distributions (**Supplementary Information, Extended Data Fig. 7**).

## Epidemiological parameter inference

Estimating *g*(*τ*) and the epidemic growth rate *r* are critical tasks for calculating *R*_0_ and predicting how an epidemic will unfold. Bursty infectiousness complicates both. First, bursty infectiousness inflates day-to-day variation in case counts, especially when *R*_0_ is high, making estimates of *r* less certain (**Methods**; **Extended Data Fig. 8**). For measles, maximum likelihood estimates of *r* from simulated outbreaks ranged from 0.18–0.29/day when *ψ* = 1, *vs*. 0.07–0.52/day when *ψ* = 0 — a roughly 4x greater width, with the latter interval spanning anywhere from half to twice the actual growth rate of 0.228/day (**Extended Data Fig. 8D–F**). This phenomenon occurs across burst models (**Extended Data Fig. 8G–I**).

Second, bursty infectiousness reduces the information that “sibling” infections from a common index case provide about *g*(*τ*). When *ψ* = 1, each index case produces *R*_0_ offspring in expectation, each representing an independent draw from *g*(*τ*); when *ψ* = 0, the offspring infections occur simultaneously and represent just a single draw. Treating serial intervals from a single infector as independent draws from *g*(*τ*), as is common practice ^25^, can therefore produce overconfident estimates of *g*(*τ*)’s parameters (**Methods**; **Extended Data Fig. 9**).

## Discussion

The burstiness of infectiousness, as captured by the parameter *ψ*, shapes how epidemics unfold. Just as the overdispersion parameter *k* illustrates how not all pathogens with the same *R*_0_ behave alike, *ψ* reveals how not all pathogens with the same *g*(*τ*) behave alike. Burstiness impacts superspreading, outbreak timing, intervention effectiveness, and epidemiological parameter estimation — a broad enough range of consequences to justify treating it as a basic epidemiological property.

A central reason why bursty infectiousness has so far received little attention may be structural: standard models that depend only on *R*_0_ and *g*(*τ*) are *ψ*-invariant by construction. Methods like the linear chain trick narrow the individual GI distribution but simultaneously change *g*(*τ*), conflating two distinct effects ^26,27^. Defining *ψ* and introducing the Gamma burst model and its relatives decouples these effects, allowing the individual GI distribution’s width to vary freely while holding *g*(*τ*) fixed.

Bursty infectiousness explains coincidence superspreading — large transmission events that arise not from any persistent individual characteristic, but from the chance alignment of a narrow infectiousness window with a high-contact period. Coincidence superspreading formalizes the long-observed but previously *ad hoc* distinction between superspreading events and super-spreaders. It also carries an important practical implication: some fraction of overdispersion is structurally untargetable, setting a floor on overdispersion regardless of how well we identify and manage high-risk individuals.

While our conceptual model principally treats bursty infectiousness as a biological feature, bursty transmission patterns could also arise from, or be intensified by, bursty contact patterns ^28,29^, as our analysis of coincidence superspreading suggests. In this way, burstiness is also similar to infectiousness overdispersion, which can arise from a mixture of biological and contact-related effects ^1,19^. We could not disentangle the two mechanisms in our empirical estimates of *ψ*, given serial interval data alone, though the SARS-CoV-2 viral kinetics analysis suggests that burstiness is at least partly biological. Operationally, however, the two mechanisms should produce similar epidemiological consequences: both narrow the window of transmission relative to *g*(*τ*) and thus create clustering among secondary infection times from a single infector, which is the feature underpinning most of our findings. The detect-and-isolate result is the exception: its sharp dependence on detection timing relies on detection (*vs*. symptoms or testing) tracking biological infectiousness. If contact patterns drive burstiness, detection times (which are biologically linked) and transmission would decouple. Pathogens for which this is the case would show less steep D&I sensitivity gradients than our analysis predicts.

Our findings carry several practical implications. By amplifying variability in epidemic timing and day-to-day case counts, burstiness can increase uncertainty in outbreak forecasts; epidemic forecasting models should therefore include *ψ* as a structural parameter. For detect-and-isolate interventions, routine shifts in detection timing — such as changes in symptom onset timing relative to infectiousness ^30^, or adjustments to screening frequencies ^31^ — can produce sharp changes in effectiveness when infectiousness is bursty; accounting for burstiness is therefore critical when designing such interventions. When inferring *g*(*τ*), treating serial intervals from the same infector as independent draws can produce overconfident parameter estimates, so contact tracing records should, where possible, link secondary infections to index cases to enable transmission-cluster-level analyses. Finally, when building epidemic models that use *R*_0_ and *g*(*τ*) as inputs (*e*.*g*. in the renewal equation framework ^4^), pairing them with *ψ* and *k* can yield individual-level simulations that recover the mean-field dynamics while admitting realistic stochasticity.

Our findings are subject to several limitations. The burst models we consider are all unimodal, which may hold better for acute respiratory infections than for pathogens with more complex infectiousness dynamics, such as HIV, tuberculosis, or malaria. To isolate the impact of *ψ*, we decoupled the magnitude of infectiousness from its duration, but *ψ* and the individual reproduction number may in practice be correlated. Our contact models are stylized and assume a well-mixed population; they do not capture mobility, contact network structures, or household, community, or workplace stratification. These choices were made to highlight *ψ*’s impact clearly; results were robust across the model alternatives we explored.

Looking ahead, the most pressing open question is how best to measure burstiness. Clarifying the link between viral kinetics and infectiousness would make *ψ* more directly measurable, providing orthogonal evidence to contact tracing data. Pathogen phylogenies offer a further approach, since the near-simultaneous transmission from a single infector may leave characteristic phylogenetic signatures. Whether *ψ* is under evolutionary selective pressure, and under which circumstances pathogens may benefit from bursty *vs*. broad infectiousness, is a longer-horizon question with implications for both evolutionary biology and surveillance. The framework introduced here opens a path toward these goals.

Epidemic dynamics are governed by a small number of parameters: *R*_0_ and *g*(*τ*) define the expected trajectory, while *k* captures the individual variation in infectiousness that creates superspreading and explosive epidemic growth. The burstiness parameter *ψ* fills the remaining place. By attending to individual variation not just in the magnitude, but also in the timing of infectiousness, we stand to improve how we monitor, model, and respond to epidemics.

## Data Availability

All data produced are available online at https://github.com/skissler/BurstyInfectiousness

## Acknowledgements

The author thanks C. Middleton and L. Hadley for comments on the manuscript. The author thanks C. Fulton, D. Larremore, and T. Brown for input on conceptualization. Claude Code, version 2.1.150, was used to assist with code development, check mathematical derivations, and improve clarity of the writing.

## Author contributions

Conceptualization, methodology, investigation, writing, review & editing: SMK.

## Data availability

All data used in this study are publicly available and are collated, together with the analysis code, at https://github.com/skissler/BurstyInfectiousness/.

## Code availability

All code for the analyses and figures is available at https://github.com/skissler/BurstyInfectiousness/.

## Methods

### The burst models

Standard compartmental models, like the SEIR model, assume that individuals are infectious at a constant level for a variable duration (**Extended Data Fig. 1A**). This ties the magnitude of a person’s infectiousness to its duration, baking overdispersion into the model and making it impossible to study infectiousness timing separately from its magnitude. More flexible frameworks that tune the time-course of individual infectiousness (*e*.*g*., individual-based models or the linear chain trick) generally perturb the population-level *g*(*τ*) in the process, confounding the role of individual infectiousness timing with its population average.

To address these limitations, we adopt an approach where the fundamental epidemiological parameters — *g*(*τ*), *R*_0_, and the distribution of the individual reproduction number *ν*_*i*_ — are all fixed (we assume *ν*_*i*_ = *R*_0_ for all *i* unless otherwise specified, but this is not required). We seek to model the individual GI distribution in a way that allows us to tune the heterogeneity of infectiousness timing across individuals while leaving these other quantities unchanged. The variance of *g*(*τ*) must come from a combination of within-individual and between-individual variation in infectiousness timing; therefore, we introduce *ψ* ∈ [0, 1] as the proportion of *g*(*τ*)’s variance that arises from within-individual variation. Thus, *ψ* = 0 implies maximally narrow (bursty) individual GI distributions, with zero variance, where the bursts occur at times distributed according to *g*(*τ*) across individuals; and *ψ* = 1 implies maximally wide individual GI distributions, where each person’s individual GI distribution is identical to *g*(*τ*).

We introduce three tractable models of the individual GI distribution (“burst models”), parameterized by *ψ*: the (type-I) Gamma burst model featured in the main text, a type-II Gamma burst model, and a Log-normal burst model. Each describes the time of infection for offspring *j* from an infector *i* (indexed by *i*’s infection time) as *τ*_*i j*_ = *l*_*i*_ + *ϵ* _*j*_, where *l*_*i*_ is an infector-specific latent period and each *ϵ* _*j*_ is drawn independently from the “burst” distribution, with density *f*_*ϵ*_. To construct the models, we assume a parametric distributional form (*e*.*g*., Gamma or Log-normal) for both *g*(*τ*) and *f*_*ϵ*_, and we parameterize *f*_*ϵ*_ so its variance is Var[*f*_*ϵ*_] = *ψ* Var[*g*(*τ*)]. We then derive the distribution of the latent period, *f*_*l*_, needed to ensure the marginal distribution of *τ*_*i j*_ is *g*(*τ*): that is, *l*_*i*_ + *ϵ* _*j*_ ~ *g*(*τ*). Quantile-quantile plots illustrating agreement between the distribution of *l*_*i*_ + *ϵ* _*j*_ and *g*(*τ*) for all three burst models are provided in **Extended Data Fig. 1H–J**.

For the type-I Gamma burst model (**Figure 1B–D**), we assume *g*(*τ*) is a Gamma(*α, β*) density with shape *α* and rate 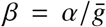, where 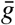 is the mean generation time. We assume the burst follows a Gamma(*ψα, β*) distribution, so that Var[*ϵ* _*j*_] = *ψα*/*β*^2^ = *ψ*Var[*g*(*τ*)] as required. By the summation property of Gamma random variables with equal rates, we can let *l*_*i*_ ~ Gamma((1 − *ψ*)*α, β*): this ensures *l*_*i*_ + *ϵ* _*j*_ ~ Gamma(*α, β*), as required.

For the type-II Gamma burst model (**Extended Data Fig. 1B–D**), we again assume *g*(*τ*) is a Gamma(*α, β*) density, but now we assume the burst follows a 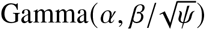 distribution: that is, the burst is a scaled version of *g*(*τ*), retaining the same shape. As before, 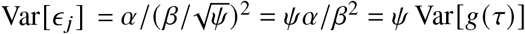. Unlike for the type-I Gamma burst model, the distribution of *l*_*i*_ = *τ*_*i j*_ − *ϵ* _*j*_ has no simple closed form; however, using the Lévy characterization of the Gamma density, we can construct an exact compound-Poisson sampler for *l*_*i*_ that ensures *l*_*i*_ + *ϵ* _*j*_ ~ Gamma(*α, β*) (**Supplementary Information**).

For the Log-normal burst model (**Extended Data Fig. 1E–G**), we assume *g*(*τ*) ~ Lognormal(*μ, σ*^2^). We assume the burst is a scaled version of *g*(*τ*), so that 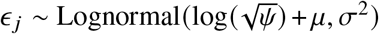, which ensures Var[*ϵ* _*j*_] = *ψ*Var[*g*(*τ*)]. We characterize the distribution of *l*_*i*_ as a Log-normal distribution with mean 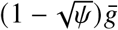 and variance (1 − *ψ*)Var[*g*(*τ*)]; this ensures that 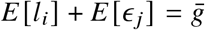 and Var[*l*_*i*_] + Var[*ϵ* _*j*_] = Var[*g*(*τ*)] (*i*.*e*., the first two moments agree). While *l*_*i*_ + *ϵ* _*j*_ is no longer precisely log-normal, it approximates a Lognormal(*μ, σ*^2^) distribution as required, by the Fenton-Wilkinson approximation ^35^.

These three models are suited to different purposes: the type-I Gamma burst model is the simplest and most analytically tractable form, which is why we feature it in our main analysis. The type-II Gamma burst model is less analytically tractable, but allows the individual GI distribution to have a more natural shape across the full range of *ψ* (starting at 0, peaking, and returning to 0; whereas when *ψ* is small, the type-I Gamma burst model places the maximum infectiousness at the moment the burst begins). The Log-normal burst model can accommodate pathogens with heavy-tailed GI distributions where the Gamma distribution is ill-suited, at the cost of the resulting *g*(*τ*) being only approximately log-normal distributed.

While we generally assume that *ψ* is a single value shared across the entire population, these models are more flexible: since *τ*_*j*_ = *l*_*i*_ +*ϵ* _*j*_ ~ *g*(*τ*) marginally for all *i, j*, we can draw a unique *ψ*_*i*_ for each person from any distribution on [0,1], and we will still recover *g*(*τ*) from the individual GI distributions in expectation.

To simulate transmission using these models, we (1) draw a latent period *l*_*i*_ for each infector, (2) draw the number of offspring *χ*_*i*_ ~ Poisson(*ν*_*i*_) for each infector (throughout, we assume *ν*_*i*_ = *R*_0_), and (3) draw the additional infection-time lags *ϵ* _*j*_ for each offspring *j* from the appropriate burst model. To simulate epidemics, we used a population size of *N* = 10,000; secondary infections were chosen uniformly at random from the population, and offspring infection times were stored in a priority queue to process them in sequential order. Epidemic simulations were run until all infections had been processed. Full details are provided in the **Supplementary Information**.

### Estimating ψ from contact tracing data

We obtained serial interval data from 315 clusters across 10 pathogens (951 total serial intervals) from the OutbreakTrees database ^12^. To estimate *ψ* from these serial intervals, we characterized the serial interval between an infector *i* and secondary case *j* as *s*_*i j*_ = *τ*_*i j*_ + *d* _*j*_ −*d*_*i*_, where *τ*_*i j*_ is the generation interval and *d*_*i*_ and *d* _*j*_ are the incubation periods for the index and secondary cases, respectively (all unobserved). We assumed *τ*_*i j*_ = *l*_*i*_ + *ϵ* _*j*_ followed the type-I Gamma burst model and that *d* ~ Gamma(*a*_obs_, *b*_obs_) — an incubation period distribution with known (literature-informed) parameters (**Extended Data Table 1**). Using these preliminaries, we derived the likelihood of *ψ* (**Supplementary Equation 17**). We evaluated the posterior of *ψ* on a fine, evenly-spaced grid on [0, 1], assuming a flat prior.

To assess the identifiability of *ψ*, we generated synthetic datasets matched to the size of the empirical dataset using known generation interval distributions, incubation period distributions, and *ψ*-values, and we used the same likelihood machinery to determine for which pathogen-*ψ* combinations the value of *ψ* was recoverable (**Extended Data Fig. 2A**). We simulated imperfect ascertainment in the same way, but with larger simulated datasets thinned to match the size of the empirical data (**Extended Data Fig. 2B**). We ran a third sensitivity analysis using simulated data, but with incubation period distributions at half and twice the width of the values we obtained from the literature and used in our main analysis (**Extended Data Fig. 2C**). Full details are given in the **Supplementary Information**.

### Estimating infectiousness windows from SARS-CoV-2 viral kinetics

Following our prior work ^36^, we fit a Bayesian hierarchical viral kinetics model to 18,496 SARS-CoV-2 RT-qPCR cycle threshold (Ct) values from 1,991 documented SARS-CoV-2 infections in players, staff, and affiliates of the National Basketball Association (NBA), occurring between March 11, 2020 and July 28, 2022. Samples were combined anterior-nares and oropharyngeal swabs processed with the Roche cobas SARS-CoV-2 assay (target 1). The model describes each infection’s trajectory on the Ct scale as a piecewise-linear (“tent”) curve defined by four quantities: the timing of peak viral concentration, the peak magnitude (the Ct drop below the limit of detection, LOD = 40), and the durations of the proliferation (pre-peak) and clearance (post-peak) phases. Ct is treated as proportional to log_10_ viral load. Because the fitted trajectory is piecewise linear, the time each infection spends above any viral load threshold has a simple closed form; we evaluated this for a sequence of thresholds in 1-Ct increments (Ct ∈ {40, 39, …, 20}, where Ct = 40 is the LOD and lower Ct corresponds to higher viral load) across posterior mean trajectories.

To correlate Ct values with potential infectiousness, we rely on a few distinct lines of evidence. Across culture studies, viable virus is rarely recovered below roughly 10^5^ genome equivalents/mL, corresponding to Ct ≈ 32 on our assay ^37–40^ (**Supplementary Information**). This threshold is consistent with the operational thresholds used by professional sports testing programs as a marker of non-infectiousness ^41,42^. Likewise, various viral culture studies indicate that 50% culture positivity occurs at roughly Ct ≈ 26^37–39^ (**Supplementary Information**). Because Ct is not directly comparable across assays, we treat these thresholds as approximate. To avoid dependence on any single cutoff, we report the time each infection spends above every integer Ct threshold from 40–20. The qualitative conclusion, that the interval of appreciable infectiousness is shorter than the width of the generation interval distribution, holds for any threshold more stringent than the limit of detection (Ct ≈ 40) (**Fig. 2E**).

### Deriving overdispersion under time-varying contact patterns

To assess the impact of coincidence superspreading, we first defined two time-varying contact models. F or the periodic contact model (**Fig. 3A–C**), contact rates vary according to *c* 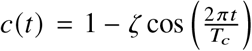 with amplitude *ζ* ∈ [0, 1] and period *T*_*c*_ > 0. Here, contact rates vary over time but not across individuals: everyone’s contact rates follow the same deterministic function. For the stochastic, Gamma-Poisson contact model (**Extended Data Fig. 3**), each person’s contact rate *c*_*i*_ (*t*) is a piecewise-constant process that switches to new levels at arrivals of a Poisson process with rate *λ*. The contact levels are drawn independently from a Gamma(*σ, σ*) distribution, which has mean 1 and variance 1/*σ*. Smaller *σ* yields more heterogeneous contacts. In contrast to the periodic model, each person’s contact trajectory is unique, but the population average is constant in time.

To derive an expression for the overdispersion constant *k* in terms of *ψ* with respect to these contact models, the critical link is the relation 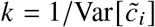, where 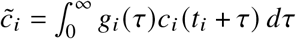. Here, *g*_*i*_(*τ*) is the individual GI distribution (defined by the burst model) and *c*_*i*_ is person *i*’s contact function; thus, 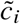 can be interpreted as the effective contact level for infector *i* over the course of their infection. This has the form of a classic problem in signal processing, where *c*_*i*_ is an input signal, *g*_*i*_ is an impulse response, and 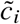 is the output signal evaluated at time *t*_*i*_; together, this comprises a linear system. The derivation of expressions for *k* in terms of *ψ* then follows standard theory ^43^ (see also the **Supplementary Information**). Methods for the related analysis of gathering-size restrictions (**Extended Data Fig. 7**) are given in the **Supplementary Information**.

### Calculating the epidemic latent period distribution

The cumulative number of infections during the early-epidemic exponential growth phase can be expressed as a branching process, *Z* (*t*). Let *r* be the epidemic’s exponential growth rate; then, as *t* → ∞, *e*^−*rt*^ *Z* (*t*) → *W* almost surely, where *W* is a random variable that effectively scales the process’ initial condition ^2^. We denote this convergence as *Z* (*t*) ≈ *W* · *e*^*rt*^. With a change of variables, *W* can be expressed as a random initial time shift: *Z* (*t*) ≈ *e*^*r* (*t*+*ς*)^ where *ς* = log(*W*)/*r*^2^.

An expression for Var[*W*] can be obtained through a direct derivation (**Supplementary Information**). Briefly, we begin from the distributional fixed point equation, which equates (in distribution) the branching process *Z* (*t*) to the sum of the branching processes initiated by its offspring: 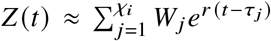, which yields 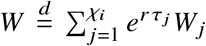; here *χ*_*i*_ ~ Poisson(*R*_0_) is the number of offspring produced by the index case *i, τ*_*j*_ are the infection times of those offspring, and *W* _*j*_ are *iid* copies of *W*. Taking the variance and isolating *W* yields an expression for Var[*W*] (**Supplementary Equation 79**); the key feature is that the *ψ*-dependence of Var[*W*] enters through a factor *ϕ*^(1−*ψ*) *α*^, where *ϕ* depends on *R*_0_ and *α*; it is increasing in *R*_0_ and is > 1 when *R*_0_ > 1, indicating that Var[*W*] is decreasing in *ψ* (*i*.*e*. increasing with burstiness) and that this *ψ*-dependence strengthens with *R*_0_.

To determine the implications for the time shift *ς*, we note that the distribution of *W* consists of a point mass at 0 (corresponding to epidemics that fail to establish) combined with a smooth density with positive support ^2^. The distribution of *W* conditional on survival (*W* |surv) is reasonably well-approximated by a Gamma distribution ^2^; thus, we assume *W* |surv ~ Gamma(*α*^∗^, *β*^∗^), and we derive expressions for *α*^∗^ and *β*^∗^ by matching the first two moments. Since *ς* = log(*W*)/*r*, and *W* is approximately Gamma-distributed, we can express the mean and variance of *ς* in terms of the digamma and trigamma functions using standard relations (**Supplementary Equations 87, 88**). These expressions confirm that *E* [*ς*] becomes more negative — *i*.*e*., the typical epidemic is delayed, establishing later — and that Var[*ς*] increases when infectiousness is bursty (*ψ* → 0).

### Calculating testing effectiveness

Testing effectiveness (TE) is the expected proportion by which a D&I intervention reduces population-level transmission ^24^. Formally, TE = *ρηP*(*τ* > *D*), where *ρ* ∈ [0, 1] is the adherence rate, *η* ∈ [0, 1] is the effectiveness of isolation (so that post-isolation transmission attempts succeed with probability 1 − *η*), *τ* is the time of a transmission attempt relative to the index case’s infection time, and *D* is the isolation time relative to infection onset.

We considered two detection mechanisms: symptom-based isolation, where symptoms arise at a Normal 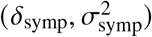-distributed offset from the peak infectiousness; and test-based screening, where diagnostic tests are administered at fixed intervals of length Δ_test_, and the first test that falls within a detectability window of total width *w* = *δ*_pre_+*δ*_post_, spanning *δ*_pre_ days before through *δ*_post_ days after peak infectiousness, triggers isolation. In both cases, *P*(*τ* > *D*) = *P*(*ϵ* _*j*_ > *m* _*ϵ*_ + *ι*), where *ϵ* _*j*_ is a draw from the burst distribution, *m* _*ϵ*_ is the mode of the burst distribution, and *ι* is the isolation time’s offset relative to *m* _*ϵ*_. Thus, we can express the TE for symptom-based isolation as

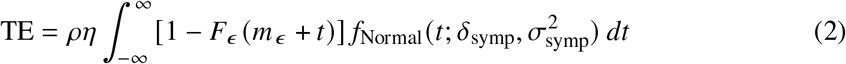

where *F*_*ϵ*_ is the CDF of the burst distribution and *f*_Normal_ is the Normal density. For testing-based isolation,

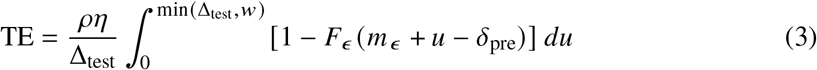

where *u* ~ Uniform(0, min(Δ_test_, *w*)) is the offset of the first test that falls in the detectability window. These expressions allow us to compute TE as a function of symptom onset time (*δ*_symp_) and testing interval (Δ_test_).

We simulated D&I interventions by drawing a random detection time *D*_*i*_ according to the detection mechanism and thinning all subsequent secondary infections with probability *η*. This allowed us to generate empirical distributions of the truncated generation interval distribution. Similarly, we computed the individual reproduction number *ν*_*i*_ by computing the CDF of person *i*’s burst distribution at time *D*_*i*_. Full details are in the **Supplementary Information**.

### Simulating epidemiological parameter inference

To assess the impact of bursty infectiousness on estimates of the epidemic growth rate *r*, we ran stochastic epidemic simulations in infinite populations (allowing all attempted secondary infections to succeed) to avoid susceptible depletion effects. For each of the benchmark pathogens and for *ψ* ∈ {0, 0.5, 1}, we simulated 5,000 epidemics seeded with a single index case, and stopped the simulations after cumulative infections exceeded a margin sufficient to cover 7 full days of growth past an epidemic establishment threshold of 100 cumulative infections. For each simulated epidemic, we estimated the empirical growth rate 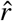 by fitting a Poisson generalized linear model with day as the predictor and daily case counts as the response, over a 7-day window beginning the day after cumulative infections first reached 100.

To assess the impact of bursty infectiousness on estimates of *g*(*τ*), we simulated 100 transmission clusters for each of the three benchmark pathogens and for *ψ* ∈ {0.1, 0.3, 0.5, 0.7, 0.9, 1.0}. Each index case *i* generated *χ*_*i*_ ~ Poisson(*R*_0_) offspring with infection times drawn from the Gamma burst model, and each infection (index cases and offspring) was assigned an independent Gamma(*a*_obs_ = 4, *b*_obs_ = 1)-day observation delay. The observed serial intervals *s*_*i j*_ = *τ*_*i j*_ +*d* _*j*_ −*d*_*i*_ were extracted from each cluster and pooled across clusters within a dataset. We then estimated the joint posterior for the shape and rate parameters (*α* and *β*) of *g*(*τ*) over a 100 × 100 grid centered on the true values under a flat prior, assuming each serial interval was an independent draw from the marginal serial interval density (*i*.*e*., not accounting for membership in transmission clusters). From each joint posterior we extracted marginal 95% credible intervals for *α* and *β* separately. We repeated this procedure across 500 simulated datasets per pathogen-*ψ* combination, and computed empirical coverage as the proportion of replicates in which the marginal 95% credible interval contained the true parameter value. Full details are in the **Supplementary Information**.

---

**Extended Data Fig. 1.**
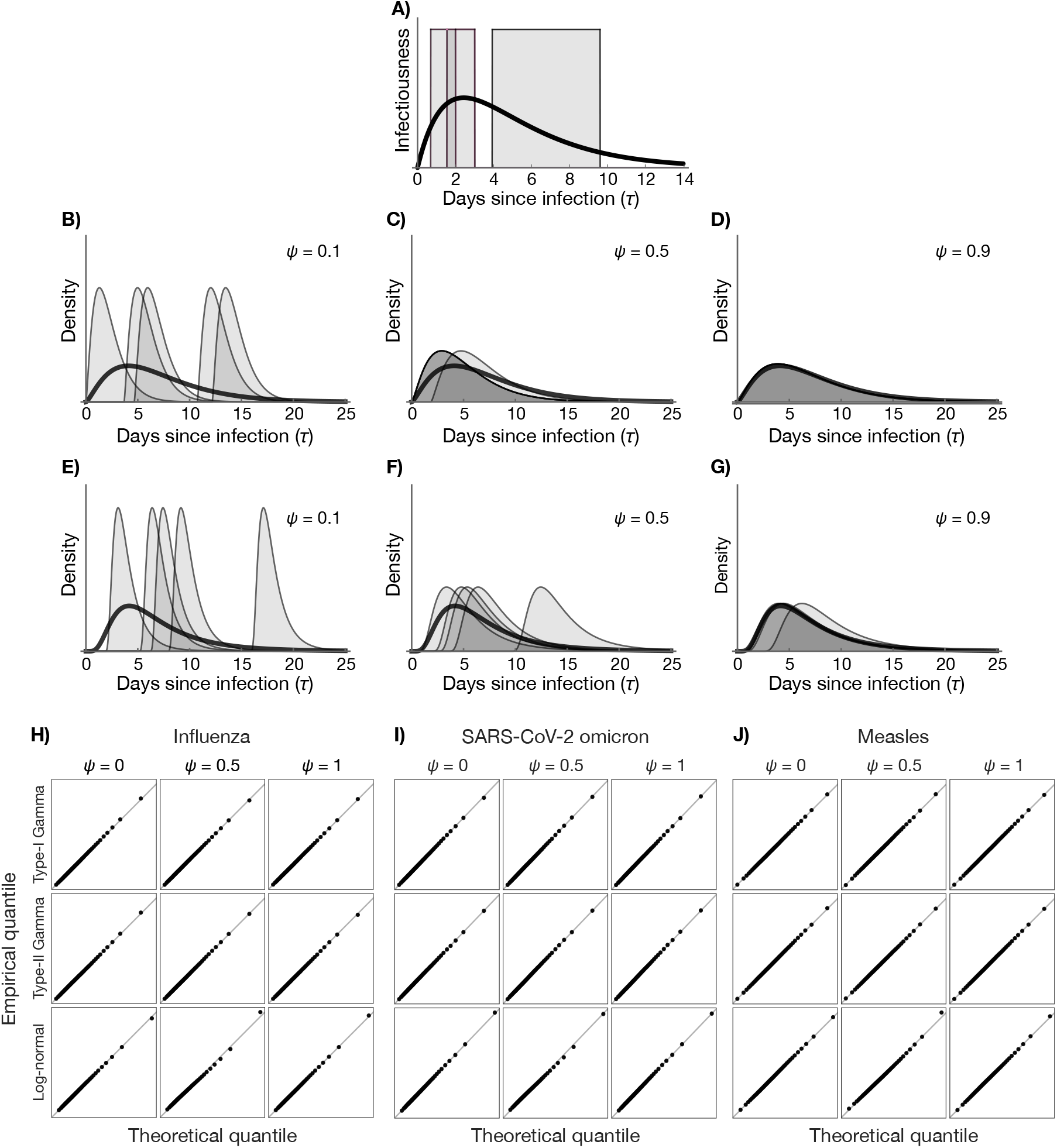
Alternative burst models. (A) For the standard compartmental SEIR model, individual-level infectiousness is assumed to have a constant level across individuals with variably-timed onsets and offsets; this is illustrated for three infectors (grey blocks). When these individual infectiousness profiles are summed across the population and normalized by the basic reproduction number *R*_0_, the generation interval distribution *g*(*τ*) is a smooth, unimodal curve (thick black curve). (B–G) Individual GI distributions (thin shaded curves) across *ψ*-values under the type-II Gamma (B–D) and Log-normal (E–G) burst models, for bursty (*ψ* = 0.1; B, E), intermediate (*ψ* = 0.5; C, F), and sustained (*ψ* = 0.9; D, G) infectiousness, using SARS-CoV-2 parameters. The individual-level curves average to the population-level generation interval distribution *g*(*τ*) (thick black curve), which is the same for B–D (a Gamma distribution) and for E–G (a Log-normal distribution). (H–J) Quantile–quantile plots comparing *n* = 50,000 simulated generation intervals against the target *g*(*τ*) distribution (points show 50 quantiles at evenly-spaced probability levels, the 0.5th–99.5th percentile, per panel) for each burst model (rows) and *ψ* ∈ {0, 0.5, 1} (columns), for influenza (H), SARS-CoV-2 omicron (I), and measles (J). The two Gamma models are exact for all *ψ*, while the Log-normal model is exact at *ψ* = 0 and *ψ* = 1, and approximate in between (slight upper-tail departures), since a sum of log-normals is only approximately log-normal.

**Extended Data Fig. 2.**
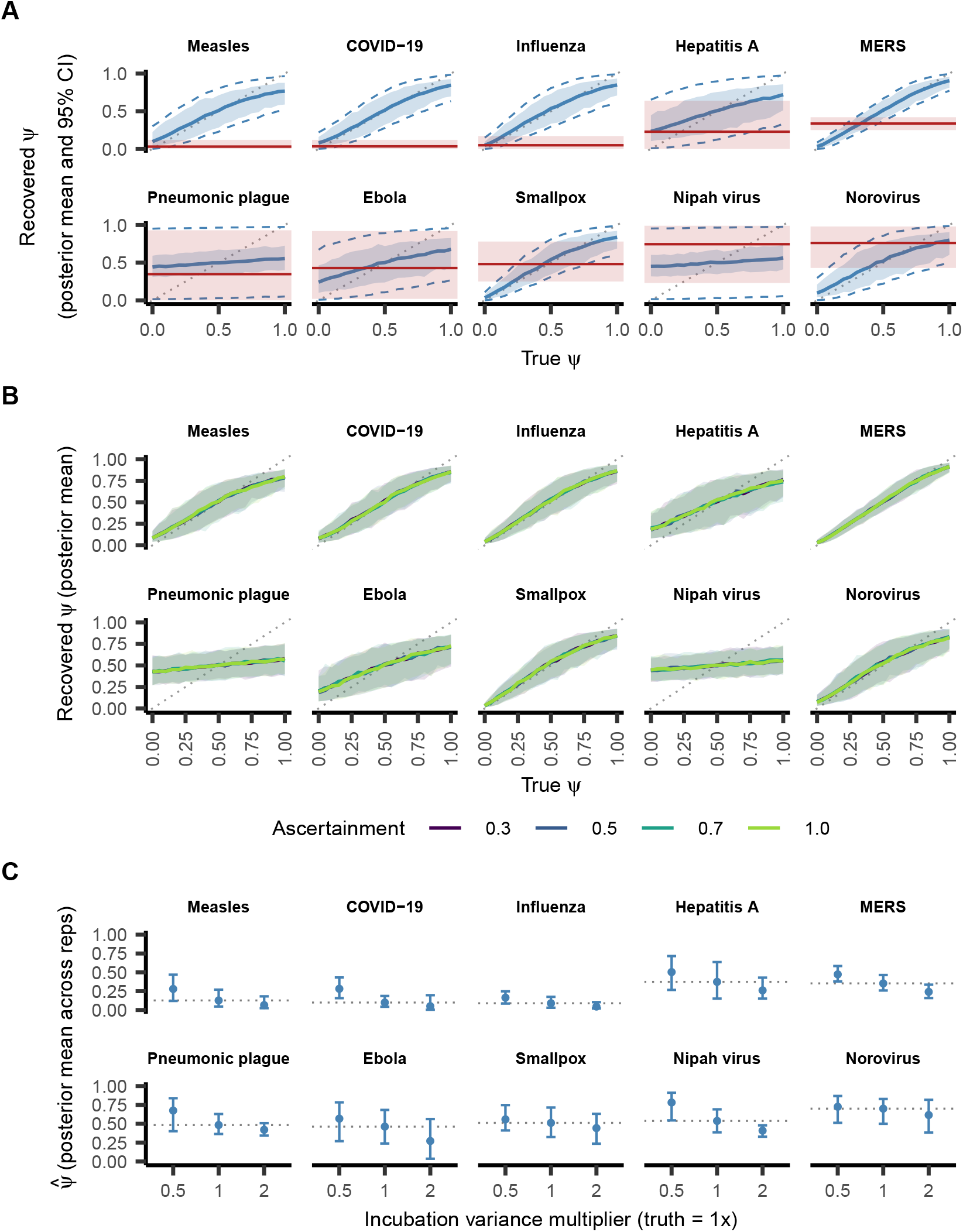
Identifiability of *ψ*. (A) Mean (blue curve) and 5%–95% range (blue band) of recovered posterior means across 500 synthetic datasets simulated at each *ψ*_true_ (horizontal axis), matched in size to the OutbreakTrees database ^12^ using known generation interval and incubation period distribution parameters (**Extended Data Table 1**). Blue dashed lines show the average 95% credible interval (CI) bounds across replicates; red bands and lines show the empirical 95% CI and posterior mean from the data (**Fig. 2D**). A narrow range of *ψ*_true_ over which the simulated (blue) and empirical (red) posteriors overlap indicates a well-localized estimate; a wide range indicates a poorly constrained one. (B) Mean (lines) and 5%–95% range (bands) of recovered posterior means across 200 synthetic datasets, *vs. ψ*_true_, for ascertainment probabilities *p*_asc_ ∈ {0.3, 0.5, 0.7, 1.0} (colors). The curves overlap in every panel, so *ψ* estimates are essentially insensitive to ascertainment. (C) Mean (points) and 5%–95% range (bars) of the recovered posterior mean across 100 synthetic datasets, generated with literature incubation period parameters but fit with the incubation period variance scaled by 0.5×, 1×, or 2× (horizontal axis); the dotted line marks the correctly specified (1×) estimate. Under-specifying the incubation period variance inflates *ψ*, as residual serial interval variance is attributed to within-infector variation in secondary infection timing. The bias magnitude differs across pathogens, but the *ψ* ordering is preserved across all three levels.

**Extended Data Fig. 3.**
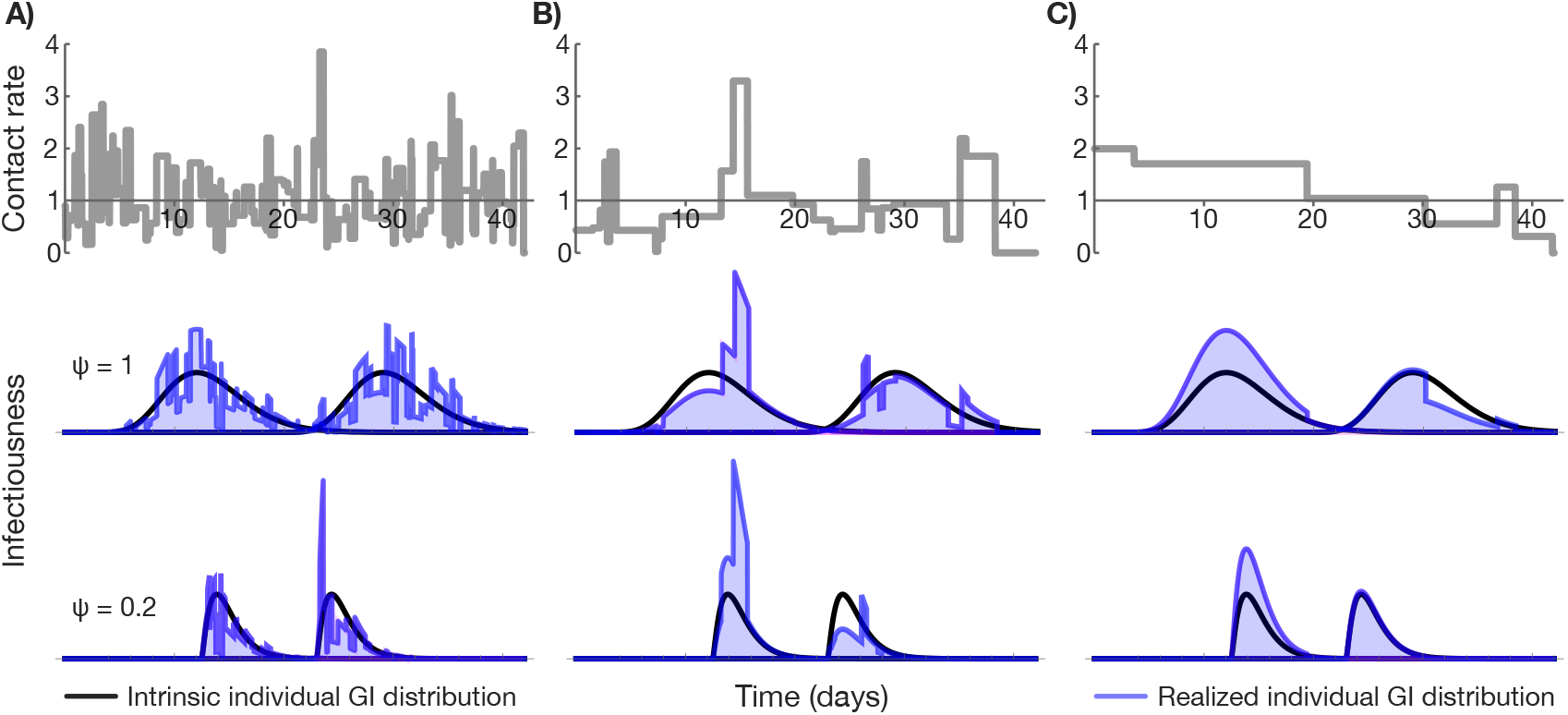
Realized individual generation interval distributions under stochastic (Gamma–Poisson) contacts. In the Gamma–Poisson contact model, each infector’s contact pattern follows a piecewise-constant process (top row) that jumps to a new Gamma (*σ, σ*)-distributed level at Poisson rate *λ*; here, *σ* = 2 throughout, and columns illustrate switching rates that are rapid (A; *λ* = 4, corresponding to 4 switches per day on average), moderate (B; *λ* = 4/7, corresponding to 4 switches per week on average), and slow (C; *λ* = 4/30, corresponding to 4 switches per month on average). The middle and bottom rows illustrate individual GI distributions under sustained (*ψ* = 1, middle) and bursty (*ψ* = 0.2, bottom) infectiousness: black curves depict the intrinsic individual GI distribution, and blue shaded curves depict the realized individual GI distribution, obtained by multiplying the intrinsic distribution by the contact process. Infectiousness is proportional to the blue shaded area. As with the periodic contact model, when the contact process varies rapidly (A), both bursty and sustained individual GI distributions effectively average over the contact process, producing little overdispersion (difference in the shaded areas within a *ψ*-level). When the switching rate is moderate (B), the sustained individual GI distribution (middle) still largely averages over the contact process, but the bursty individual GI distribution (bottom) is sensitive to shifts in the contact level, leading to overdispersion. When the switching rate is slow (C), both sustained and bursty individual GI distributions are sensitive to the contact process and yield overdispersion.

**Extended Data Fig. 4.**
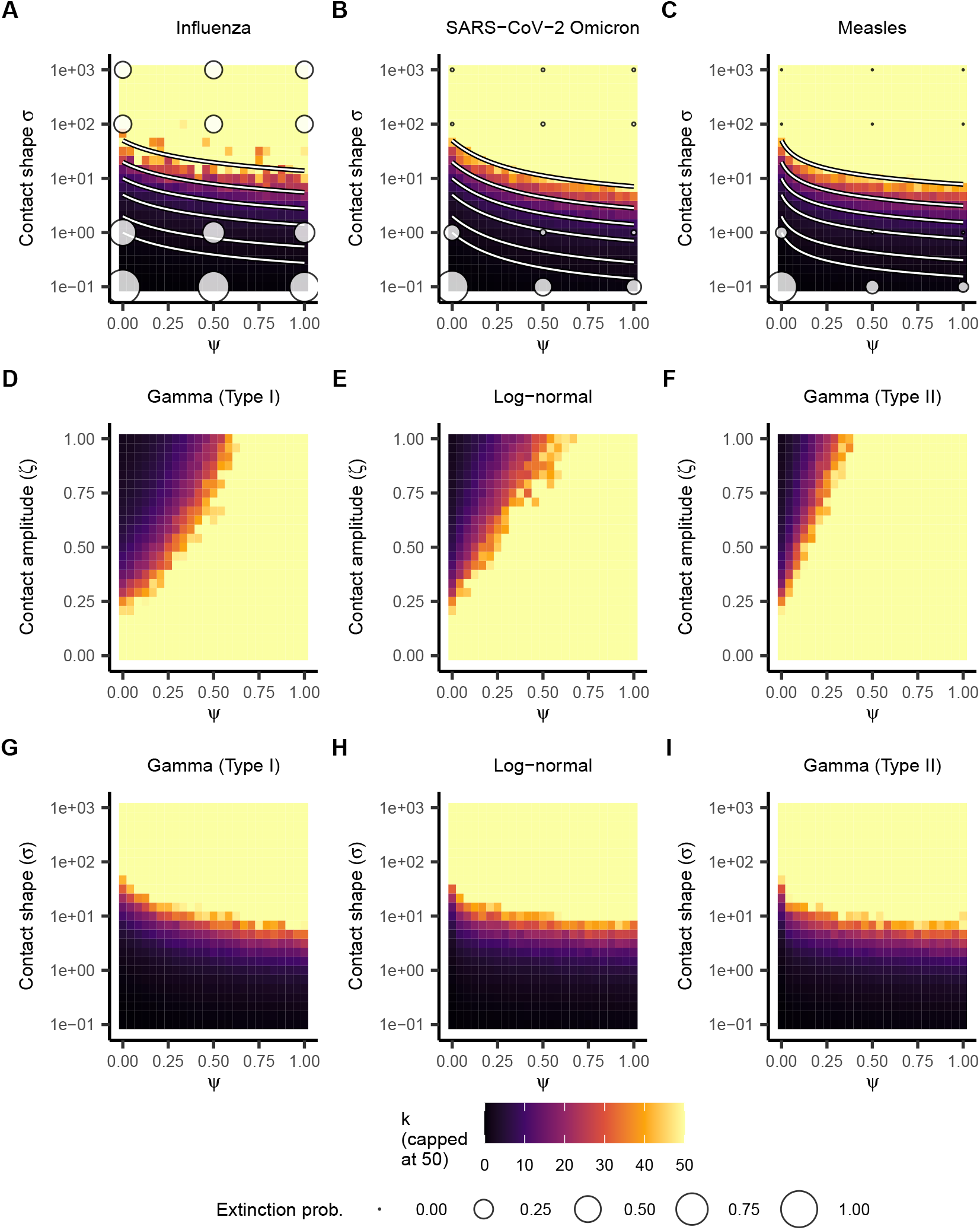
Overdispersion under stochastic (Gamma–Poisson) contacts. (A–C) Estimated value of the overdispersion parameter *k* (colored tiles) as a function of *ψ* and the contact level shape parameter *σ* for influenza (A), SARS-CoV-2 omicron (B), and measles (C), with switching rate *λ* = 1. Estimates of *k* were generated using 10,000 simulated clusters (index cases + offspring) per cell, using moment matching to the mean and variance of the secondary case counts. Overlaid lines depict level curves from the analytic derivation (**Supplementary Information, Eq. 69**). Lower *σ* (more dispersed contacts) yields more overdispersion (lower *k*). Discs represent epidemic extinction probabilities at select (*ψ, σ*) pairs; disc area is proportional to the epidemic extinction probability, estimated from 5,000 epidemic simulations per disc. (D–I) Estimated value of the overdispersion parameter *k* (colored tiles), using the same method as for A–C, as a function of *ψ* and contact amplitude/shape, for SARS-CoV-2, across contact models (periodic, D–F; Gamma-Poisson, G–I) and burst models (type-I Gamma: D, G; Log-normal: E, H; type-II Gamma: F, I). Overdispersion patterns are similar across burst models.

**Extended Data Fig. 5.**
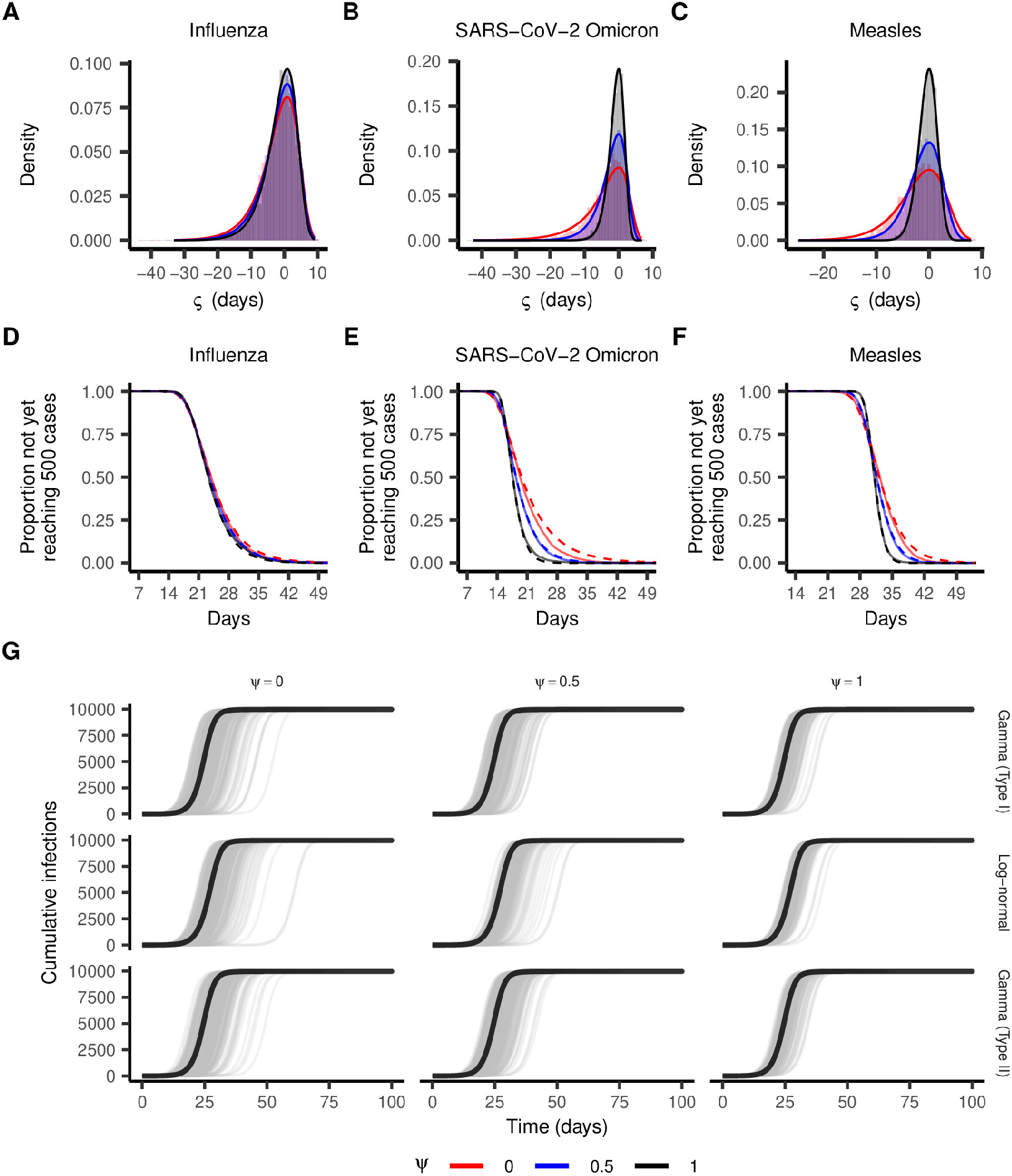
Burstiness and the epidemic latent period. (A–C) Empirical distributions (histograms) of the time shift *ς* from 5,000 simulated epidemics in a population of 10,000, seeded with a single index case, for influenza (A), SARS-CoV-2 omicron (B), and measles (C). Solid curves depict the theoretical distribution (**Supplementary Information, Eq. 84**) obtained by moment matching to the distribution of *W* (the effective initial condition). Bursty infectiousness (*ψ* → 0) shifts *E* [*ς*] downward (toward later epidemic onset) and increases Var [*ς*], especially for pathogens with high *R*_0_ (*e*.*g*., SARS-CoV-2, measles). (D–F) Survival curves depicting the proportion of epidemics that have not yet reached 5% cumulative prevalence (an indicator of epidemic establishment) as a function of time since the first case for influenza (D), SARS-CoV-2 omicron (E), and measles (F), for *ψ* = 0 (red), *ψ* = 0.5 (blue), and *ψ* = 1 (black). Solid curves depict the results from 5,000 simulated epidemics per pathogen-*ψ* combination in a population of size *N* = 10, 000 seeded with a single index case, while dashed curves depict the analytic predictions derived by shifting a deterministic epidemic according to the approximate time shift distribution (the distribution of *ς*). Divergence between the solid and dashed curves (most visible for SARS-CoV-2 omicron at *ψ* = 0) reflects the two-moment Gamma approximation to the distribution of *W*. (G) Simulated cumulative infection trajectories (grey) for SARS-CoV-2 omicron under alternative burst models with varying *ψ*, with deterministic mean-field trajectory (black), under the type-I Gamma burst model (top row), the Log-normal burst model (middle row), and the type-II Gamma burst model (bottom row), and for *ψ* = 0 (left-hand column), *ψ* = 0.5 (middle column), and *ψ* = 1 (right-hand column). Across burst models, bursty infectiousness (low *ψ*) amplifies variance in epidemic timing, reflected by greater spread of the stochastic trajectories.

**Extended Data Fig. 6.**
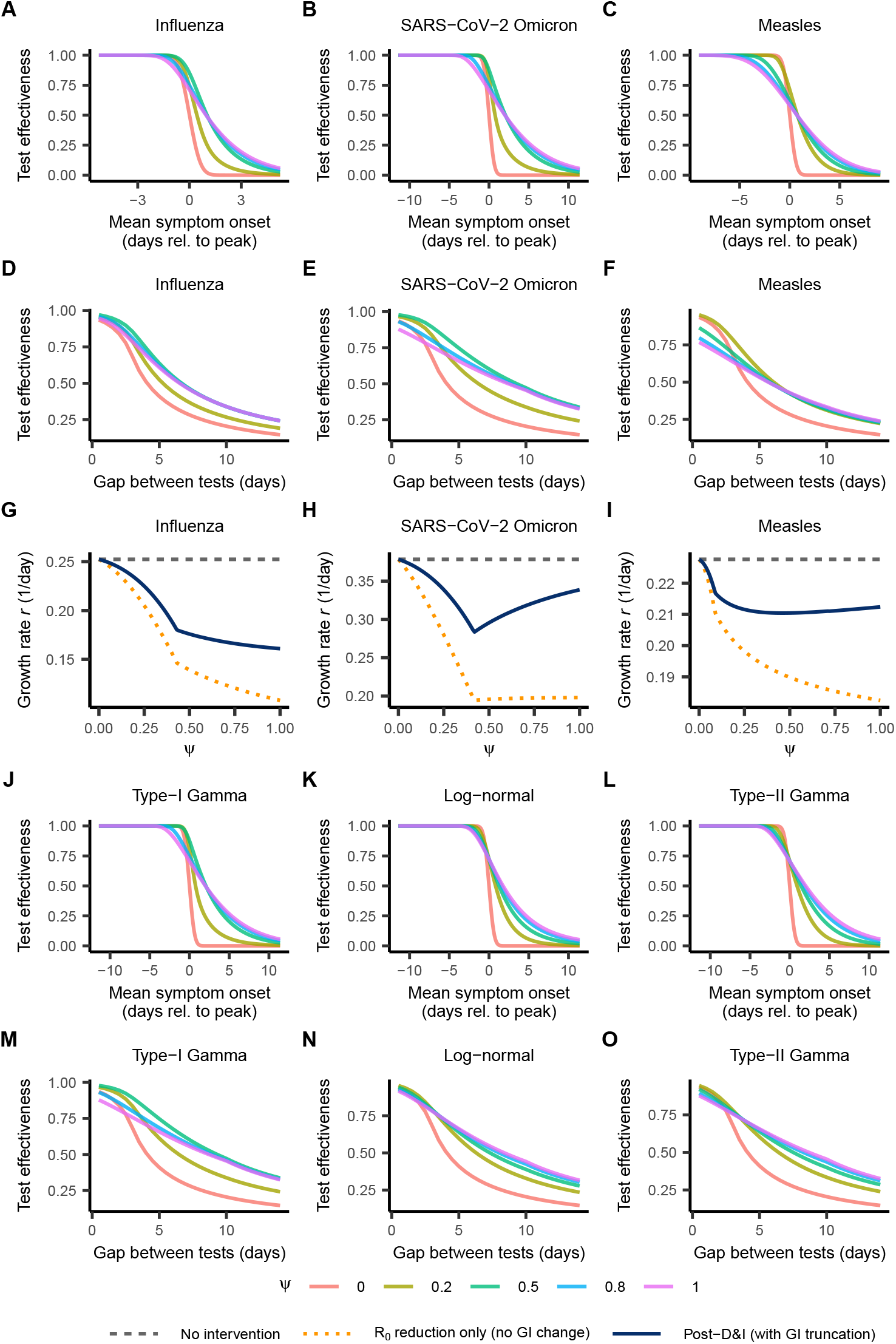
Detect-and-isolate interventions across pathogens. (A–F) Testing effectiveness (TE) for symptom-based isolation *vs*. mean symptom onset time (A–C) and for testing-based isolation *vs*. the gap between screening tests (D–F), for *ψ* ∈ {0, 0.2, 0.5, 0.8, 1} (colors), for influenza (A, D), SARS-CoV-2 omicron (B, E), and measles (C, F). Symptom onset occurs at a random Normal (*δ*_symp_, *σ*_symp_ = 0.5)-distributed time relative to peak infectiousness (the individual GI mode), with immediate isolation; for testing, the detection window spans 3 days before to 7 days after peak, with a Exp (1)-distributed isolation delay. Bursty infectiousness (*ψ* → 0) gives sharp drops in TE. (G–I) Epidemic growth rate *vs. ψ* under D&I intervention (solid navy) with deterministic, perfect isolation at *ι* = 2 days after peak, for influenza (G), SARS-CoV-2 omicron (H), and measles (I). Dashed grey depicts the growth rate under no intervention; dotted orange depicts the naive growth rate prediction if D&I only scaled *R*_0_ by 1 − TE without reshaping *g*(*τ*). D&I-controlled epidemics grow faster than the naive prediction, especially when infectiousness is sustained (*ψ* → 1), where generation interval truncation is greatest. (J–O) As in A–F, but for SARS-CoV-2 omicron across burst models: symptom-based (J–L) and testing-based (M–O) TE for the type-I Gamma (J, M), Log-normal (K, N), and type-II Gamma (L, O) models. The relationship of TE with *ψ* is similar across burst models.

**Extended Data Fig. 7.**
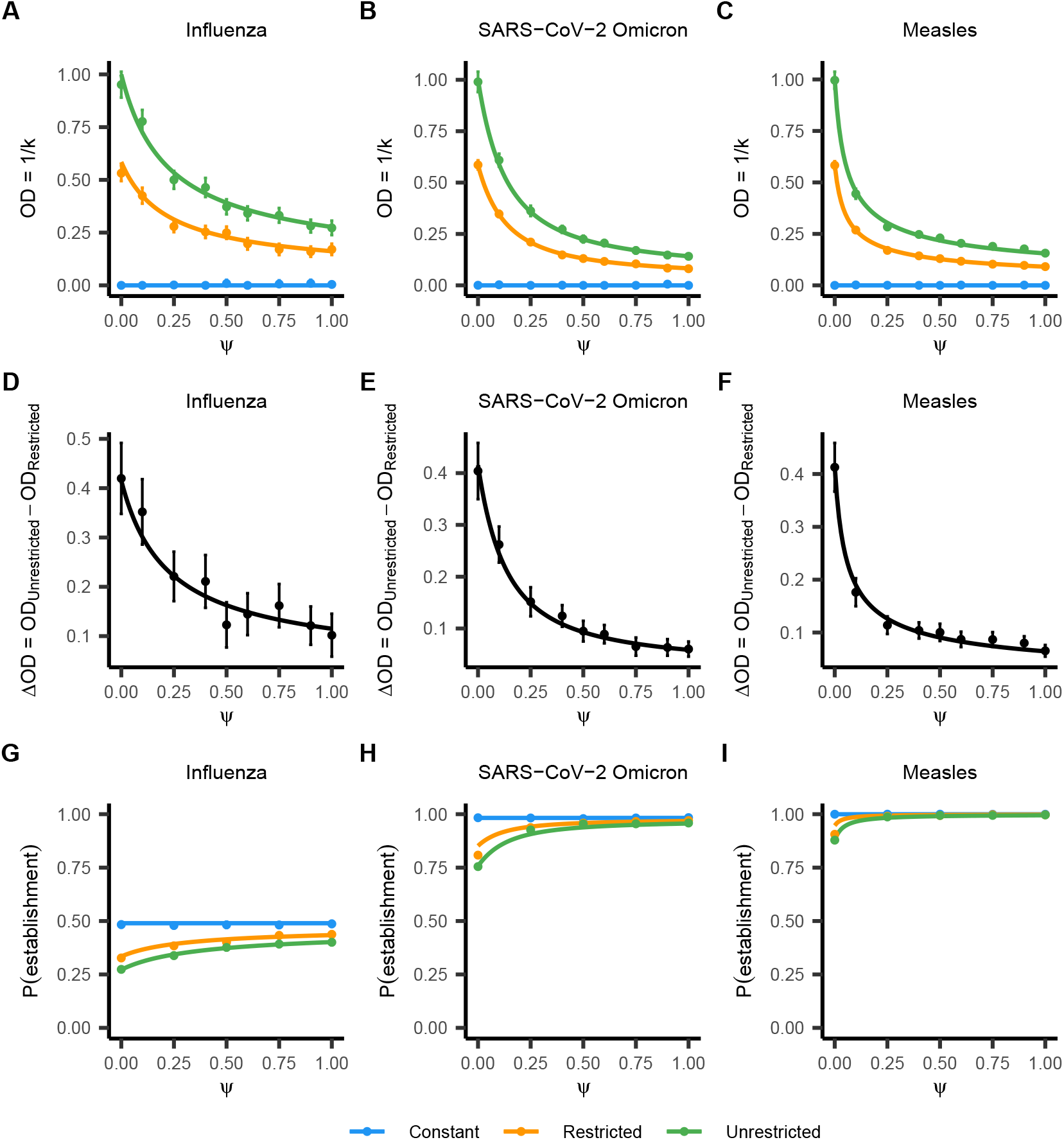
Gathering size restrictions. (A–C) Overdispersion (OD = 1/*k*) of secondary infection counts for influenza (A), SARS-CoV-2 omicron (B), and measles (C), under the Gamma-Poisson contact model (*σ* = 1, *λ* = 1); the “Restricted” scenario (orange) caps the contact process at *c*_max_ = 2 (twice the baseline contact rate), compared with “Unrestricted” (green; no cap) and “Constant” (blue; no contact variation) scenarios, both with effective reproduction number matched to Restricted. Lines depict the analytic prediction (**Supplementary Information**); points depict estimates from 10,000 simulated index cases per *ψ* and scenario (error bars: ±1.96 bootstrap standard error [SE] from 200 resamples). Restrictions narrow the overdispersion gap between bursty and sustained GI distributions (the “Restricted” curve decreases less sharply than “Unrestricted”). (D–F) Absolute difference in OD between the Restricted and Unrestricted scenarios, for influenza (D), SARS-CoV-2 omicron (E), and measles (F). Solid lines depict the analytic prediction (**Supplementary Information**); points depict the difference of the estimates shown in A–C (bars: ±1.96 bootstrap SE). The absolute OD reduction is largest with bursty infectiousness (*ψ* = 0). (G–I) Expected establishment probability *vs. ψ* from the Lloyd-Smith Negative Binomial extinction formula ^1^, across the three scenarios (colors as above), for influenza (G), SARS-CoV-2 omicron (H), and measles (I). Points depict the empirical establishment probabilities from 5,000 simulated epidemics per *ψ*–pathogen combination (bars: ±1.96 SE), with establishment defined as reaching 5% cumulative prevalence (500 cases in a population of *N* = 10, 000). The establishment boost from restrictions (Restricted *vs*. Unrestricted, matched *R*_eff_) concentrates at the bursty (*ψ* → 0) limit.

**Extended Data Fig. 8.**
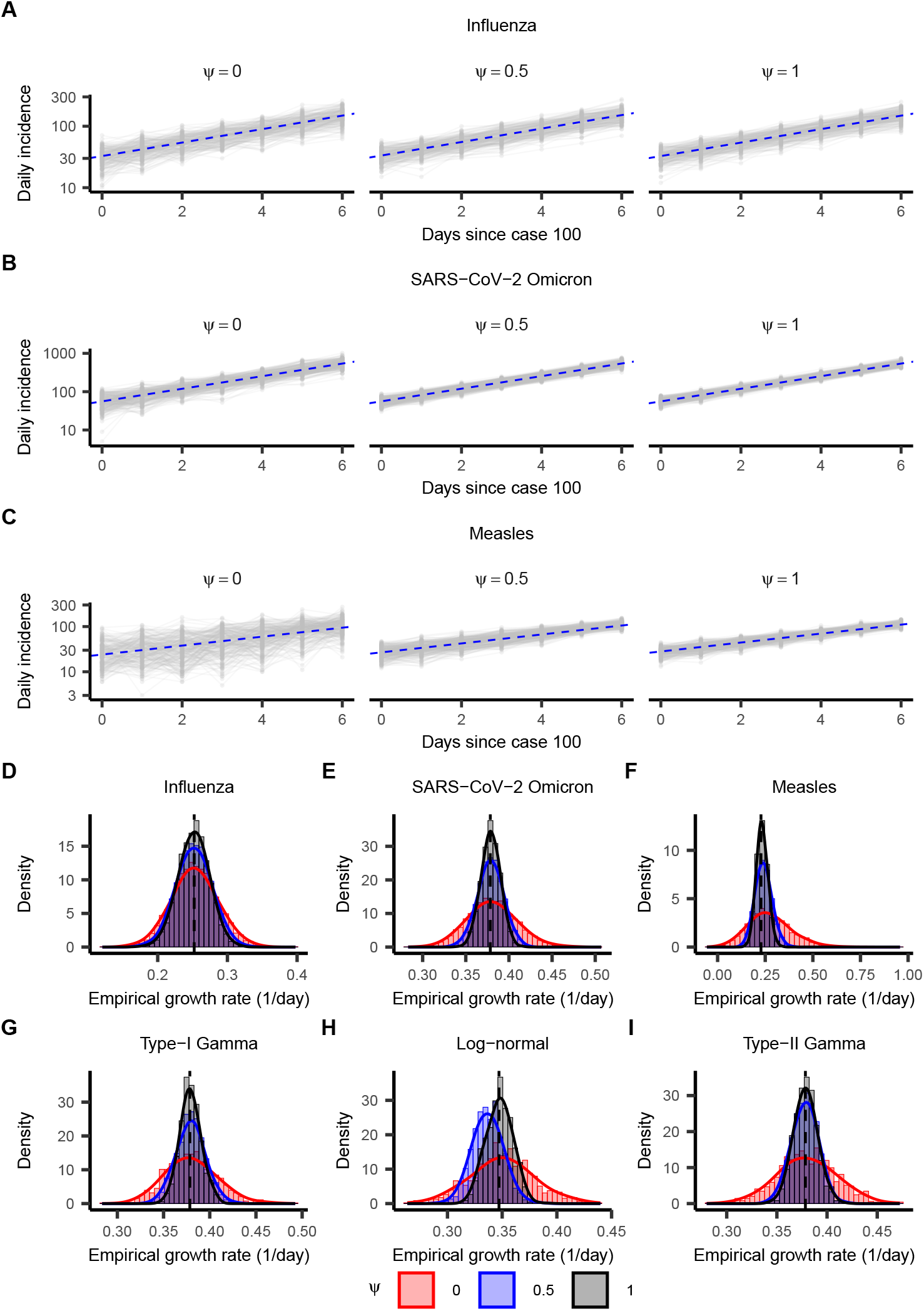
Burstiness and growth-rate estimation. (A–C) Log-scale daily infection counts (grey points connected by lines) from 250 epidemic simulations in an infinite population for influenza (A), SARS-CoV-2 omicron (B), and measles (C), for *ψ* ∈ {0, 0.5, 1} (sub-panels). The dashed blue line depicts the theoretical epidemic growth rate predicted by the Euler-Lotka equation. Bursty infectiousness increases the variation in daily case counts, especially for pathogens with high *R*_0_. (D–F) Estimated epidemic growth rates across pathogens and values of *ψ*. Histograms (bars) and kernel density estimates (solid lines) depict the distribution of the estimated growth rate (*r*) for influenza (D), SARS-CoV-2 omicron (E), and measles (F), for *ψ* ∈ {0, 0.5, 1} (colors), derived from 5,000 simulated epidemics per pathogen-*ψ* combination in an infinite population. The vertical reference line (dashed) marks the true growth rate derived from the Euler-Lotka equation. Bursty infectiousness (low *ψ*) makes estimates of *r* less certain due to inflation in the variance of daily case counts. (G–I) Estimated epidemic growth rates across *ψ* for SARS-CoV-2 and for the alternative burst models: the type-I Gamma burst model (G), the Log-normal model (H), and the type-II Gamma burst model (I). Histograms and lines are generated in the same way as in D–F. Across burst models, bursty infectiousness (low *ψ*) increases uncertainty in estimates of the growth rate *r*. The shift in the *ψ* = 0.5 histogram for the Log-normal burst model (H) reflects higher-moment discrepancies between the realized and target generation interval distribution, since the sum of independent log-normally distributed random variables is not perfectly log-normal.

**Extended Data Fig. 9.**
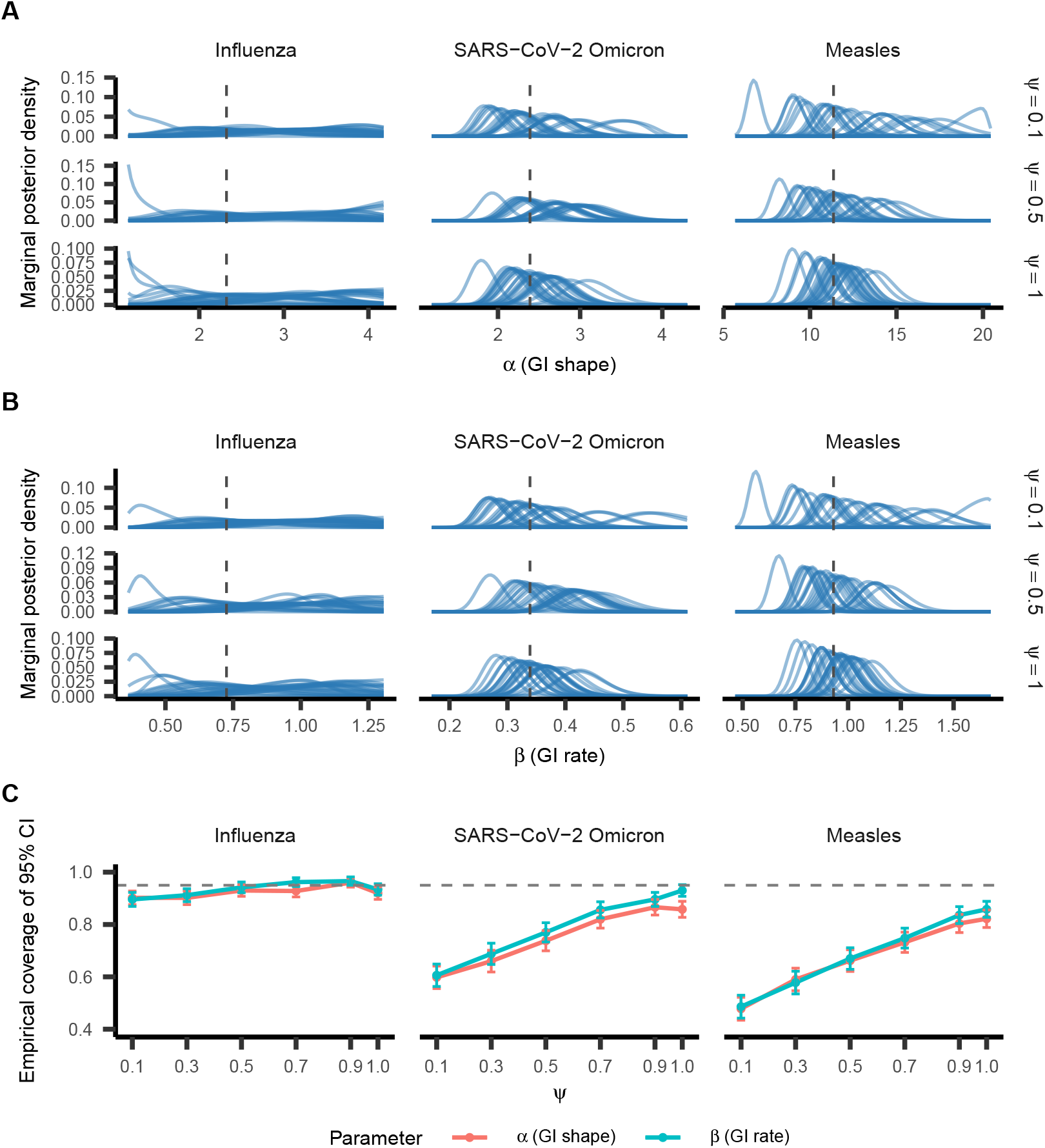
Burstiness and generation-interval estimation. Marginal posterior densities of the parameters *α* (A) and *β* (B) of the generation interval distribution, *g*(*τ*) ~ Gamma (*α, β*), for influenza (left-hand column), SARS-CoV-2 omicron (middle column), and measles (right-hand column), and for *ψ* = 0.1 (top row), *ψ* = 0.5 (middle row), and *ψ* = 1 (bottom row), generated by assuming that serial intervals reflect independent draws from *g*(*τ*) (*i*.*e*., ignoring the relatedness of “sibling” infections from a common infector). Dashed vertical lines indicate the parameter’s true value. Each panel overlays the posterior densities from 25 simulated datasets (**Methods**), each consisting of 100 transmission clusters for each pathogen-*ψ* combination. When serial intervals are treated as independent draws from the marginal distribution implied by *g*(*τ*) and the incubation period distribution, as is common practice, smaller values of *ψ* can lead to confident estimates of the generation interval parameters that miss the true value. (C) Empirical coverage of the shape (*α*) and rate (*β*) parameters (colored points connected with lines) of the generation interval distribution across values of *ψ* (horizontal axis) for influenza (left-hand panel), SARS-CoV-2 omicron (middle panel), and measles (right-hand panel). Error bars represent the ±1.96 standard error range of the estimate. Estimates were obtained from 500 simulated datasets consisting of 100 transmission clusters for each pathogen-*ψ* combination, assuming the serial intervals were independent draws from the marginal serial interval distribution implied by *g*(*τ*) and the incubation period distribution. Coverage was defined as the proportion of replicates in which the 95% credible interval contained the true value. The dashed horizontal line at 0.95 represents the expected coverage achieved when this independence assumption holds. The independence assumption leads to overconfident estimates of *α* and *β*, and this overconfidence intensifies as the individual GI distribution becomes more bursty (*ψ* → 0).

**Extended Data Table 1.**
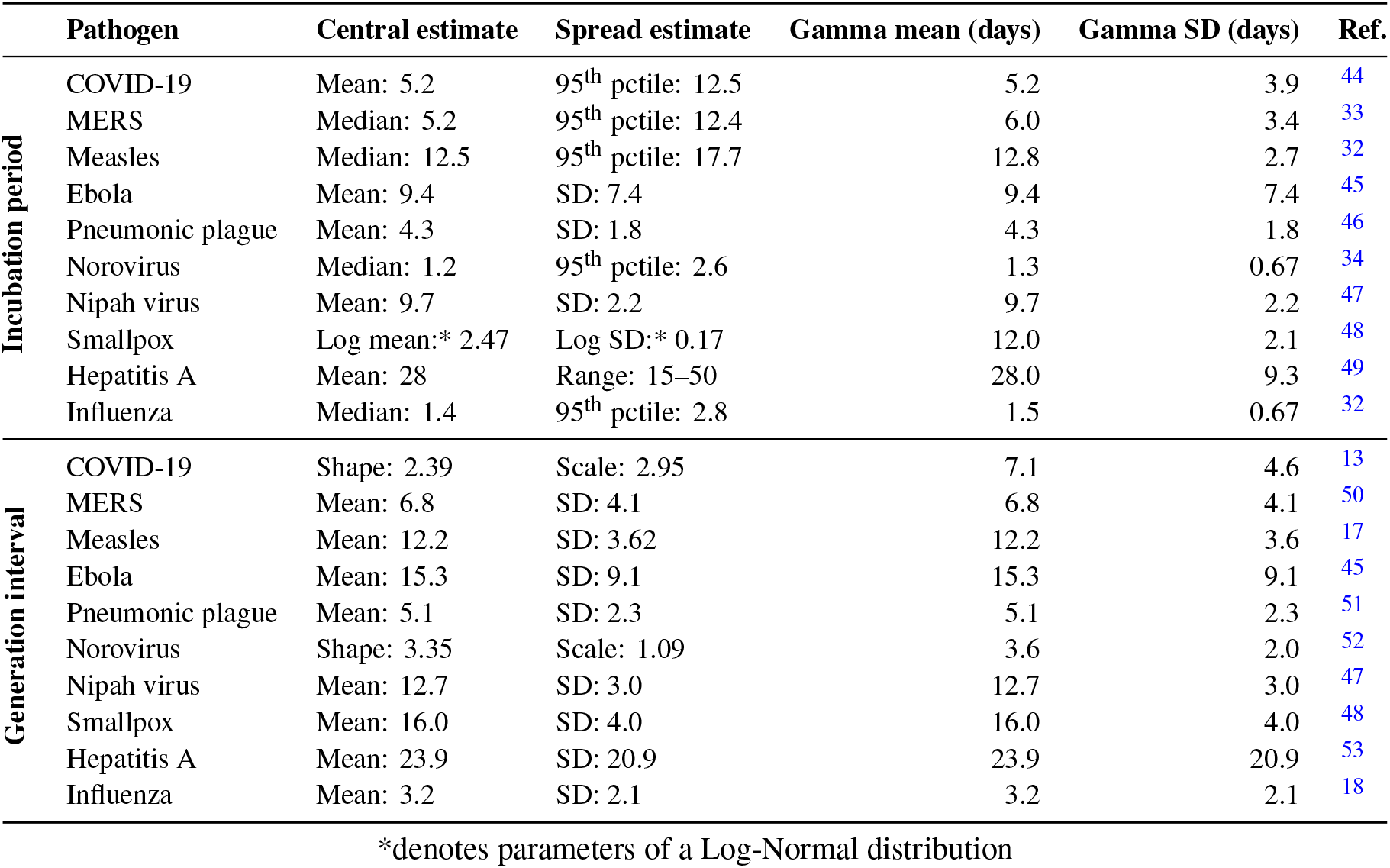
Incubation period and generation interval distribution parameters used for empirical estimation of *ψ*. Central and spread estimates for each pathogen were obtained from the cited reference. The Gamma mean and standard deviation were derived *via* moment- or quantile-matching. Generation interval parameters were used as inputs for inferring *ψ* only when empirical estimation of the generation interval distribution failed (influenza, COVID-19, and Ebola).

