## Supplementary Information for "How bursty infectiousness shapes epidemic dynamics"

Stephen M. Kissler

### 1 Supplementary Methods

#### 1.1 Notation: the individual generation interval distribution and its relationship to standard epidemiological models

The mean-field dynamics of an epidemic are determined by two fundamental quantities: the basic reproduction number, $R_0$, and the intrinsic generation interval (GI) distribution, $g(\tau)$[1], where $\tau$ represents time since infection. These can be combined into a single object, the "infectiousness profile" $A(\tau) = R_0\, g(\tau)$, which captures both the expected amount of transmission arising from an index case and its timing. $A(\tau)$ is the key object in the renewal equation framework for epidemic modeling[2]:

$$F(t) = \int_0^\infty F(t-\tau)S(t-\tau)A(\tau)\,d\tau \tag{1}$$

where $F(t)$ and $S(t)$ are the force of infection and density of susceptible individuals, respectively, at time $t$.

Analogously, at the individual level, the individual reproduction number, $\nu_i$, captures the expected number of infections person $i$ would produce in an otherwise susceptible population[3], and the individual intrinsic generation interval distribution, $g_i(\tau)$, captures the timing of those infections. These can be combined into the individual infectiousness profile (sometimes called the "infectiousness kernel"), $a_i(\tau) = \nu_i g_i(\tau)$[4,5]. The population-level quantities are the expectations of the individual-level ones, with $R_0 = E_i[\nu_i]$, $g(\tau) = E_i[g_i(\tau)]$, and $A(\tau) = E_i[a_i(\tau)]$. Substantial work has gone into understanding how variation in $\nu_i$ (*i.e.*, the area under $a_i(\tau)$) impacts epidemic dynamics, but the impact of variation in $g_i(\tau)$ (*i.e.*, how $a_i(\tau)$ is distributed over time), holding $R_0$ and $g(\tau)$ fixed and without confounding from variation in $\nu_i$, is less well understood.

Since $A(\tau)$ is an expectation over individuals, it need not resemble any single person's $a_i(\tau)$. The standard susceptible-exposed-infectious-recovered (SEIR) model provides a useful illustration: it assumes each person undergoes a latent period of length $x_i \sim \text{Exp}(\gamma_{\text{SEIR}})$ during which they are not infectious, followed by an infectious period of length $y_i \sim \text{Exp}(\alpha_{\text{SEIR}})$ during which they transmit at rate $\beta_{\text{SEIR}}$. The resulting "stepwise" individual infectiousness profile is

$$a_i(\tau) = \begin{cases} \beta_{\text{SEIR}} & \tau \in [x_i, x_i + y_i] \\ 0 & \text{otherwise} \end{cases} \tag{2}$$

Averaging over individuals yields the population-level infectiousness profile

$$A_{\text{SEIR}}(\tau) = E_i[a_i(\tau)] = \beta_{\text{SEIR}} \frac{\gamma_{\text{SEIR}}}{\gamma_{\text{SEIR}} - \alpha_{\text{SEIR}}} (e^{-\alpha_{\text{SEIR}}\tau} - e^{-\gamma_{\text{SEIR}}\tau}) \tag{3}$$

*i.e.*, a smooth curve that emerges from a collection of discontinuous $a_i(\tau)$ (**Extended Data Fig. 1**)[2]. Other choices of $a_i(\tau)$ can also produce the same $A_{\text{SEIR}}(\tau)$: for example, setting $a_i(\tau) \equiv A_{\text{SEIR}}(\tau)$ for all individuals gives a smooth, "sustained" profile in which each person's infectiousness follows the population-average distribution exactly. At the opposite extreme, $a_i(\tau) = (\beta_{\text{SEIR}}/\alpha_{\text{SEIR}})\delta_{\tau_i^*}(\tau)$, with $\delta_{\tau_i^*}(\tau)$ a delta function that has mass at time $\tau_i^*$ drawn from $g(\tau) = A_{\text{SEIR}}(\tau)/R_0$, gives a "spike" profile in which each person's infectiousness is concentrated at a single moment. All three families — stepwise, sustained, and spike — produce the same $A_{\text{SEIR}}(\tau)$ and hence the same deterministic, mean-field epidemic dynamics, but the resulting stochastic dynamics may differ.

#### 1.2 The burstiness parameter and the Gamma burst model

The population-level generation interval distribution $g(\tau)$ contains two distinct sources of variation: variation across individuals in when the infectiousness window falls, and variation within individuals in how long the infectiousness window lasts. Because both are folded into $g(\tau)$, two pathogens can share identical $R_0$ and $g(\tau)$ while differing markedly in the shape of $g_i(\tau)$.

Existing modeling frameworks cannot easily separate variation in $g_i(\tau)$ from $\nu_i$, or from the population-level $g(\tau)$ and $R_0$. The stepwise individual infectiousness profiles that conceptually underlie compartmental transmission models (*e.g.*, **Extended Data Fig. 1**) implicitly link $g_i(\tau)$ with $\nu_i$: individuals with longer infectious periods necessarily also have higher total infectiousness, making it difficult to disentangle the independent effects of $g_i(\tau)$ *vs.* $\nu_i$ on epidemic dynamics. Furthermore, existing modeling frameworks that allow for more precise tuning of $g_i(\tau)$ — *e.g.*, agent-based simulations, which allow for an arbitrary specification of $g_i(\tau)$, or the linear chain trick[2] , which allows for varying the typical width of $g_i(\tau)$ — also generally introduce associated changes to $g(\tau)$.

To isolate the impact of $g_i(\tau)$ on epidemic dynamics, we introduce the parameter, $\psi \in [0, 1]$, which specifies the proportion of $g(\tau)$'s variation that comes from within-individual *vs.* between-individual variation in infectiousness timing. When $\psi = 0$, there is no within-individual variation in infectiousness timing: all infections from an index case happen simultaneously (transmission is "bursty"), and the variation in $g(\tau)$ comes entirely from between-individual variation in when this infectious burst occurs. In this case, $g_i(\tau)$ is a delta function. When $\psi = 1$, there is no between-individual variation in infectiousness timing: everyone's individual GI distribution follows $g(\tau)$ exactly, so that the variation in $g(\tau)$ arises entirely from within-individual variation in the timing of infectiousness. In this case, $g_i(\tau) \equiv g(\tau)$. Critically, different choices of $\psi$ change the width of $g_i(\tau)$, but the population-level $g(\tau)$ is unchanged.

To examine the impact of $\psi$ on epidemic dynamics within a tractable framework, we introduce the (type-I) Gamma burst model (**Fig 1**). The model assumes that $g(\tau)$ follows a Gamma$(\alpha, \beta)$ distribution with rate $\beta = \alpha/\bar{g}$, where $\bar{g}$ is the mean generation time. The Gamma distribution is a common choice for modeling generation interval distributions, especially for acute infections[6] . Next, we define each person's individual intrinsic GI distribution as

$$g_i(\tau) = \begin{cases} 0 & \tau \leq l_i \\ f_\epsilon(\tau - l_i) & \tau > l_i \end{cases} \tag{4}$$

where $l_i \sim$ Gamma$((1-\psi)\alpha, \beta)$ is a random latent period and $f_\epsilon$ is the Gamma$(\psi\alpha, \beta)$ density. Equivalently, in terms of random variables, the secondary infections $j$ from an index case $i$ are distributed at times $\tau_{ij} = l_i + \epsilon_j$, where $l_i \sim$ Gamma$((1-\psi)\alpha, \beta)$ and $\epsilon_j \overset{iid}{\sim}$ Gamma$(\psi\alpha, \beta)$.

Since $l_i$ and $\epsilon_j$ are independent Gamma-distributed random variables with common rate $\beta$, their sum follows a Gamma$((1-\psi)\alpha + \psi\alpha, \beta)$ = Gamma$(\alpha, \beta)$ distribution; thus $E_i[g_i(\tau)] = g(\tau)$, as required. Furthermore, when $\psi \to 1$, the latent period vanishes ($l_i \to 0$) and $f_\epsilon$ approaches the full Gamma$(\alpha, \beta)$ density, which gives the "sustained" extreme (no between-individual variation). Similarly, when $\psi \to 0$, $f_\epsilon$ concentrates to a point mass and $g_i(\tau)$ becomes a delta function at $l_i \sim$ Gamma$(\alpha, \beta)$, which gives the "spike" extreme (no within-individual variation). The variance of the individual infectiousness burst is $\psi\alpha/\beta^2$, while the variance of the full generation interval distribution is $\alpha/\beta^2$; thus, $\psi$ gives exactly the fraction of $g(\tau)$'s variance that is attributable to within-infector variation in infection timing.

We generally assume that $\psi$ is a single value shared by all individuals for a given pathogen, but the framework is more general: the equal-rate Gamma addition property ensures that drawing unique $\psi_i$ for each infector from a distribution on $[0, 1]$ still yields $E_i[g_i(\tau)] = g(\tau)$; thus, it is possible to model scenarios where individuals have a mixture of narrow and broad individual GI distributions. For example, we might consider $\psi_i \sim$ Beta$(0.5, 0.5)$, a Ushaped distribution where roughly half of the population has bursty infectiousness and half has sustained infectiousness; this still yields the same $g(\tau) \sim \text{Gamma}(\alpha, \beta)$ in expectation.

This Gamma burst model is just one of many possible choices; we introduce two additional burst models in Section 1.14.

##### 1.3 Incorporating interpersonally- and time-varying contacts

For $A(\tau)$ and $a_i(\tau)$ to be functions of $\tau$ (time since infection) alone, and not also of calendar time $t$, it is necessary to assume that interpersonal contact rates do not vary over time. We follow this convention for most of our analyses. However, the interaction between bursty infectiousness and time-varying contacts may be consequential: a burst of infectiousness that happens to coincide with a high-contact period could create superspreading, even if a person does not have a systematically higher-than-average infectiousness or number of contacts. Thus, we briefly introduce a generalization that accounts for time-varying contacts. We also introduce two specific contact models that serve as useful illustrations.

The fundamental epidemiological parameters $R_0$, $g(\tau)$, $\nu_i$, and $g_i(\tau)$ are all implicitly defined with respect to a reference, uniform contact level, which we denote $\bar{c} \equiv 1$. To allow for time-varying contacts, we introduce a contact process $\mathbf{c}(t)$: a nonnegative, wide-sense stationary stochastic process with time-mean $E_t[\mathbf{c}(t)] = 1$. Each person $i$ has a contact modulator $c_i(t)$ that is a realization of $\mathbf{c}(t)$, representing the proportional scaling of that person's contact rate relative to the reference level at calendar time $t$ (*e.g.*, **Fig. 3A–C, Extended Data Fig. 3A–C**). The constraint $E_t[\mathbf{c}(t)] = 1$ is an ensemble average over all possible realizations at time $t$; individual realizations $c_i$ need not have time-average 1, allowing for individuals with persistently higher or lower contact rates than the population average. We additionally define the "biological infectiousness", $b_i$, as the expected number of secondary infections person $i$ would produce under $\bar{c} \equiv 1$. The individual infectiousness profile thus generalizes to

$$a_i(t_i, \tau) = b_i g_i(\tau) c_i(t_i + \tau) \tag{5}$$

where $t_i$ is person $i$'s infection time. If $c(t) \equiv \bar{c} = 1$, this reduces back to the familiar $a_i(\tau) = \nu_i g_i(\tau)$, with $b_i = \nu_i$. More generally,

$$\nu_i = b_i \int_0^\infty g_i(\tau) c_i(t_i + \tau)\, d\tau \tag{6}$$

*i.e.*, the individual reproduction number is a function of the person's inherent biological infectiousness $b_i$ as well as the interaction between their infectiousness timing and their contact process. Likewise, the realized individual generation interval distribution is

$$g_i^{\text{re}}(t_i, \tau) = \frac{g_i(\tau) c_i(t_i + \tau)}{\int_0^\infty g_i(u) c_i(t_i + u)\, du} \tag{7}$$

(**Fig. 3A–C, Extended Data Fig. 3A–C**). So, an equivalent rendering of Eq. 5 is

$$a_i(t_i, \tau) = \nu_i g_i^{\text{re}}(t_i, \tau) \tag{8}$$

If we assume $c_i$ is independent of $(b_i, g_i)$, then we can reconstruct population-level quantities. The basic reproduction number, which is the expected individual reproduction number, is also the mean biological infectiousness:

$$R_0 = E[\nu_i] = \int_0^\infty E[b_i\, g_i(\tau)\, c_i(t_i+\tau)]\, d\tau \tag{9}$$

$$= \int_0^\infty E[b_i\, g_i(\tau)] \cdot \underbrace{E[c_i(t_i+\tau)]}_{=1}\, d\tau \tag{10}$$

$$= E\left[b_i \int_0^\infty g_i(\tau) d\tau\right] = E[b_i] \tag{11}$$

The population-level intrinsic generation interval distribution is the $b_i$-weighted average of individual intrinsic generation interval distributions:

$$g(\tau) = E[b_i g_i(\tau)]/R_0 \tag{12}$$

Combining these, the population-level infectiousness profile is

$$A(t,\tau) = R_0\, g(\tau)\, C(t+\tau) \tag{13}$$

where $C(s) = E_i[c_i(s)]$ is the population-average contact level at calendar time $s$. We note that $b_i$ and $g_i$ need not be independent: biologically, wider individual GI distributions may correlate with higher total infectiousness. The generation interval distribution $g$ accounts for any such correlation; when $b_i$ and $g_i$ are independent, $g$ reduces to the unweighted average $E[g_i(\tau)]$. When $C$ is constant, $A$ depends only on $\tau$ and the renewal equation is time-homogeneous. When $C$ varies in time, $A$ depends on both $t$ and $\tau$.

For illustration, we consider two contact models. We define the "periodic" contact model

$$c_i(t) = 1 - \zeta \cos\left(\frac{2\pi t}{T_c}\right) \quad \forall i \tag{14}$$

with amplitude $\zeta \in [0,1]$ and period $T_c > 0$ (**Fig. 3A–C**). In this model, all individuals share the same deterministic contact function; this is a degenerate case of the contact process $\mathbf{c}(t)$ in which every realization is identical. While there is temporal variation within each person's contacts, there is no variation across individuals. The process' stationarity is achieved by considering a random infection time $t_i$ which is uniformly distributed over one period — while this assumption may not hold in general, we use it as a simplifying assumption for analytic tractability[7] . At the population level, $C(t) = E_i[c_i(t)] = 1 - \zeta \cos(2\pi t/T_c)$.

We also define the "stochastic" contact model, where each person's $c_i(t)$ is a piecewise-constant process that switches to a new level at arrivals of a Poisson process with rate $\lambda$ (**Extended Data Fig. 3A–C**). The contact levels are drawn independently from a Gamma($\sigma, \sigma$) distribution, which has mean 1 (satisfying the constraint $E_t[\mathbf{c}(t)] = 1$) and variance $1/\sigma$. This creates a two-parameter family indexed by the dispersion parameter $\sigma > 0$ and switching rate $\lambda$. Smaller $\sigma$ yields more heterogeneous contacts, while large $\sigma$ concentrates the contact levels near 1. As $\sigma \to \infty$, the distribution collapses to a point mass at 1, recovering the constant-contact reference process. In contrast to the periodic model, each person's contact trajectory is unique, but the population average is constant in time: $C(t) = E_i[c_i(t)] = 1$.

##### 1.4 Simulation approach

**Stochastic simulations.** We simulated epidemics using a stochastic, individual-based algorithm. The simulation model has a modular design: an infection attempt generator produces a set of transmission attempts for each infected individual, and a population-level simulation script processes these attempts to propagate the epidemic. We implemented two variants: a finite-population model with susceptible depletion, and an infinite-population model where every infection attempt succeeds. Both are implemented in C++. Unless otherwise stated, simulations use a population size of $N = 10,000$ with 5,000 replicate simulations per scenario.

To generate infection attempts, the algorithm proceeds as follows:

1. **Draw the number of infection attempts.** The number of secondary infection attempts $\chi_i$ is drawn from a Poisson($\nu_i$) distribution, where $\nu_i$ is the individual reproduction number. Throughout, we set $b_i \equiv R_0$ for all individuals (where $\nu_i = b_i$ when contacts are uniform, otherwise $\nu_i$ is given by Eq. 6), so that variation in realized transmission arises solely from infectiousness timing and contact dynamics, and not from inherent infectiousness.

2. **Draw the latent period.** A latent period $l_i$ is drawn from a Gamma($(1-\psi)\alpha, \beta$) distribution, representing the delay between infection and the onset of transmissibility.

3. **Draw the timing of each infection attempt.** For each infection attempt $j \in 1, \ldots, \chi_i$, an independent $\epsilon_j$ is drawn from a Gamma($\psi\alpha, \beta$) distribution. The time of infection attempt $j$ relative to the index case's infection is thus $\tau_{ij} = l_i + \epsilon_j$.

4. **Thin for time-varying contacts (if applicable).** When a time-varying contact process $c_i(t)$ is in place, each infection attempt at calendar time $t_i + \tau_{ij}$ (where $t_i$ is the index case's infection time) is accepted with probability $c_i(t_i + \tau_{ij})/c_{max}$, where $c_{max}$ is the supremum of the contact process (an implementation of Poisson thinning). For the periodic contact model, all individuals share the same deterministic contact function $c_i(t) = 1 - \zeta\cos(2\pi t/T_c)$, and thus $c_{max} = 1 + \zeta$. For the stochastic contact model, each person's piecewise-constant contact trajectory is generated by drawing switching times from a Poisson process with rate $\lambda$ and contact levels from a Gamma($\sigma, \sigma$) distribution (or a truncated Gamma($\sigma, \sigma$) distribution when a gathering size restriction is in place). We generate the contact process over an interval $[0, 5\,\bar{g}]$ to ensure enough runway to accommodate all infection attempts. Thinning is performed against the realized maximum contact level over that individual's contact trajectory.

5. **Thin for detect-and-isolate interventions (if applicable).** When a detect-and-isolate intervention is in place, an individual-specific detection time $D_i$ (relative to infection onset) is drawn, according to the detection mechanism in effect (symptom-based or screening-based). All infection attempts occurring after detection ($\tau_{ij} > D_i$) are removed with probability $\eta$, representing the effectiveness of post-detection isolation.

In a finite population of size $N$, the epidemic is propagated according to the following rules:

1. **Initialization.** A single index case is infected at time $t = 0$. All other $N - 1$ individuals begin as susceptible.

2. **Queue creation.** Each infection attempt is stored as a pending event with its scheduled calendar time $t_i + \tau_{ij}$. Infection attempts are maintained in a priority queue, ensuring that the earliest pending event is always processed next.

3. **Infection attempt processing.** The earliest event is selected from the queue. A target individual is selected uniformly at random from the population. If the target is susceptible, infection occurs: the target is marked as infected, and its own infection attempts are generated and queued. If the target was previously infected, the attempt fails (it is "wasted" on a non-susceptible individual).

4. **Ending the epidemic.** The simulation terminates when the event queue is empty or when a pre-specified number of infections is reached.

To simulate epidemics in an infinite population, all infection attempts succeed: in Step 3, every queued event produces a new infection, with no target selection or susceptibility check.

**Deterministic simulations.** To generate the expected (mean-field) epidemic trajectory given $R_0$ and $g(\tau)$, we numerically solved the standard renewal equation $i(t) = S(t)R_0 \int_0^t i(t-\tau)g(\tau)d\tau$, where $i(t)$ is the incidence at time $t$ and $S(t) = 1 - \int_0^t i(s)\, ds$ is the susceptible density[2] . We discretized on a uniform grid $\{t_0, t_1, t_2, \ldots, t_K\}$ with step $dt = 0.01$ days. We initialized with a single index case at $t_0 = 0$ whose forward contributions enter through an explicit kernel $g(t_k)/N$ (where $N$ is the population size). The approximate incidence at time $t_k$ can be calculated as a Riemann sum: $i_{t_k} = S_{t_{k-1}} R_0 \left( g(t_k)/N + \sum_{j=1}^{k-1} g(t_j) i_{t_{k-j}} dt \right)$. The cumulative incidence at time $t_k$ is then $C_{t_k} = C_{t_{k-1}} + i_{t_k} dt$.

##### 1.5 Empirical estimation of $\psi$

Ideally, estimates of $\psi$ would be informed by detailed contact-tracing data where all infections from each index case are identified and where infection times are precisely recorded. In practice, such data are rarely available; so, taking inspiration from approaches to estimate $g(\tau)$[8] , we use serial intervals (symptom onset lag between an index case and secondary infection) to inform the value of $\psi$.

**Mathematical approach.** The serial interval $s_{ij}$ between an infector $i$ and secondary case $j$ can be expressed in terms of the generation interval $\tau_{ij}$ and the incubation periods $d_i$ and $d_j$:

$$s_{ij} = \tau_{ij} + d_j - d_i \tag{15}$$

Under the Gamma burst model, $\tau_{ij} = l_i + \epsilon_j$, where the latent period $l_i \sim \text{Gamma}((1-\psi)\alpha, \beta)$ is shared across all of infector $i$'s secondary infections and $\epsilon_j \overset{iid}{\sim} \text{Gamma}(\psi\alpha, \beta)$. We assume that the incubation periods are also Gamma-distributed: $d \sim \text{Gamma}(a_{obs}, b_{obs})$. Substituting into Eq. 15 and grouping terms gives:

$$s_{ij} = (l_i - d_i) + (\epsilon_j + d_j) \equiv \tilde{l}_i + \tilde{\epsilon}_j \tag{16}$$

Given $K$ clusters consisting of $\chi_i$ infections for a given pathogen, the likelihood of $\psi$ is

$$L(\psi) = \prod_{i=1}^{K} \int_{-\infty}^{\infty} f_{\tilde{l}}(u;\psi) \prod_{j=1}^{\chi_i} f_{\tilde{\epsilon}}(s_{ij} - u;\psi)\, du \tag{17}$$

where

$$f_{\tilde{l}}(\bullet;\psi) \quad \text{is the density of} \quad \tilde{l}_i = \text{Gamma}((1-\psi)\alpha,\beta) - \text{Gamma}(a_{obs}, b_{obs}), \quad \text{and}$$
$$f_{\tilde{\epsilon}}(\bullet;\psi) \quad \text{is the density of} \quad \tilde{\epsilon}_j = \text{Gamma}(\psi\alpha,\beta) + \text{Gamma}(a_{obs}, b_{obs})$$

We note that this derivation assumes a single value of $\psi$ is shared across all infectors for each pathogen.

**Data sources.** We obtained serial interval data from 315 clusters across 10 pathogens (951 total serial intervals) from the OutbreakTrees database[9] . We extracted Gamma shape and rate parameters for the incubation period distributions from the literature (**Extended Data Table 1**), either directly or by matching moments or quantiles to reported values. Generation interval parameters $\alpha$ and $\beta$ were inferred from the serial interval clusters: assuming generation and incubation intervals are independent, $\bar{s} = \bar{g}$ (where $\bar{s}$ is the mean serial interval), and $\text{Var}[s] = \text{Var}[g] + 2\text{Var}[d]$ (*i.e.*, the variance of the serial interval distribution equals the variance of the generation interval distribution plus twice the variance of the incubation period distribution). The values of $\alpha$ and $\beta$ were then obtained from the empirical $\bar{g}$ and $\mathrm{Var}[g]$ using moment matching. If the empirical $\widehat{\mathrm{Var}}[g] < 0$, we fell back to literature-reported values for the generation interval distribution (**Extended Data Table 1**); this happened for influenza, Ebola, and COVID-19.

**Computation.** For numerical implementation, we evaluated the posterior of $\psi$ at 101 equally spaced values on $[0, 1]$. For each $\psi$ value, we precomputed the marginal densities $f_{\tilde{l}}$ and $f_{\tilde{\epsilon}}$ on a fine grid, with $f_{\tilde{l}}$ spanning $[-(\bar{d}+5\sigma_d), \bar{g}+5\sigma_g]$ and $f_{\tilde{\epsilon}}$ spanning $[0, \bar{g}+\bar{d}+5(\sigma_g+\sigma_d)]$, where $\bar{g}$, $\sigma_g$, $\bar{d}$, and $\sigma_d$ are the means and standard deviations of the generation interval and incubation period distributions, respectively. For each infection cluster, the integral (Eq. 17) was evaluated as a left Riemann sum over the precomputed values of $f_{\tilde{l}}$, with the log-sum-exp transformation applied before summing to avoid numerical underflow. $f_{\tilde{\epsilon}}$ was evaluated at off-grid arguments $s_{ij} - u_k$ by linear interpolation from the precomputed grid. Log-likelihoods were summed across all clusters within each pathogen, then exponentiated and normalized over the $\psi$ grid to obtain the posterior, equivalent to Bayesian inference with a flat prior on $\psi \in [0, 1]$.

##### 1.6 Identifiability of $\psi$

Estimates of $\psi$ can be biased due to various factors, including imperfect ascertainment and mis-specification of the detection delay distribution (*e.g.*, the incubation period). Here, we use simulated datasets, matched to the cluster-size characteristics and known epidemiological parameters of the OutbreakTrees dataset[9] , to assess the identifiability of $\psi$.

Using the same likelihood approach as in Section 1.5, we performed three identifiability analyses: an assessment of identifiability across $\psi$ values for each pathogen, using empirical GI distribution parameters and literature incubation period parameters; an assessment of sensitivity to imperfect ascertainment, with ascertainment rates ranging from 30% to 100%; and an assessment of sensitivity to mis-specification of the incubation period distribution, with the distribution's variance ranging from 0.5× to 2× the literature value.

**Identifiability of $\psi$ across pathogens.** For each pathogen, we used the empirically estimated generation interval parameters $\hat{\alpha}$ and $\hat{\beta}$, derived from the observed serial intervals, along with literature values for the incubation period distribution ($a_{\text{obs}}$ and $b_{\text{obs}}$), to generate synthetic infection clusters. For each of 21 equally-spaced $\psi$-values on [0,1], and for each pathogen, we generated 500 synthetic datasets by (i) bootstrap-sampling cluster sizes with replacement from the empirical cluster size distribution for that pathogen, and (ii) drawing serial intervals under the Gamma burst model with incubation-period delays. For each synthetic dataset $s$, we re-computed the posterior over $\psi$ and extracted the posterior mean $\mu_s$ and 95% credible interval $[\mathrm{lwr}_s, \mathrm{upr}_s]$. Then, for each pathogen-$\psi$ pair, we characterized (a) the mean of posterior means $\mu_s$, to capture the expected posterior mean across all simulations; (b) the 5%–95% range of the posterior means $\mu_s$, to capture the range of plausible posterior means; and (c) and the means of the lower and upper credible interval bounds $\mathrm{lwr}_s$ and $\mathrm{upr}_s$, to capture the typical credible interval spread (**Extended Data Fig. 2A**). This gives the range of posterior estimates that could plausibly arise from the empirical sample given any "true" $\psi$-value.

**Sensitivity of $\psi$ inference to imperfect ascertainment.** To assess sensitivity to incomplete ascertainment of secondary cases, we repeated the same identifiability assessment as before, but with ascertainment rates $p_{\text{asc}} \in \{0.3, 0.5, 0.7, 1.0\}$, with 200 replicates per pathogen-$\psi$-$p_{\text{asc}}$ combination. To simulate incomplete ascertainment, we set the true $R_0 = \bar{\chi}_i / p_{\text{asc}}$, where $\bar{\chi}_i$ is the mean empirical cluster size, so that the expected simulated cluster size (after ascertainment thinning) matched the empirical mean. After generating secondary infections for each cluster, we thinned them with binomial probability $p_{\text{asc}}$ and used these thinned serial intervals for inference (**Extended Data Fig. 2B**).

**Sensitivity of $\psi$ inference to mis-specified detection delays.** To assess sensitivity to mis-specified incubation period parameters, we generated 100 synthetic datasets for each pathogen and for each of three incubation period distribution scenarios: setting the variance of the distribution ($a_{\text{obs}}/b^2_{\text{obs}}$) to 0.5×, equal to, and 2× the literature value, while holding the incubation period distribution mean ($a_{\text{obs}}/b_{\text{obs}}$) fixed and setting $\psi$ to its empirical posterior mean (Section 1.5). We then ran the same $\psi$ inference as before (**Extended Data Fig. 2C**).

**Summary of findings.** The empirical $\psi$ estimates are well-localized for most pathogens (measles, COVID-19, influenza, MERS, smallpox, norovirus), insensitive to ascertainment bias, and qualitatively robust to incubation period misspecification — though incubation period misspecification does yield non-trivial quantitative shifts in the estimates of $\psi$. Based on the main identifiability analysis and the available data, we have reasonable confidence that measles, COVID-19, and influenza have high burstiness (low $\psi$); MERS and smallpox have intermediate burstiness (intermediate $\psi$); and norovirus has low burstiness (high $\psi$). This is illustrated in **Extended Data Fig. 2A**, where the range of $\psi_{\text{true}}$ values for which the simulation-recovered posteriors (blue) overlap with the empirical posterior (red) is narrow, indicating that the empirical data localize $\psi$ to a small interval. For Hepatitis A, Ebola, Pneumonic plague, and Nipah virus, the simulation-recovered posteriors overlap with the empirical posteriors across a wide range of $\psi$-values, indicating that the available data are insufficient to localize $\psi$ for these pathogens. Posterior estimates from the simulations were robust to different levels of ascertainment (**Extended Data Fig. 2B**). Mis-specification of the incubation period produced substantial shifts in posterior estimates of $\psi$; for example, the mean posterior estimate for MERS ranged from 0.47 when the assumed incubation period was too narrow (0.5× variance), to 0.24 when it was too wide (2× variance), with a correctly specified baseline of 0.35 (**Extended Data Fig. 2C**). Nevertheless, the relative ordering and approximate range of $\psi$-values across the pathogens was robust to mis-specification of the incubation period distribution.

##### 1.7 Estimating the SARS-CoV-2 infectiousness window from viral kinetics data

**Data and preprocessing.** For each documented infection $i$ we assembled the longitudinal series of Ct values $y_{ij}$ measured at times $t_{ij}$ (days, with day 0 corresponding to the day of the lowest observed Ct value/highest observed viral load for infection $i$). Following our prior work[10], each series was trimmed to the positive tests together with up to two flanking negative tests on either side, so that the model is informed by the shoulders of each trajectory without being dominated by long runs of undetectable samples. We modeled the data on the "drop" scale, $d_{ij} = \text{LOD} - y_{ij}$, with LOD = 40; a larger drop corresponds to a higher viral load.

**Viral trajectory model.** The expected drop for infection $i$ at time $t$ is the piecewise-linear "tent"

$$v_i(t) = \begin{cases} \dfrac{\delta_i}{\omega_i^p}\left(t - (\tau_i - \omega_i^p)\right), & t \le \tau_i, \\[2ex] \delta_i - \dfrac{\delta_i}{\omega_i^r}(t - \tau_i), & t > \tau_i, \end{cases} \tag{18}$$

where $\tau_i$ is the peak time, $\delta_i > 0$ is the peak drop below the LOD, and $\omega_i^p, \omega_i^r > 0$ are the proliferation and clearance durations. Outside the interval $[\tau_i - \omega_i^p,\ \tau_i + \omega_i^r]$ the trajectory is taken to lie at or below the LOD (drop $\le 0$), i.e. undetectable.

**Hierarchical priors.** The peak magnitude and the proliferation and clearance phase durations were given log-normal population distributions, parameterized around fixed midpoints $(\delta_0, \omega_0^p, \omega_0^r) = (20, 5, 12)$:

$$\delta_i = \delta_0 \exp(\mu_\delta + \sigma_\delta z_i^\delta), \qquad \omega_i^p = \omega_0^p \exp(\mu_p + \sigma_p z_i^p), \qquad \omega_i^r = \omega_0^r \exp(\mu_r + \sigma_r z_i^r), \tag{19}$$

with standard-normal individual effects $z_i^\cdot \sim \text{Normal}(0, 1)$ (non-centered parameterization). We placed weakly informative priors $\mu_\bullet \sim \text{Normal}(0, 0.25)$ and $\sigma_\bullet \sim \text{Normal}^+(0, 0.25)$ on the log-scale hyperparameters, $\tau_i \sim \text{Normal}(0, 2)$ on the peak times, and $\sigma \sim \text{Normal}^+(0, 0.5)$ on the observation scale (throughout this section, the second parameter of the Normal density corresponds to the standard deviation). This is a single-category fit, where all infections share one common population mean, with no adjustment for infection order, vaccination, or variant.

**Observation model.** Given the expected Ct value at a given time, $\mu_{ij} = v_i(t_{ij})$, each observation was modeled as a two-component mixture,

$$p(d_{ij} \mid \cdot) = \lambda\, \text{Exponential}\big(d_{ij};\, \text{rate} = 1/\phi\big) + (1-\lambda) \begin{cases} \text{Normal}(d_{ij};\, \mu_{ij}, \sigma), & d_{ij} > 0, \\ \Phi\left(\dfrac{0-\mu_{ij}}{\sigma}\right), & d_{ij} = 0, \end{cases} \tag{20}$$

where the Gaussian component is censored at the LOD (observations exactly at the LOD, $d_{ij} = 0$, contribute the normal CDF mass below zero), and the small-weight exponential component ($\lambda = 0.01$, mean $\phi = 1/\ln 10 \approx 0.43$ Ct) accounts for false negatives (which may register either at or near the LOD) with probability $\lambda$.

**Inference.** We sampled the posterior with Hamiltonian Monte Carlo (NUTS) in Stan[11] via `rstan`, running 4 chains of 4,000 iterations each (2,000 warmup). Convergence of the population-level parameters was assessed by the Gelman-Rubin $\hat{R}$ statistic[12] (all $\hat{R} < 1.01$); there were no divergent transitions.

**Time above a viral load threshold.** Because $v_i$ is piecewise-linear, the duration for which infection $i$ exceeds a threshold specified as a drop $D = \text{LOD} - \text{Ct}^*$ is

$$W_i(\text{Ct}^*) = (\omega_i^p + \omega_i^r)\, \frac{\max(\delta_i - D,\ 0)}{\delta_i}, \tag{21}$$

obtained from the width of the tent at height $D$ on each leg. We evaluated Eq. (21) at every integer Ct from 40 to 20 for each posterior draw, yielding a posterior distribution of the infectious window per infection. For each distribution, we extracted the posterior mean trajectory. We report, per threshold, the proportion of infections whose posterior mean trajectory reaches it and the median and 95% quantiles of time spent above that threshold across all infections' posterior mean trajectories (**Figure 2E**).

**Relating cycle threshold to infectiousness.** Based on various viral culture studies, we adopt two approximate reference thresholds linking Ct values with possible infectiousness. First, we set the threshold of "bare infectiousness" at Ct ≈ 32, a ceiling above which virus is rarely culturable and disease transmission does not appear to occur. In one study, viable virus was recovered in 0 of 33 samples below $10^5$ genome equivalents per mL (GE/mL) of viral transport media[13], which corresponds to a Ct value of roughly 31 on our assay. In another, 8% of tested samples were culture-positive at Ct > 35; a logistic fit to the available data suggested viral culture positivity was ≈ 20% for Ct > 32[14]. A third study found no viral culture positivity above Ct = 34[15], and a fourth found no culture positivity above Ct = 24[16]. Operationally, occupational testing programs in the National Basketball Association and National Football League adopted Ct thresholds of 31.7 and 35 as indicators of non-infectiousness, respectively[17,18]; no transmissions were reported from individuals with Ct values above these thresholds.

Second, we note that Ct ≈ 26 corresponds to a roughly 50% chance of viral culture positivity, which we take to be an indicator of potentially moderate infectiousness. One study recovered viable virus in 3 of 11 (27%) of samples with viral loads of $10^6$–$10^7$ GE/mL (Ct 28–24, on our assay); and in 12 of 17 (71%) samples above $10^7$ GE/mL (Ct < 24 on our assay)[13]. In another, a logistic fit suggests 50% viral culture positivity fell near Ct = 28 on their assay, approaching 100% by Ct < 24[14]. A third study reported roughly 50% viral culture positivity near Ct = 28[15].

We note that Ct values are assay- and platform-dependent, and that viral culturability is only a proxy for (specifically, a necessary condition for) infectiousness. These thresholds are therefore necessarily approximate. By reporting the time spent above each integer Ct value from 40-20, we aim to avoid sensitivity to any particular choice of infectiousness threshold. Our central conclusion, that the interval of appreciable infectiousness is substantially shorter than the width of the generation interval distribution, is insensitive to the exact threshold: it holds for any cutoff more stringent than the limit of detection (Ct ≈ 40).

##### 1.8 Coincidence superspreading

Coincidence superspreading arises when, by chance, a person's infectiousness window happens to align with a period of high contacts. To estimate how $\psi$ and a time-varying contact process impact overdispersion in secondary infection counts, we derived an analytic expression for the overdispersion parameter $k$[3] in terms of the burstiness parameter $\psi$. We begin with the standard assumption that the number of offspring $\chi_i$ produced by an index case $i$ is Poisson-distributed:

$$\chi_i \sim \text{Poisson}(\nu_i) \tag{22}$$

where $\nu_i$ is the individual reproduction number for infector $i$. When $\nu_i$ is uniform across people ($\nu_i \equiv R_0$), then the $\chi_i$ are truly Poisson-distributed. Overdispersion occurs when $\nu_i$ varies across individuals. To capture this overdispersion, the standard approach is to describe $\chi_i$ using a Negative Binomial distribution:

$$\chi_i \sim \text{NegBin}(R_0, k) \tag{23}$$

which has expectation $R_0$ and variance $R_0(1 + R_0/k)$. Thus, $k$ captures overdispersion, with $k \to \infty$ yielding Poisson-distributed $\chi_i$ and $k \to 0$ yielding increasingly heavier Negative Binomial tails (*i.e.*, more overdispersion). The Negative Binomial expression is exact when $\nu_i$ are Gamma-distributed; otherwise, it is an approximation to the distribution of $\chi_i$. Either way, the parameter $k$ can be obtained *via* moment matching by re-arranging the variance expression:

$$k = \frac{R_0^2}{\text{Var}[\chi_i] - R_0} \tag{24}$$

Furthermore, since

$$\text{Var}[\chi_i] = \text{Var}[E[\chi_i|\nu_i]] + E[\text{Var}[\chi_i|\nu_i]] = \text{Var}[\nu_i] + E[\nu_i] = \text{Var}[\nu_i] + R_0 \tag{25}$$

the expression for $k$ simplifies to

$$k = \frac{R_0^2}{\text{Var}[\nu_i]} \tag{26}$$

Next, if we assume everyone's inherent biological infectiousness is the same ($b_i \equiv R_0$), we have

$$v_i = R_0 \int_0^\infty g_i(\tau) c_i(t_i + \tau) d\tau := R_0 \tilde{c}_i \tag{27}$$

Plugging in to 26 gives

$$k = \frac{1}{\text{Var}[\tilde{c}_i]} \tag{28}$$

Thus, the overdispersion $k$ depends only on the variance of $\tilde{c}_i = \int_0^\infty g_i(\tau) c_i(t_i + \tau) d\tau$, which can be interpreted as the effective contact level for infector $i$ over the course of their infection.

**Deriving an expression for Var[$\tilde{c}_i$].** The random variable $\tilde{c}_i$ inherits randomness from both $g_i(\tau)$ and $c_i(t_i + \tau)$. To manage this, we again apply the Law of Total Variance:

$$\begin{aligned}
\text{Var}[\tilde{c}_i] &= \text{Var}[E[\tilde{c}_i | g_i]] + E[\text{Var}[\tilde{c}_i | g_i]] && (29)\\
&= \text{Var}[1] + E[\text{Var}[\tilde{c}_i | g_i]] && (30)\\
&= E[\text{Var}[\tilde{c}_i | g_i]] && (31)
\end{aligned}$$

where the second line follows from the fact that $E[c_i(t)] = 1$, and $g_i$ integrates to 1, so $E[\tilde{c}_i | g_i] = 1$ regardless of $g_i$. Thus, we are interested in

$$\tilde{c}_i | g_i = \int_0^\infty g_i(\tau) c_i(t_i + \tau) d\tau \tag{32}$$

where $g_i$ is known (*i.e.*, conditioned upon). This has the form of a classic problem in signal processing: $c_i$ is a wide-sense stationary stochastic process (an input signal); $g_i$ is an impulse response; and $\tilde{c}_i | g_i$ is the output signal evaluated at time $t_i$. Together, this comprises a linear system. Following standard theory[7] , we can express $\text{Var}[\tilde{c}_i | g_i]$ in terms of the autocorrelation $R(\tau)$ of the signal $\tilde{c}_i | g_i$ at time lag $\tau$. Specifically,

$$\begin{aligned}
\text{Var}[\tilde{c}_i | g_i] &= E[(\tilde{c}_i | g_i)^2] - E[\tilde{c}_i | g_i]^2 && (33)\\
&= R_{\tilde{c}_i | g_i}(0) - E[\tilde{c}_i | g_i]^2 && (34)\\
&= R_{\tilde{c}_i | g_i}(0) - 1 && (35)
\end{aligned}$$

**Deriving an expression for $R_{\tilde{c}_i | g_i}(0)$.** Deriving an expression for the second moment of the output process, $R_{\tilde{c}_i | g_i}(0)$, is easiest in frequency space. We can introduce $S_\bullet$ as the Fourier transform of $R_\bullet$ and $\hat{g}_i$ as the Fourier transform of $g_i$, *i.e.*

$$S_\bullet(\omega) = \int_{-\infty}^\infty e^{-i\omega\tau} R_\bullet(\tau) d\tau \qquad \text{and} \qquad \hat{g}_i(\omega) = \int_{-\infty}^\infty e^{-i\omega\tau} g_i(\tau) d\tau \tag{36}$$

By the inverse Fourier transform,

$$R_\bullet(\tau) = \frac{1}{2\pi} \int_{-\infty}^\infty S_\bullet(\omega) e^{i\omega\tau} d\omega \tag{37}$$

and thus

$$R_\bullet(0) = \frac{1}{2\pi} \int_{-\infty}^\infty S_\bullet(\omega) d\omega \tag{38}$$

We thus seek an expression for $S_{\tilde{c}_i | g_i}(\omega)$, which we can obtain from the fundamental theorem for linear systems[7] :

$$S_{\tilde{c}_i | g_i}(\omega) = |\hat{g}_i(\omega)|^2 S_{c_i}(\omega) \tag{39}$$

**Deriving $\hat{g}_i(\omega)$.** We begin by examining $\hat{g}_i(\omega)$. Under the Gamma time-shift model, $g_i(\tau) = f_\epsilon(\tau - l_i)$, where $f_\epsilon$ is the Gamma($\psi\alpha, \beta$) density and $l_i \sim$ Gamma($(1-\psi)\alpha, \beta$). Taking the Fourier transform,

$$\hat{g}_i(\omega) = \int_{-\infty}^{\infty} g_i(\tau) e^{-i\omega\tau} d\tau \tag{40}$$

$$= \int_{l_i}^{\infty} f_\epsilon(\tau - l_i) e^{-i\omega\tau} d\tau \tag{41}$$

$$= \int_{0}^{\infty} f_\epsilon(\tau) e^{-i\omega(\tau + l_i)} d\tau \tag{42}$$

$$= e^{-i\omega l_i} \int_{0}^{\infty} f_\epsilon(\tau) e^{-i\omega\tau} d\tau \tag{43}$$

$$= e^{-i\omega l_i} \int_{0}^{\infty} \frac{\beta^{\psi\alpha}}{\Gamma(\psi\alpha)} \tau^{\psi\alpha - 1} e^{-\beta\tau} e^{-i\omega\tau} d\tau \tag{44}$$

$$= e^{-i\omega l_i} \left(\frac{\beta}{\beta + i\omega}\right)^{\psi\alpha} \tag{45}$$

Taking the squared modulus gives

$$|\hat{g}_i(\omega)|^2 = \left(1 + \frac{\omega^2}{\beta^2}\right)^{-\psi\alpha} \tag{46}$$

This is the power transfer function; it is constant across individuals and does not depend on $l_i$.

**Deriving $S_{c_i}(\omega)$ for the periodic contact model.** For the periodic contact model

$$c_i(t) = 1 - \zeta \cos\left(\frac{2\pi t}{T_c}\right) \tag{47}$$

where $T_c$ is the period of oscillation and $\zeta \in [0, 1]$ is the amplitude, the autocorrelation is

$$R_{c_i}(\tau) = E\left[\left(1 - \zeta \cos\left(\tfrac{2\pi t}{T_c}\right)\right)\left(1 - \zeta \cos\left(\tfrac{2\pi(t+\tau)}{T_c}\right)\right)\right] \tag{48}$$

$$= E[1] - \zeta\, E\left[\cos\left(\tfrac{2\pi t}{T_c}\right)\right] - \zeta\, E\left[\cos\left(\tfrac{2\pi(t+\tau)}{T_c}\right)\right] + \zeta^2\, E\left[\cos\left(\tfrac{2\pi t}{T_c}\right)\cos\left(\tfrac{2\pi(t+\tau)}{T_c}\right)\right] \tag{49}$$

$$= 1 + \frac{\zeta^2}{2}\cos\left(\frac{2\pi\tau}{T_c}\right) \tag{50}$$

Taking the Fourier transform,

$$S_{c_i}(\omega) = \int_{-\infty}^{\infty} \left[1 + \frac{\zeta^2}{2}\cos\left(\tfrac{2\pi\tau}{T_c}\right)\right] e^{-i\omega\tau} d\tau \tag{51}$$

$$= 2\pi\delta(\omega) + \frac{\pi\zeta^2}{2}\left[\delta\left(\omega - \tfrac{2\pi}{T_c}\right) + \delta\left(\omega + \tfrac{2\pi}{T_c}\right)\right] \tag{52}$$

**Constructing $k$ for the periodic contact model.** Putting everything together, we have:

$$\begin{aligned}
\text{Var}[\tilde{c}_i|g_i] &= R_{\tilde{c}_i|g_i}(0) - 1 && (53)\\
&= \frac{1}{2\pi}\int_{-\infty}^{\infty} S_{\tilde{c}_i|g_i}(\omega)d\omega - 1 && (54)\\
&= \frac{1}{2\pi}\int_{-\infty}^{\infty}\left(1+\frac{\omega^2}{\beta^2}\right)^{-\psi\alpha}\left[2\pi\delta(\omega)+\frac{\pi\zeta^2}{2}\left[\delta\left(\omega-\tfrac{2\pi}{T_c}\right)+\delta\left(\omega+\tfrac{2\pi}{T_c}\right)\right]\right]d\omega - 1 && (55)\\
&= 1+\frac{\zeta^2}{2}\left(1+\left(\frac{2\pi}{T_c\beta}\right)^2\right)^{-\psi\alpha} - 1 && (56)\\
&= \frac{\zeta^2}{2}\left(1+\left(\frac{2\pi}{T_c\beta}\right)^2\right)^{-\psi\alpha} && (57)
\end{aligned}$$

Thus,

$$k = \frac{2}{\zeta^2}\left(1+\left(\frac{2\pi\bar{g}}{T_c\alpha}\right)^2\right)^{\psi\alpha} \tag{58}$$

since $\beta = \alpha/\bar{g}$.

**Deriving $S_{c_i}(\omega)$ for the Gamma-Poisson contact model.** Let $X \sim \text{Gamma}(\sigma,\sigma)$ be a contact-level draw from the Gamma-Poisson contact model with switching rate $\lambda$. Then, the autocorrelation

$$R_{c_i}(\tau) = E[c_i(t)c_i(t+\tau)] \tag{59}$$

is equal to $E[X^2] = \text{Var}[X] + E[X]^2 = \frac{1}{\sigma} + 1$ with probability $e^{-\lambda|\tau|}$ (*i.e.*, if the contact levels have not switched between times $t$ and $t+\tau$), and is equal to $E[X]^2 = 1$ otherwise. That is:

$$\begin{aligned}
R_{c_i}(\tau) &= \left(\frac{1}{\sigma}+1\right)e^{-\lambda|\tau|} + (1)(1-e^{-\lambda|\tau|}) && (60)\\
&= \frac{1}{\sigma}e^{-\lambda|\tau|} + 1 && (61)
\end{aligned}$$

Thus:

$$\begin{aligned}
S_{c_i}(\omega) &= \int_{-\infty}^{\infty}\left[\frac{1}{\sigma}e^{-\lambda|\tau|}+1\right]e^{-i\omega\tau}d\tau && (62)\\
&= \frac{1}{\sigma}\int_{-\infty}^{0} e^{\lambda\tau}e^{-i\omega\tau}d\tau + \frac{1}{\sigma}\int_{0}^{\infty} e^{-\lambda\tau}e^{-i\omega\tau}d\tau + 2\pi\delta(\omega) && (63)\\
&= \frac{1}{\sigma}\left[\frac{1}{\lambda-i\omega}+\frac{1}{\lambda+i\omega}\right] + 2\pi\delta(\omega) && (64)\\
&= \frac{2\lambda}{\sigma(\lambda^2+\omega^2)} + 2\pi\delta(\omega) && (65)
\end{aligned}$$

**Constructing $k$ for the Gamma-Poisson contact model.** Putting everything together, we have:

$$\begin{aligned}
\mathrm{Var}[\tilde{c}_i|g_i] &= R_{\tilde{c}_i|g_i}(0) - 1 && (66)\\
&= \frac{1}{2\pi}\int_{-\infty}^{\infty}\Big[\frac{2\lambda}{\sigma(\lambda^2+\omega^2)} + 2\pi\delta(\omega)\Big]\Big(1+\frac{\omega^2}{\beta^2}\Big)^{-\psi\alpha} d\omega - 1 && (67)\\
&= \frac{1}{2\pi}\int_{-\infty}^{\infty}\Big[\frac{2\lambda}{\sigma(\lambda^2+\omega^2)}\Big]\Big(1+\frac{\omega^2}{\beta^2}\Big)^{-\psi\alpha} d\omega && (68)\\
&= \frac{1}{\sigma}\int_{-\infty}^{\infty}\frac{\lambda}{\pi(\lambda^2+\omega^2)}\Big(1+\frac{\omega^2}{\beta^2}\Big)^{-\psi\alpha} d\omega && (69)
\end{aligned}$$

and $k$ is the reciprocal of this integral. The integral can be solved in closed form using special functions, but its closed form does not yield additional insights; in practice, we evaluate this expression by numerically solving the integral.

**Empirical estimation of $k$ as a function of $\psi$ and the contact process.** To validate the closed-form expressions for $k$ derived above, we estimated $k$ by direct simulation of secondary infection counts. For each of the benchmark pathogens, and for finely spaced grids of $\psi$ and $\zeta$ or $\sigma$ (contact amplitude or heterogeneity), we simulated $n = 10{,}000$ index cases, generated each index case's secondary infections under the relevant contact process, and computed the method-of-moments estimate $\hat{k} = \bar{\chi}^2/(\mathrm{Var}[\chi] - \bar{\chi})$ from the sample mean $\bar{\chi}$ and variance $\mathrm{Var}[\chi]$ of the secondary-infection counts (setting $\hat{k} = \infty$ whenever $\mathrm{Var}[\chi] \le \bar{\chi}$). Secondary infection times were generated by Poisson thinning: we first proposed $\chi_{\mathrm{raw}} \sim \mathrm{Poisson}(R_0\, c_{\max})$ candidate transmission attempts with times drawn from the index case's Gamma-burst individual GI distribution, then independently retained each candidate $t_j$ with probability $c(t_j)/c_{\max}$, where $c(t)$ is the contact rate and $c_{\max}$ is the infector's maximum contact rate. The number of retained attempts is the realized secondary-infection count for that index case. For the periodic contact model, each index case was assigned a uniformly-distributed infection time within one period $T_c$, and the contact rate was $c(t) = 1 - \zeta\cos(2\pi t/T_c)$ with $c_{\max} = 1 + \zeta$. For the Gamma-Poisson contact model, each index case was assigned a fresh piecewise-constant contact trajectory whose levels were drawn iid from $\mathrm{Gamma}(\sigma, \sigma)$ at $\mathrm{Exp}(\lambda)$-distributed switching times, with $c_{\max}$ set to the maximum level realized along the trajectory. Heatmaps illustrating these findings with the theoretical prediction curves are depicted in **Fig. 3D–F** and **Extended Data Fig. 4**.

##### 1.9 Burstiness and the epidemic latent period

Like infections, epidemics may have a "latent period": a period of cryptic transmission preceding epidemic establishment. The duration of this latent period depends on early-epidemic stochastic events [19] . Here, we determine how the epidemic latent period's length depends on $\psi$.

**Setup.** Following Morris *et al.* [19], we describe the cumulative infections during the early-epidemic exponential growth phase as a branching process, $Z(t)$. Let $r$ be the epidemic's exponential growth rate; then, as $t \to \infty$, $e^{-rt}Z(t) \to W$ almost surely, where $W$ is a random variable that effectively scales the branching process' initial condition, capturing stochastic fluctuations in the early-epidemic process. We denote this convergence as $Z(t) \approx W \cdot e^{rt}$.

With a change of variables, $W$ can be expressed as a random initial time shift:

$$Z(t) \approx e^{r(t+\varsigma)} \qquad \text{where} \qquad \varsigma = \frac{\log W - \log E[W]}{r} = \frac{\log W}{r} \tag{70}$$

when $I_0 = E[W] = 1$ (*i.e.*, when the epidemic is seeded with a single infectious person). Our goal is to determine how $E[\varsigma]$ and $\mathrm{Var}[\varsigma]$ depend on the burstiness parameter $\psi$. To achieve this, we begin by examining the distribution of $W$.

**Approximating the distribution of $W$.** First, we note that the distribution of $W$ consists of a point mass at 0 with mass $q$ (the probability of epidemic extinction), combined with a smooth density with positive support. The point mass translates into a time shift of $\varsigma = \log(0)/r = -\infty$, corresponding to an epidemic that never establishes. Thus, we restrict our attention to $W|\text{surv}$ and $\varsigma|\text{surv}$, that is, $W$ and $\varsigma$ conditioned on epidemic survival.

To approximate the distribution of $W|\text{surv}$, we use a $\text{Gamma}(k, \lambda)$ distribution matched to the first two moments. This is a version of the five-moment-matching approach described by Morris *et al.* [19], simplified for analytic tractability. Specifically, we let

$$k = \frac{E[W|\text{surv}]^2}{\text{Var}[W|\text{surv}]} \qquad \text{and} \qquad \lambda = \frac{E[W|\text{surv}]}{\text{Var}[W|\text{surv}]} \tag{71}$$

(Note: these $k$ and $\lambda$ are distinct from the overdispersion parameter and the Gamma-Poisson switching rate defined above; their scope is strictly for this subsection). Our task is now to compute $E[W|\text{surv}]$ and $\text{Var}[W|\text{surv}]$.

**Deriving $E[W|\text{surv}]$.** We note that

$$E[W] = E[W|\text{surv}]P(\text{surv}) + E[W|\text{extinct}]P(\text{extinct}) = E[W|\text{surv}] \cdot p \tag{72}$$

since $E[W|\text{extinct}] = 0$, and where $p = 1 - q$ is the probability of epidemic survival. Thus,

$$E[W|\text{surv}] = \frac{1}{p} \tag{73}$$

under our assumption that $E[W] = 1$.

**Deriving $\text{Var}[W|\text{surv}]$.** The derivation for $\text{Var}[W|\text{surv}]$ is somewhat more involved. Recall the Gamma time-shift model, where an infectious person produces $\chi_i \sim \text{Poisson}(R_0)$ secondary infections distributed at times $\tau_j = l_i + \epsilon_j,\ j \in \{1, \ldots, \chi_i\}$ and

$$l_i \sim \text{Gamma}((1-\psi)\alpha, \beta), \qquad \epsilon_j \sim \text{Gamma}(\psi\alpha, \beta) \tag{74}$$

This ensures that a random secondary infection time $\tau$ is marginally distributed according to the generation interval distribution $g(\tau)$, *i.e.*,

$$\tau \sim \text{Gamma}(\alpha, \beta) \tag{75}$$

For notational convenience, we introduce

$$E[e^{-cr\tau}] = \int_0^\infty e^{-cr\tau} \frac{\beta^\alpha}{\Gamma(\alpha)} \tau^{\alpha-1} e^{-\beta\tau}\, d\tau = \left(\frac{1}{1+cz}\right)^\alpha := \rho_c^\alpha \tag{76}$$

that is,

$$\rho_c := \frac{1}{1+cz} \tag{77}$$

where $z = r/\beta = r\bar{g}/\alpha = R_0^{1/\alpha} - 1$, a dimensionless quantity that simplifies expressions and allows one to easily plug in different combinations of epidemiological parameters. Here, $\bar{g}$ is the mean of the generation interval distribution $g(\tau)$, and $c$ is a positive integer. The final expression, $z = R_0^{1/\alpha} - 1$, comes from the Euler-Lotka equation [1] :

$$1 = R_0 E[e^{-r\tau}] = R_0 \rho_1^\alpha = R_0 \left(\frac{1}{1+z}\right)^\alpha \tag{78}$$

and solving for $z$.

Next, we introduce the distributional fixed point equation, which is a standard starting point for computing moments of $W$. This equation exploits the self-similarity of the branching process, in which an epidemic initiated by a single individual is, in distribution, a scaled and time-shifted copy of the whole epidemic. At large $t$, the epidemic seeded by the index case $i$ consists of the downstream epidemics caused by their offspring $j$, which have grown approximately to size $Z_j(t) = W_j e^{r(t-\tau_j)}$. That is,

$$Z(t) \approx \sum_{j=1}^{\chi_i} W_j e^{rt} e^{-r\tau_j}$$

Multiplying through by $e^{-rt}$:

$$W \stackrel{d}{=} \sum_{j=1}^{\chi_i} e^{-r\tau_j} W_j$$

We now use this expression to calculate $\text{Var}[W]$. Since randomness enters through both $W$ and $\tau$, we first condition on the number of offspring and their infection times, $\tau_\bullet = \{\chi_i, \tau_1, \tau_2, \ldots, \tau_{\chi_i}\}$, using the law of total variance:

$$\text{Var}[W] = E[\text{Var}\Big[\sum_j e^{-r\tau_j} W_j | \tau_\bullet\Big]] + \text{Var}[E\Big[\sum_j e^{-r\tau_j} W_j | \tau_\bullet\Big]]$$

$$= E\Big[\sum_j e^{-2r\tau_j} \cdot \text{Var}[W]\Big] + \text{Var}\Big[\sum_j e^{-r\tau_j} \cdot E[W]\Big]$$

$$= E\Big[\sum_j e^{-2r\tau_j}\Big] \cdot \text{Var}[W] + (E[W])^2 \cdot \text{Var}\Big[\sum_j e^{-r\tau_j}\Big]$$

Since $E[W] = I_0 = 1$, we have

$$\text{Var}[W] = \text{Var}[W] E\Big[\sum_j e^{-2r\tau_j}\Big] + \text{Var}\Big[\sum_j e^{-r\tau_j}\Big]$$

Solving for $\text{Var}[W]$,

$$\text{Var}[W] = \frac{\text{Var}[\sum_j e^{-r\tau_j}]}{1 - E[\sum_j e^{-2r\tau_j}]}$$

To simplify the denominator, note that

$$E\Big[\sum_{j=1}^{\chi_i} e^{-2r\tau_j}\Big] = E[\chi_i] E[e^{-2r\tau_j}] = R_0 E[e^{-2r\tau_j}] = R_0 \rho_2^\alpha$$

Thus, the variance expression simplifies to

$$\text{Var}[W] = \frac{\text{Var}[\sum_j e^{-r\tau_j}]}{1 - R_0 \rho_2^\alpha}$$

Next, we examine the numerator:

$$\text{Var}[\sum_j e^{-r\tau_j}] = E[(\sum_j e^{-r\tau_j})^2] - E[\sum_j e^{-r\tau_j}]^2 = E[(\sum_j e^{-r\tau_j})^2] - 1$$

where the last step follows from the Euler-Lotka equation (Eq. 78). Then,

$$E[(\sum_j e^{-r\tau_j})^2] = E[\sum_j e^{-2r\tau_j}] + E[\sum_{j \neq k} e^{-r(\tau_j + \tau_k)}]$$

$$= R_0 \rho_2^\alpha + E[\chi_i(\chi_i - 1)] E[e^{-r\tau_j} e^{-r\tau_k}]$$

$$= R_0 \rho_2^\alpha + R_0^2 E[e^{-r\tau_j} e^{-r\tau_k}]$$

Here, the $\chi_i(\chi_i - 1)$ term enters because there are this many terms in the sum over $j \neq k$. Also,

$$E[\chi_i(\chi_i - 1)] = E[\chi_i^2] - E[\chi_i] = \text{Var}[\chi_i] + E[\chi_i]^2 - E[\chi_i] = R_0 + R_0^2 - R_0 = R_0^2$$

under the assumption that $\chi_i \sim \text{Poisson}(R_0)$.

Since $\tau_j = l_i + \epsilon_j$, and since $l_i$, $\epsilon_j$, and $\epsilon_k$ are independent,

$$E[e^{-r\tau_j}e^{-r\tau_k}] = E[e^{-2rl_i}e^{-r\epsilon_j}e^{-r\epsilon_k}] = E[e^{-2rl_i}](E[e^{-r\epsilon}])^2 = \rho_2^{(1-\psi)\alpha} \cdot \rho_1^{2\psi\alpha}$$

Finally, we can assemble $\text{Var}[W]$:

$$\text{Var}[\sum_j e^{-r\tau_j}] = R_0\rho_2^\alpha + R_0^2\rho_2^{(1-\psi)\alpha}\rho_1^{2\psi\alpha} - 1$$

and thus, substituting in the $z$ expressions for $\rho_1$ and $\rho_2$ and noting $R_0 = (1+z)^\alpha$, we obtain

$$\text{Var}[W] = \frac{\left(\frac{1+z}{1+2z}\right)^\alpha + \left(\frac{(1+z)^2}{1+2z}\right)^{(1-\psi)\alpha} - 1}{1 - \left(\frac{1+z}{1+2z}\right)^\alpha} := \frac{\phi_1^\alpha + \phi_2^{(1-\psi)\alpha} - 1}{1 - \phi_1^\alpha} \tag{79}$$

where $\phi_c = \frac{(1+z)^c}{1+2z}$ for positive integer $c$.

Last, we note that $E[W^n|\text{surv}] = E[W^n]/p$ (by the same logic as in Eq. 72), so

$$\text{Var}[W|\text{surv}] = E[W^2|\text{surv}] - E[W|\text{surv}]^2 = \frac{1}{p}E[W^2] - \frac{1}{p^2}E[W]^2 \tag{80}$$

$$= \frac{1}{p}(1 + \text{Var}[W]) - \frac{1}{p^2} \tag{81}$$

**Expressions for $k$ and $\lambda$.** Combining the previous derivations and plugging into Eq. 71 gives

$$k = \frac{1 - \phi_1^\alpha}{p\phi_2^{(1-\psi)\alpha} - 1 + \phi_1^\alpha}, \qquad \lambda = pk \tag{82}$$

These are the parameters of the moment-matched Gamma distribution for $W|\text{surv}$.

**Deriving the approximate density of $\varsigma$.** Let $f_W(x)$ be the Gamma-distributed approximate density of $W|\text{surv}$:

$$f_W(x) = \frac{\lambda^k}{\Gamma(k)}x^{k-1}e^{-\lambda x} \tag{83}$$

Since $\varsigma = \frac{1}{r}\log W$, we have $W = e^{r\varsigma}$, or, in terms of the arguments of the density functions, $x = e^{rs}$. By the change of variables formula, the density of $\varsigma$ is

$$f_\varsigma(s) = f_W(e^{rs})|\frac{dw}{ds}| = \frac{\lambda^k}{\Gamma(k)}e^{rs(k-1)}e^{-\lambda e^{rs}}re^{rs} = \frac{r\lambda^k}{\Gamma(k)}e^{rsk}e^{-\lambda e^{rs}} \tag{84}$$

**Deriving the mode of $\varsigma$.** By taking the logarithm, differentiating with respect to $s$, and setting equal to 0, we can determine the mode of the distribution:

$$\text{Mode}[\varsigma] = \frac{1}{r}\log\left(\frac{k}{\lambda}\right) = \frac{1}{r}\log\frac{1}{p} \tag{85}$$

Thus, the modal time shift depends on the epidemic growth rate $r$ and the establishment probability $p$ (and thus the reproduction number $R_0$), but not on the burstiness parameter $\psi$. This derivation of the mode will be helpful for assessing how the skewness of $W|\text{surv}$ depends on $\psi$.

**Deriving $E[\varsigma]$.** It would be possible to derive $E[\varsigma]$ directly from Eq.84, but there is a more direct route: since $W \sim \text{Gamma}(k, \lambda)$ and $\varsigma = \frac{1}{r}\log(W)$, it follows that

$$E[\varsigma] = \frac{1}{r}(\Psi^{(0)}(k) - \log\lambda) \tag{86}$$

where $\Psi^{(0)}(k)$ is the digamma function evaluated at $k$. Plugging in $\lambda = pk$:

$$E[\varsigma] = \frac{\log(1/p)}{r} + \frac{\Psi^{(0)}(k) - \log(k)}{r} \tag{87}$$

The first term is $\text{Mode}[\varsigma]$ (Eq. 85), and is independent of the burstiness parameter $\psi$. The second term is a $\psi$-dependent adjustment that is:

1. **Strictly negative.** *Proof:* Let $G \sim \text{Gamma}(k, 1)$. Then, $E[G] = k$ and $E[\log G] = \Psi^{(0)}(k)$. By Jensen's inequality, since log is strictly concave,

$$\Psi^{(0)}(k) = E[\log G] < \log(E[G]) = \log k$$

   Thus, $\left(\Psi^{(0)}(k) - \log(k)\right)/r < 0$.

2. **Strictly increasing (less negative) in $\psi$.** *Proof:* Consider

$$\frac{d}{dk}\left[\Psi^{(0)}(k) - \log k\right] = \frac{d}{dk}\left[\int_0^\infty \left(\frac{e^{-t}}{t} - \frac{e^{-kt}}{1-e^{-t}}\right)dt\right] - \frac{1}{k} = \int_0^\infty \frac{te^{-kt}}{1-e^{-t}}\,dt - \frac{1}{k}$$

   Note that $1 - e^{-t} < t$ for $t > 0$, and thus $\frac{t}{1-e^{-t}} > 1$. So,

$$\int_0^\infty \frac{te^{-kt}}{1-e^{-t}} > \int_0^\infty e^{-kt}\,dt = \frac{1}{k}$$

   So,

$$\frac{d}{dk}\left[\Psi^{(0)}(k) - \log k\right] > 0$$

   and thus $(\Psi^{(0)}(k) - \log k)/r$ is increasing in $k$.
   Next, we note that $k$ is increasing in $\psi$. The only $\psi$-dependent quantity in $k$ (Eq.71) is $\phi_2^{(1-\psi)\alpha}$, which shows up in the denominator. Since

$$\phi_2 = \frac{(1+z)^2}{(1+2z)} = 1 + \frac{z^2}{1+2z} > 1 \quad \text{for } z > 0$$

   the term is maximized when $\psi = 0$ and minimized when $\psi = 1$. Thus, $k$ is minimized when $\psi = 0$ and maximized when $\psi = 1$ — *i.e.*, $k$ is increasing in $\psi$.

Together, these observations demonstrate that the expected epidemic onset time sits later than the modal onset (*i.e.*, the time shift distribution is left-skewed; 1), and the expected time shift moves increasingly later as $\psi$ decreases (2).

**Deriving Var[$\varsigma$].** Following again from the properties of $\log W$ where $W$ is Gamma$(k, \lambda)$-distributed,

$$\text{Var}[\varsigma] = \frac{1}{r^2}\Psi^{(1)}(k) \tag{88}$$

where $\Psi^{(1)}(k)$ is the trigamma function. The trigamma function is strictly decreasing in $k$, since

$$\frac{d}{dk}\Psi^{(1)}(k) = \frac{d}{dk}\sum_{n=0}^{\infty}\frac{1}{(k+n)^2} = -2\sum_{n=0}^{\infty}\frac{1}{(k+n)^3} \tag{89}$$

which is strictly negative when $k > 0$. Thus, bursty individual GI distributions ($\psi \to 0$) yield decreasing $k$, which yields increasing Var[$\varsigma$].

**Summary of the derivation.** Using a branching process approximation to early-epidemic growth, we derived expressions for the mean and variance of $W|\text{surv}$, which is a random initial condition that accounts for early-epidemic stochasticity (conditioning on epidemic survival). It turns out that Var[$W|\text{surv}$] depends on $\psi$: bursty infectiousness ($\psi \to 0$) yields higher Var[$W|\text{surv}$]. Then, using $E[W|\text{surv}]$ and Var[$W|\text{surv}$], we generated a moment-matched Gamma approximation for the distribution of $W|\text{surv}$. From this, we derived an approximate density for the time shift random variable, $\varsigma$. We found that Mode[$\varsigma$] is independent of $\psi$, but $E[\varsigma] <$ Mode[$\varsigma$] (*i.e.*, the distribution of $\varsigma$ is left-skewed). Furthermore, $E[\varsigma]$ is increasing in $\psi$, so bursty infectiousness ($\psi \to 0$) yields smaller $E[\varsigma]$, which translates into later-shifted epidemics on average. The derivation of $E[\varsigma]$ was simplified by noting that the mean of a log-Gamma-distributed random variable is given by the digamma function. Finally, we relied on the fact that the variance of a log-Gamma-distributed random variable is given by the trigamma function to show that bursty individual GI distributions increase Var[$\varsigma$].

**Empirical description of the epidemic latent period.** To validate the theoretical $\varsigma$ distribution derived above and to visualize how $\psi$ shapes the spread of epidemic establishment times, we ran stochastic individual-based simulations and compared the resulting establishment time distributions to the theoretical predictions. For each benchmark pathogen and each of $\psi \in \{0, 0.5, 1\}$, we generated 5,000 simulated outbreaks in a population of $N = 10,000$. Each outbreak was seeded with a single index case at $t = 0$, and we recorded the establishment time as the time when cumulative infections first reached 500 cases (5% of the population). The empirical survival curve $\hat{S}(t)$ for a given $\psi$ is the fraction of simulated epidemics that have not yet reached 500 cases by day $t$.

To compare against theory, we computed the time $t_{\text{det}}$ at which the deterministic cumulative incidence curve, generated using the renewal equation simulation model, crosses the establishment threshold (500 cases). We then drew values of $W$ from the moment-matched Gamma approximation to $W|\text{surv}$ derived above. Each draw was mapped to a stochastic establishment time via the time-shift relation $t_{\text{stoch}} = t_{\text{det}} - \log(W)/r$, and the theoretical survival curve was constructed as the empirical CDF of these $t_{\text{stoch}}$ values. The empirical and theoretical survival curves for each $\psi$ are overlaid in **Extended Data Fig. 5D–F**. The empirical and theoretical distributions of $\varsigma$ are depicted in **Extended Data Fig. 5A–C**.

##### 1.10 Detect-and-isolate interventions with bursty infectiousness

Detect-and-isolate (D&I) interventions are critical non-pharmaceutical interventions, aimed at identifying individuals at an early stage of infection to prevent onward spread. Detection can happen *via* the development of symptoms and/or diagnostic testing; thus, the timing of symptom onset and the window of test-based detectability, relative to the window of infectiousness, are critical factors influencing the success of D&I interventions[20] .

The efficacy of D&I interventions can be summarized in terms of testing effectiveness (TE), defined as the expected proportion by which a D&I intervention reduces population-level transmission[21] . Formally,

$$\mathrm{TE} = \rho\eta P(\tau > D) \tag{90}$$

where $\rho \in [0, 1]$ is the adherence rate, $\eta \in [0, 1]$ is the effectiveness of isolation (so that post-isolation transmission attempts succeed with probability $1 - \eta$), $\tau$ is the time of a transmission attempt relative to the index case's infection time, and $D$ is the isolation time relative to infection onset.

When the isolation time is anchored to the index case's infection time (*e.g.*, the detectability window spans 2–7 days after infection), TE depends only on the shape of the population-level GI distribution; the shape of the individual GI distribution integrates out. (Justification: under the Gamma burst model, $\tau = l_i + \epsilon_j \sim \mathrm{Gamma}(\alpha, \beta)$ marginally, regardless of $\psi$. When $D$ is anchored to the infection time, $P(\tau > D) = P(\mathrm{Gamma}(\alpha, \beta) > D)$, which depends only on $\alpha$ and $\beta$.) If, however, $D$ is anchored to the individual GI distribution (*e.g.*, the window of detectability extends ± 3 days from peak infectiousness), TE depends on the width of the individual GI distribution, as we illustrate here. Anchoring $D$ to the individual GI distribution is arguably the more realistic scenario since symptom onset, test-based detectability, and peak pathogen shedding are all linked to viral load, which in turn shapes the individual's infectiousness timing[22] .

As illustrations, we considered two detection mechanisms: symptom-based isolation, where symptoms arise at a $\mathrm{Normal}(\delta_{\mathrm{symp}}, \sigma^2_{\mathrm{symp}})$-distributed offset from the peak infectiousness and isolation is immediate; and test-based screening, where diagnostic tests are administered at fixed intervals of length $\Delta_{\mathrm{test}}$, and the first test that falls within a detectability window spanning $\delta_{\mathrm{pre}}$ through $\delta_{\mathrm{post}}$ days before/after peak infectiousness triggers isolation after a $\mathrm{Exp}(\lambda_{\mathrm{act}})$-distributed turnaround delay. In both cases, the computation of $P(\tau > D)$ simplifies because the shared latent shift $l_i$ cancels: since $\tau = l_i + \epsilon_j$ and the detection/isolation time is anchored to the individual's peak infectiousness at $l_i + m_\epsilon$ (where $m_\epsilon$ is the mode of the $\mathrm{Gamma}(\psi\alpha, \beta)$ distribution), $P(\tau > D) = P(\epsilon_j > m_\epsilon + \iota)$, where $\iota$ is the isolation time's offset relative to $m_\epsilon$, and $\epsilon_j$ is a draw from $f_\epsilon = \mathrm{Gamma}(\psi\alpha, \beta)$. TE thus reduces to the survival function of $f_\epsilon$ evaluated at $m_\epsilon + \iota$, marginalized over the distribution of $\iota$ induced by the detection/isolation protocol.

For symptom-based isolation, the isolation offset is $\iota \sim \mathrm{Normal}(\delta_{\mathrm{symp}}, \sigma^2_{\mathrm{symp}})$, so

$$\mathrm{TE} = \rho\,\eta \int_{-\infty}^{\infty} \left[1 - F_\epsilon(m_\epsilon + t)\right] f_{\mathrm{Normal}}(t;\, \delta_{\mathrm{symp}}, \sigma^2_{\mathrm{symp}})\, dt \tag{91}$$

where $F_\epsilon$ is the CDF of $f_\epsilon$ and $f_{\mathrm{Normal}}$ is the Normal density. For test-based screening, regularly-spaced tests at interval $\Delta_{\mathrm{test}}$ fall within a detectability window of length $w = \delta_{\mathrm{pre}} + \delta_{\mathrm{post}}$ spanning from $\delta_{\mathrm{pre}}$ days before peak to $\delta_{\mathrm{post}}$ days after, and the first test that falls in the window triggers isolation (we assume perfect sensitivity/specificity). When a test does land in the window, its offset from the window's start is $u \sim \mathrm{Uniform}(0, \min(\Delta_{\mathrm{test}}, w))$. When $\Delta_{\mathrm{test}} > w$, it is possible for no test to land in the window; this happens with probability $(\Delta_{\mathrm{test}} - w)/\Delta_{\mathrm{test}}$. When isolation follows immediately upon a positive test, the isolation offset is $\iota = u - \delta_{\mathrm{pre}}$ and

$$\mathrm{TE} = \frac{\rho\,\eta}{\Delta_{\mathrm{test}}} \int_0^{\min(\Delta_{\mathrm{test}}, w)} \left[1 - F_\epsilon(m_\epsilon + u - \delta_{\mathrm{pre}})\right] du. \tag{92}$$

When isolation instead follows a turnaround delay $v \sim \mathrm{Exp}(\lambda_{\mathrm{act}})$, the isolation offset becomes $\iota = u - \delta_{\mathrm{pre}} + v$. Marginalizing over $v$ gives

$$\mathrm{TE} = \frac{\rho\,\eta}{\Delta_{\mathrm{test}}} \int_0^{\min(\Delta_{\mathrm{test}}, w)} \int_0^{\infty} \left[1 - F_\epsilon(m_\epsilon + u - \delta_{\mathrm{pre}} + v)\right] f_{\mathrm{Exp}}(v;\, \lambda_{\mathrm{act}})\, dv\, du \tag{93}$$

where $f_{\mathrm{Exp}}$ is the Exponential density. These integrals can be solved numerically to illustrate how TE varies with features of the detection/isolation mechanism (**Extended Data Fig. 6A–F**).

Beyond TE, mode-anchored D&I interventions produce two additional $\psi$-dependent effects: generation interval truncation, which can accelerate epidemic growth; and increased overdispersion in secondary infection counts, which decreases the probability of epidemic survival. To build intuition for the truncation, we can consider the simplest D&I protocol: a fixed (deterministic) mode-anchored isolation offset $\iota$. A transmission attempt that occurs at time-after-latent-period $\epsilon < m_\epsilon + \iota$, where $\epsilon \sim \text{Gamma}(\psi\alpha, \beta)$, occurs before isolation and is unaffected; an attempt with $\epsilon \geq m_\epsilon + \iota$ occurs after isolation and is suppressed with probability $\rho\eta$ (the index case must both participate, with probability $\rho$, and the isolation must successfully prevent the attempt, with probability $\eta$). Thus, the generation interval distribution under D&I intervention is

$$g^*(\tau) = \frac{(1 - \rho\eta)\, g(\tau) + \rho\eta \int_0^{\min(\tau,\, m_\epsilon + \iota)} f_l(\tau - \epsilon)\, f_\epsilon(\epsilon)\, d\epsilon}{1 - \text{TE}} \tag{94}$$

The first term represents unaffected transmissions, arising because a person doesn't participate ($\rho$) or because isolation is imperfect ($\eta$); these contribute the full $g(\tau)$. The second term captures the modification due to D&I intervention: infection attempts occur at time $\tau = l + \epsilon$, and thus the probability density of $\tau$ is the convolution $f_l * f_\epsilon$. Under no intervention, this would be $\int_0^\tau f_l(\tau - \epsilon) f_\epsilon(\epsilon)\, d\epsilon$ (where the bounds arise because $f_\epsilon(\epsilon) = 0$ when $\epsilon < 0$ and because $f_l(\tau - \epsilon) = 0$ when $\epsilon > \tau$). However, under D&I intervention, $f_\epsilon(\epsilon) = 0$ also when $\epsilon > m_\epsilon + \iota$, giving the upper bound of the integral in Eq. 94. We can gain some intuition by considering the extremes of $\psi$: in the sustained extreme ($\psi = 1$), $f_l$ is a point mass at 0 and $f_\epsilon = g$; thus, the integral becomes

$$\int_0^{\min(\tau, m_\epsilon + \iota)} \delta(\tau - \epsilon) g(\epsilon)\, d\epsilon = \begin{cases} g(\tau) & \tau \leq m_\epsilon + \iota \\ 0 & \text{otherwise} \end{cases} \tag{95}$$

With complete adherence and perfect isolation ($\rho = \eta = 1$), $g^*(\tau)$ becomes a scaled version of $g(\tau)$ with the tail fully cut off. Alternatively, at the spike extreme ($\psi = 0$), $f_l = g$ and $f_\epsilon$ is a point mass at 0, so the integral becomes

$$\int_0^{\min(\tau, m_\epsilon + \iota)} g(\tau - \epsilon) \delta(\epsilon)\, d\epsilon = g(\tau) \tag{96}$$

That is, $g^*(\tau) = g(\tau)$: the generation interval distribution is unaffected by intervention. A simulation-based illustration of the shifted GI distribution under symptom-based detection is provided in **Figure 5D**.

This GI distortion feeds directly into the post-intervention exponential growth rate $r^*$. Continuing with deterministic isolation time $\iota$, and with $\rho = \eta = 1$, we can derive the epidemic growth rate using the Euler-Lotka equation:

$$1 = (1 - \mathrm{TE})\, R_0 \int_0^\infty e^{-r^*\tau}\, g^*(\tau)\, d\tau \tag{97}$$

$$= R_0 \int_0^\infty e^{-r^*\tau} \int_0^{\min(\tau,\, m_\epsilon+\iota)} f_l(\tau-\epsilon)\, f_\epsilon(\epsilon)\, d\epsilon\, d\tau \tag{98}$$

$$= R_0 \int_0^{m_\epsilon+\iota} f_\epsilon(\epsilon) \int_0^\infty e^{-r^*\tau}\, f_l(\tau-\epsilon)\, d\tau\, d\epsilon \tag{99}$$

$$= R_0 \int_0^{m_\epsilon+\iota} f_\epsilon(\epsilon)\, e^{-r^*\epsilon} \left(\frac{\beta}{\beta+r^*}\right)^{(1-\psi)\alpha} d\epsilon \tag{100}$$

$$= R_0 \left(\frac{\beta}{\beta+r^*}\right)^{(1-\psi)\alpha} \int_0^{m_\epsilon+\iota} e^{-r^*\epsilon}\, f_\epsilon(\epsilon)\, d\epsilon \tag{101}$$

$$= R_0 \left(\frac{\beta}{\beta+r^*}\right)^{\alpha} F_{\mathrm{Gamma}}(m_\epsilon + \iota;\, \psi\alpha,\, \beta + r^*), \tag{102}$$

The growth rate $r^*$ can then be solved for numerically as a function of $\psi$, as illustrated in **Extended Data Fig. 6G–I** for deterministic $\iota = 2$ days and for the three benchmark pathogens. D&I interventions reduce the epidemic growth rate ($r^* \leq r$), but by less than would be expected by considering TE alone. The gap between the actual D&I-reduced and "naive" (considering only TE) growth rate is largest for sustained infectiousness ($\psi = 1$): for example, with $\psi = 1$, the SARS-CoV-2 omicron growth rate under D&I intervention is 0.34/day, *vs.* a predicted growth rate of 0.20/day when considering TE alone. Thus, the GI truncation effect can substantially offset the TE-induced reduction in epidemic growth rates at the sustained infectiousness limit, meaning that D&I interventions targeting pathogens with sustained individual GI distributions could meaningfully reduce $R_0$ but slow epidemic growth only modestly.

Additionally, under mode-anchored D&I interventions, the individual reproduction number becomes

$$\nu_i^* = \nu_i \big[1 - \rho\eta\, [1 - F_\epsilon(m_\epsilon + \iota_i)]\big], \tag{103}$$

where $\nu_i$ is the individual reproduction number in the absence of intervention and $\iota_i$ is the mode-anchored detection offset for person $i$. To justify this expression: in the absence of intervention, person $i$ makes $\nu_i$ transmission attempts at offsets $\epsilon_{ij} \sim f_\epsilon$ from the start of their burst. Infection attempt $j$ is at risk of suppression if $\epsilon_{ij} > m_\epsilon + \iota_i$, which happens with probability $1 - F_\epsilon(m_\epsilon + \iota_i)$. These at-risk infection attempts are suppressed with probability $\rho\eta$ (adherence × effectiveness), so the expected fraction of infection events that are suppressed is $\rho\eta[1 - F_\epsilon(m_\epsilon + \iota_i)]$. The expected number that succeed, then, is given by Eq. 103. Variability in $\iota_i$ creates additional variability in $\nu_i^*$ relative to $\nu_i$, *i.e.*, more overdispersion. The amount of overdispersion depends on how sensitively $F_\epsilon(m_\epsilon + \iota_i)$ changes with shifts in $\iota_i$. For example, under symptom-based detection, if symptoms tend to arise near the peak of infectiousness ($E[\iota_i] \approx 0$), bursty infectiousness ($\psi \to 0$) can cause small variations in symptom onset time to translate into large variations in $F_\epsilon$ (due to the sharp drop depicted in **Extended Data Fig. 6A–C**), creating substantial overdispersion. For smooth infectiousness ($\psi \to 1$), $F_\epsilon$ varies less sharply near $\iota = 0$, so that variation in $\iota_i$ does not produce as much additional variation in $\nu_i^*$. This is illustrated using simulations for symptomatic screening in **Figure 5E**. In sum, D&I interventions produce additional overdispersion when isolation times are random, and this effect can be accentuated when infectiousness is bursty.

##### 1.11 Gathering size restrictions with bursty infectiousness

Gathering size restrictions are another important class of non-pharmaceutical intervention, aimed at limiting opportunities for major transmission events. Gathering size restrictions truncate the tail of the interpersonal contact distribution, reducing the mean contact rate and thus the effective reproduction number. This truncation also reduces overdispersion in secondary infection counts in a $\psi$-dependent way, where the absolute drop in overdispersion is greatest when the original, uncontrolled process was most overdispersed.

To demonstrate this, we considered a modified Gamma-Poisson stochastic contact process: as before, the contact level $c_i(t)$ for person $i$ switches at $\text{Exp}(\lambda)$-distributed times, but the new levels are drawn from a truncated $\text{Gamma}(\sigma, \sigma)$ distribution, rather than from the full distribution. The truncation threshold, $c_{\max}$, captures the severity of the restriction.

Our goal is to describe the overdispersion of secondary infection counts under gathering size restrictions *vs.* without restrictions. The first step is to denote the individual reproduction number, $\nu_i$, as a function of the restricted and unrestricted contact processes. We assume everyone has the same biological infectiousness $b_i = R_0$. Thus,

$$\nu_i = R_0 \int_0^\infty g_i(\tau) c_i(t_i + \tau) d\tau = R_0 \tilde{c}_i \qquad \text{and} \qquad \nu_i^* = R_0 \int_0^\infty g_i(\tau) c_i^*(t_i + \tau) d\tau = R_0 \tilde{c}_i^* \tag{104}$$

for the unrestricted and restricted contact processes $c_i(t)$ and $c_i^*(t)$, respectively, following Eq. 6. For the unrestricted contact process, we have $E[c_i(t)] = 1$ and $\text{Var}[c_i(t)] = 1/\sigma$. For the restricted contact process, we denote the mean and variance $E[c_i^*(t)] = \mu_T$ and $\text{Var}[c_i^*(t)] = V_T$, with $\mu_T < 1$ and $V_T < 1/\sigma$.

Next, we can compute the overdispersion of secondary case counts for each contact process. Recalling Eq. 26, we have

$$\text{OD} := \frac{1}{k} = \frac{\text{Var}[\nu_i]}{E[\nu_i]^2} = \text{Var}[\tilde{c}_i] \qquad \text{and} \qquad \text{OD}^* = \frac{1}{k^*} = \frac{\text{Var}[\nu_i^*]}{E[\nu_i^*]^2} = \frac{\text{Var}[\tilde{c}_i^*]}{\mu_T^2} \tag{105}$$

as the overdispersion under the uncontrolled and controlled contact processes, respectively.

We have already calculated $\text{Var}[\tilde{c}_i]$ in Eq. 69. More generally, following the same derivation but keeping general $\text{Var}[X]$ and $E[X]$ (where $X$ is a random variable denoting the contact-level draws), we see that

$$\text{Var}[\tilde{c}_i] = \text{Var}[X] \int_{-\infty}^{\infty} \frac{\lambda}{\pi(\lambda^2 + \omega^2)} \left(1 + \frac{\omega^2}{\beta^2}\right)^{-\psi\alpha} d\omega := \text{Var}[X] h(\lambda, \alpha, \beta, \psi) \tag{106}$$

defining $h$ as the integral expression. So, $\text{Var}[\tilde{c}_i]$ (or $\text{Var}[\tilde{c}_i^*]$) decomposes into the variance of the contact-level distribution multiplied by an integral equation that depends on the contact switching rate $\lambda$, the generation interval distribution parameters $\alpha$ and $\beta$, and on $\psi$, but not on the contact-level distribution.

With this, we can calculate the relative and absolute difference in overdispersion generated by gathering size restrictions, relative to the unrestricted scenario. The relative difference is

$$\Delta\text{OD}_{\text{rel}} = \frac{\text{OD}^*}{\text{OD}} = \frac{\text{Var}[\tilde{c}_i^*]/\mu_T^2}{\text{Var}[\tilde{c}_i]} = \frac{V_T\ h(\lambda, \alpha, \beta, \psi)/\mu_T^2}{(1/\sigma)\ h(\lambda, \alpha, \beta, \psi)} = \frac{\sigma V_T}{\mu_T^2} \tag{107}$$

and the absolute difference is

$$\Delta\text{OD}_{\text{abs}} = \text{OD}^* - \text{OD} = \text{OD}\left[\frac{\text{OD}^*}{\text{OD}}\right] - \text{OD} = \text{OD}\left[\frac{\sigma V_T}{\mu_T^2} - 1\right] \tag{108}$$

The relative difference in OD is independent of $\psi$: it depends only on the means and variances of the contact processes, but not on the distribution of infectiousness. The absolute difference in OD depends on $\psi$ insofar as the overdispersion in the unrestricted scenario, OD, depends on $\psi$. In Sec. 1.8, we found that bursty infectiousness ($\psi \to 0$) increases overdispersion when contacts vary over time. Thus, bursty infectiousness causes larger OD, and thus larger absolute reduction in OD, under gathering size restrictions. Intuitively, when the unrestricted OD is small (which correlates with broad infectiousness, $\psi \to 1$), the additional reduction in overdispersion caused by gathering size restrictions has little absolute impact. When the unrestricted overdispersion is large (which correlates with bursty infectiousness, $\psi \to 0$), the reduction in overdispersion caused by gathering size restrictions can be consequential.

Simulations illustrate this principle further. We considered an unrestricted Gamma-Poisson stochastic contact process with $\sigma = \lambda = 1$. We then considered three comparable epidemic scenarios: restricted gathering sizes with $c_{\max} = 2$ (limiting contact rates to twice the baseline); unrestricted gathering sizes but with $R_0$ matched to the gathering-size-restricted scenario; and constant (time-invariant) contact rates with $R_0$ matched to the gathering-size-restricted scenario. We computed the overdispersion of secondary infection counts directly from simulated index cases. For each of the three benchmark pathogens and for $\psi \in \{0, 0.1, 0.25, 0.4, 0.5, 0.6, 0.75, 0.9, 1\}$, we simulated 10,000 index cases under each of the contact process scenarios. We computed the method-of-moments estimator $\widehat{\text{OD}} = (\widehat{\text{Var}}[\chi_i] - \widehat{E}[\chi_i])/\widehat{E}[\chi_i]^2)$ from the offspring counts $\chi_i$ for each index case $i$. The resulting $\widehat{\text{OD}}$ estimates closely track the analytic predictions (Eq. 105–106; **Extended Data Fig. 7A–C**). As $\psi$ increases, OD for the variable contact scenarios decreases, and the absolute gap between OD values for the "Restricted" and "Unrestricted" contact scenarios shrinks. This can be illustrated further by plotting $\Delta\text{OD}_{\text{abs}}$ *vs.* $\psi$ (**Extended Data Fig. 7D–F**): the absolute OD reduction is largest at $\psi = 0$ and decays as $\psi$ increases.

To translate these findings into epidemic establishment probabilities, we simulated 5,000 epidemics in a population of size 10,000 for each of the three contact scenarios (Restricted, Unrestricted, Constant), for each of the three benchmark pathogens, and for $\psi \in \{0, 0.25, 0.5, 0.75, 1\}$. We then calculated the proportion of epidemics that successfully established, which we defined as reaching at least 5% cumulative prevalence (500 cases). Additionally, we calculated the analytic epidemic extinction probabilities as a function of $R_0$ and OD using a Negative Binomial branching process approximation[3] . The simulated and analytic extinction probabilities match closely (**Extended Data Fig. 7G–I**), and both show that the gap in establishment probability between the unrestricted and restricted scenarios is greatest when $\psi$ is small.

##### 1.12 How bursty infectiousness impacts growth rate estimates

Burstiness makes estimates of the early-epidemic growth rate $r$ less certain by increasing the amount of variance in daily case counts. Here, we derive the index of dispersion of the daily infection count attributable to a single infector and show that it is monotonically decreasing in the burstiness parameter $\psi$ (*i.e.*, bursty individual GI distributions, with low $\psi$, yield higher index of dispersion).

Consider an index case $i$, infected at time $t_i$, who generates $\chi_i \sim \text{Poisson}(R_0)$ total secondary infections. Under the Gamma burst model, the generation interval for each secondary infection is $\tau = l_i + \epsilon_j$, where $l_i \sim \text{Gamma}((1-\psi)\alpha, \beta)$ is a shared latent period and $\epsilon_j \sim \text{Gamma}(\psi\alpha, \beta)$ is an independent jitter applied to each secondary infection $j$. Let $N_i(\tau)$ be the number of person $i$'s secondary infections that fall on day-since-infection $[\tau, \tau+1)$, and define the probability that a given infection lands on day $\tau$ as

$$Q_i(\tau) = \int_{\tau}^{\tau+1} f_\epsilon(u - l_i)\, du \tag{109}$$

where $f_\epsilon$ is the Gamma$(\psi\alpha, \beta)$ density. The randomness in $Q_i(\tau)$ arises from the draw of $l_i$.

Taking the expectation over $l_i$ gives

$$E[Q_i(\tau)] = \int_{\tau}^{\tau+1} E_{l_i}[f_\epsilon(u - l_i)]\, du = \int_{\tau}^{\tau+1} (f_l * f_\epsilon)(u)\, du = \int_{\tau}^{\tau+1} g(u) du \equiv q(\tau) \tag{110}$$

where $*$ denotes convolution, and the final equality holds since $f_l * f_\epsilon = g$ by construction. Thus, $E[Q_i(\tau)]$ depends only on the population-level generation interval distribution $g$ and is independent of $\psi$.

Using this, we can derive the index of dispersion of $N_i(\tau)$. Conditional on $Q_i(\tau)$, the $\chi_i$ offspring are independently assigned to day $[\tau, \tau + 1)$ with probability $Q_i(\tau)$. By the Poisson thinning property:

$$N_i(\tau)|Q_i(\tau) \sim \text{Poisson}(R_0\, Q_i(\tau)) \tag{111}$$

Applying the law of total variance:

$$\begin{aligned} \text{Var}[N_i(\tau)] &= E[\text{Var}[N_i(\tau) \mid Q_i(\tau)]] + \text{Var}[E[N_i(\tau) \mid Q_i(\tau)]] \\ &= R_0\, E[Q_i(\tau)] + R_0^2\, \text{Var}[Q_i(\tau)] \end{aligned} \tag{112}$$

Since $E[N_i(\tau)] = R_0\, E[Q_i(\tau)] = R_0\, q(\tau)$, the index of dispersion is

$$\text{ID}[N_i(\tau)] = \frac{\text{Var}[N_i(\tau)]}{E[N_i(\tau)]} = 1 + R_0\, \frac{\text{Var}[Q_i(\tau)]}{E[Q_i(\tau)]} \tag{113}$$

In the extreme sustained case ($\psi = 1$), $f_\epsilon = g$ and $l_i = 0$ deterministically, so $Q_i(\tau) = q(\tau)$ is constant across individuals. Thus, $\text{Var}[Q_i(\tau)] = 0$ and $\text{ID}[N_i(\tau)] = 1$. In the extreme spike case ($\psi = 0$), $f_\epsilon$ is a point mass at zero. Then, $Q_i(\tau)$ follows a Bernoulli distribution with parameter $q(\tau)$. Thus, $\text{Var}[Q_i(\tau)] = q(\tau)(1 - q(\tau))$ and

$$\text{ID}[N_i(\tau)] = 1 + R_0(1 - q(\tau)) \tag{114}$$

The index of dispersion therefore varies between these two extremes, and higher $R_0$ inflates the impact of $\psi$ on the daily variation in case counts.

Simulations illustrate the impact of bursty infectiousness on daily case counts and estimates of $r$. We ran infinite-population stochastic simulations (to avoid susceptible depletion effects) for each of the three benchmark pathogens and for $\psi \in \{0, 0.5, 1\}$. For each pathogen-$\psi$ combination, we simulated 5,000 epidemics, each seeded with a single index case at $t = 0$. The simulations were stopped after cumulative infections exceeded a margin sufficient to cover 7 full days of exponential growth past an epidemic establishment threshold of 100 cumulative infections. For each simulated epidemic, we estimated the empirical growth rate $\hat{r}$ by fitting a Poisson generalized linear model with day as the predictor and daily case counts as the response, over a 7-day window beginning the day after cumulative infections first reached 100. The empirical distributions of $\hat{r}$ across simulations, alongside the closed-form Euler-Lotka prediction, are shown in **Extended Data Fig. 8D–F**. The daily infection counts used for the fits are shown in **Extended Data Fig. 8A–C**. In alignment with the theory, bursty infectiousness ($\psi \to 0$) inflates variance in the daily case counts, widening the empirical distribution of $\hat{r}$ around the Euler-Lotka-predicted value. This effect is most pronounced for the pathogens with high $R_0$ (SARS-CoV-2 omicron, measles), as predicted by Eq. 113.

##### 1.13 How bursty infectiousness impacts estimation of the generation interval distribution

Bursty individual GI distributions can make estimates of the population-level $g(\tau)$ less certain. Typically, $g(\tau)$ is estimated using serial intervals obtained by contact tracing, from which generation intervals are inferred by assuming an observation delay model (*e.g.*, an incubation period distribution). Often, these serial intervals are assumed to reflect independent draws from $g(\tau)$ (with additional observation noise). However, when infectiousness is bursty, serial intervals that arise from a single index case are highly correlated, reducing the effective amount of information that each serial interval provides about $g(\tau)$.

This idea can be formalized by estimating the effective sample size obtained by observing $n$ offspring from $k$ index cases, relative to observing $n$ truly independent draws from $g(\tau)$. Kish's design effect for cluster sampling[23] gives an effective sample size of $n_{\text{eff}} = n/(1 + (R_0 - 1)\text{ICC})$, where ICC is the intraclass correlation coefficient. Under the Gamma burst model, and assuming independent Gamma$(a_{\text{obs}}, b_{\text{obs}})$ observation delays applied to each infection time, the ICC is

$$\text{ICC} = \frac{\text{Cov}(s_{ij}, s_{ik})}{\text{Var}(s_{ij})} = \frac{(1-\psi)\alpha/\beta^2 + \sigma_d^2}{\alpha/\beta^2 + 2\sigma_d^2}. \tag{115}$$

where $\text{Cov}(s_{ij}, s_{ik})$ is the covariance between serial interval times from two siblings of the same index case $i$, and $\text{Var}(s_{ij})$ is the variance of a single observed serial interval. These quantities can be derived by noting that

$$s_{ij} = (l_i + \epsilon_j) + (d_j - d_i) = (l_i - d_i) + (\epsilon_j + d_j) \tag{116}$$

where $d \overset{iid}{\sim} \text{Gamma}(a_{\text{obs}}, b_{\text{obs}})$, $l_i \sim \text{Gamma}((1-\psi)\alpha, \beta)$, and $\epsilon_j \sim \text{Gamma}(\psi\alpha, \beta)$. Thus,

$$\text{Cov}(s_{ij}, s_{ik}) = \text{Var}(l_i - d_i) = \text{Var}(l_i) + \text{Var}(d_i) = \frac{(1-\psi)\alpha}{\beta^2} + \sigma_d^2 \tag{117}$$

where $\sigma_d^2 = a_{\text{obs}}/b_{\text{obs}}^2$; and

$$\text{Var}(s_{ij}) = \text{Var}(l_i) + \text{Var}(\epsilon_j) + \text{Var}(d_i) + \text{Var}(d_j) = \frac{\alpha}{\beta^2} + 2\sigma_d^2 \tag{118}$$

The ICC is maximized at $\psi = 0$ and minimized at $\psi = 1$; thus, $n_{\text{eff}}$ decreases as $\psi \to 0$, with the size of the effect modulated by the variance of $g(\tau)$ (*i.e.*, $\alpha/\beta^2$). This reflects the intuition that, when $\psi \to 0$, the offspring of an index case — even if there are many — reflect effectively just one draw from $g(\tau)$.

Simulations demonstrate that, if within-cluster correlations are ignored and serial intervals are instead treated as noisy *iid* draws from $g(\tau)$, bursty individual GI distributions cause posterior estimates of the population-level $\alpha$ and $\beta$ to frequently miss the true value. For each of the three benchmark pathogens and for $\psi \in \{0.1, 0.3, 0.5, 0.7, 0.9, 1.0\}$, we simulated $k = 100$ transmission clusters. Each index case $i$ generated $\chi_i \sim \text{Poisson}(R_0)$ offspring with infection times drawn from the Gamma burst model, and each infection (index case and offspring) was assigned an independent Gamma$(a_{\text{obs}} = 4, b_{\text{obs}} = 1)$ observation delay. The observed serial intervals $s_{ij} = \tau_{ij} + d_j - d_i$ were extracted from each cluster and pooled across clusters within a dataset. We then estimated the joint posterior over $(\alpha, \beta)$ on a $100 \times 100$ grid centered on the true values, under a flat prior and an *iid* likelihood that treats every observed serial interval as an independent draw from the marginal serial-interval density $g * f_{d_j - d_i}$. From each joint posterior we extracted marginal 95% credible intervals for $\alpha$ and $\beta$ separately. We repeated this procedure across 500 simulated datasets per pathogen-$\psi$ combination, and computed empirical coverage as the proportion of replicates in which the marginal 95% credible interval contained the true parameter value. The resulting coverage curves are shown in **Extended Data Fig. 9C**, with representative posterior densities for $\alpha$ and $\beta$ in **Extended Data Fig. 9A–B**. Empirical coverage drops sharply below the nominal 95% as $\psi \to 0$, consistent with the ICC-driven loss of effective sample size derived above.

##### 1.14 Alternatives to the Gamma burst model

Our central findings — that bursty infectiousness inflates coincidence superspreading, makes epidemic establishment times more variable, intensifies the sensitivity of detect-and-isolate intervention to subtle detection-time shifts, and inflates uncertainty in epidemiological parameter estimates — do not depend on the specific parametric form of the burst model. We illustrate this by re-deriving the central results (focusing on SARS-CoV-2) under two alternative burst models.

To further motivate the consideration of alternative burst models, the Gamma burst model has two central limitations: first, it requires $g(\tau)$ to be Gamma-distributed; and since the Gamma distribution decays exponentially, it cannot capture "heavy-tailed" dynamics (*i.e.*, a non-negligible risk of long generation intervals) that are a feature of some pathogens. Second, it requires the latent period distribution $f_l$, the burst distribution $f_\epsilon$, and the generation interval distribution $f_g$, to all share a common rate $\beta$. This is a significant structural constraint: when the Gamma shape parameter is $< 1$, this introduces a singularity at 0; so, while the distribution remains integrable, the burst begins with a brief period of extremely high infectiousness that monotonically decays, which lacks firm biological grounding. The main advantage of the Gamma burst model is its tractability: the latent period distribution, burst distribution, and population-level GI distribution are all Gammas with simple parameters, making it straightforward to simulate secondary infection times and to conduct rigorous mathematical derivations.

For our burst-model alternatives, we require two fundamental properties: (a) by definition, $\psi$ must represent the proportion of $g(\tau)$'s variance that is attributable to within-individual variation in transmission timing; and (b) the infection time $\tau_{ij}$ of secondary case $j$ from index case $i$ should be decomposable as $\tau_{ij} = l_i + \epsilon_j$, where $l_i$ represents a latent period and $\epsilon_j$ follows the burst distribution. Together, these requirements imply that $\text{Var}[l_i] = (1-\psi)\text{Var}[g]$ and $\text{Var}[\epsilon_j] = \psi\text{Var}[g]$. (Note that requirement (a) is necessary by definition, but requirement (b) could be relaxed to obtain a wider family of burst models; we keep the requirement here because the existence of a variable individual-level latent period is biologically motivated[24] and helps provide a more direct comparison to the Gamma burst model.)

**The Log-normal burst model.** The first alternative we will consider is a Log-normal burst model (**Extended Data Fig. 1E–G**), where the generation interval distribution $f_g$ and the burst distribution $f_\epsilon$ are both log-normally distributed. The log-normal distribution is a canonical example of a "heavy-tailed" distribution, allowing for an appreciable chance of long generation intervals.

Let the population-level GI distribution be $f_g \sim \text{Lognormal}(\mu, \sigma^2)$. Next, let the burst distribution be a scaled version of the population-level GI distribution: $f_\epsilon \sim \text{Lognormal}(\mu_\epsilon, \sigma^2)$. We require

$$\psi = \frac{\text{Var}[\epsilon]}{\text{Var}[g]} = \frac{[e^{\sigma^2}-1]e^{2\mu_\epsilon+\sigma^2}}{[e^{\sigma^2}-1]e^{2\mu+\sigma^2}} = e^{2(\mu_\epsilon-\mu)} \tag{119}$$

Thus, $\mu_\epsilon = \log(\sqrt{\psi}) + \mu$.

The mean and variance of $\tau$ and $\epsilon$ are

$$E[\tau] = e^{\mu+\sigma^2/2} \qquad \text{Var}[\tau] = \left(e^{\sigma^2}-1\right)E[\tau]^2 \tag{120}$$

$$E[\epsilon] = \sqrt{\psi}E[\tau] \qquad \text{Var}[\epsilon] = \psi\text{Var}[\tau] \tag{121}$$

Since $f_l$ has no closed form, we approximate it as log-normal with moment-matched parameters. Under the Fenton-Wilkinson approximation[25] , the sum of two log-normals is approximately log-normal, so $\tau = l + \epsilon$ is approximately distributed as $f_g$. Specifically, since $E[l] = E[\tau] - E[\epsilon]$ and $\text{Var}[l] = \text{Var}[\tau] - \text{Var}[\epsilon]$,

$$E[l] = (1-\sqrt{\psi})E[\tau] \qquad \text{Var}[l] = (1-\psi)\text{Var}[\tau] \tag{122}$$

Thus, for the parameters of the moment-matched $f_l \sim \text{Lognormal}(\mu_l, \sigma_l^2)$ distribution, we have

$$\sigma_l^2 = \log\left(1 + \frac{\text{Var}[l]}{E[l]^2}\right) \qquad \mu_l = \log E[l] - \frac{\sigma_l^2}{2} \tag{123}$$

To implement the model, then, we let $l_i \sim \text{Lognormal}(\mu_l, \sigma_l^2)$ (with the parameters given by Eq. 123) and $\epsilon_j \sim \text{Lognormal}(\log(\sqrt{\psi}) + \mu, \sigma^2)$. This gives $\tau_{ij} = l_i + \epsilon_j$ as the secondary infection time, relative to the index case's infection. Now, $\tau$ has the correct mean and variance by construction, but is not exactly log-normal – *i.e.*, it is not an exact draw from $f_g \sim \text{Lognormal}(\mu, \sigma^2)$. However, for a range of plausible parameters, including for the three reference pathogens across the full range of $\psi$, $\tau$ is approximately log-normal-distributed (**Extended Data Fig. 1H–J**).

The log-normal burst model recovers our central findings: bursty individual GI distributions produce coincidence superspreading (**Extended Data Fig. 4D–I**), inflate the variance of the epidemic latent period (**Extended Data Fig. 5G**), create sharp time-dependence in detect and isolate interventions (**Extended Data Fig. 6J–O**), and inflate the variance of daily case counts, making estimates of the epidemic growth rate less certain (**Extended Data Fig. 8G–I**).

**The type-II Gamma burst model** The second alternative burst model begins, like the Gamma burst model, with a Gamma-distributed population-level GI distribution. Now, though, rather than keeping the rate fixed, we match the shape of the burst with the shape of $g(\tau)$. Specifically, if $f_g \sim \text{Gamma}(\alpha, \beta)$, then $f_\epsilon \sim \text{Gamma}(\alpha, \beta/\sqrt{\psi})$. This ensures that $\text{Var}[\epsilon] = \psi \text{Var}[\tau]$. Like the log-normal burst model, this type-II Gamma burst model expresses the shape of the burst as a scaled-down version of the population-level GI distribution (**Extended Data Fig. 1B–D**).

The Gamma distribution is not closed under convolution when the rates are unequal, so $f_l$ is not Gamma-distributed, and we must find another way of characterizing the distribution of $l$. Conveniently, there is a straightforward way to generate exact samples from $f_l$ so that $\tau_{ij} = l_i + \epsilon_j$ follows the population-level $\text{Gamma}(\alpha, \beta)$ distribution exactly. Our strategy is to represent the latent period as a sum of a random (Poisson) number of Exponential jumps. The number of jumps and their sizes are chosen so that their sum, when added to $\epsilon_j \sim \text{Gamma}(\alpha, \beta/\sqrt{\psi})$, yields exactly $\text{Gamma}(\alpha, \beta)$.

For $\tau \sim \text{Gamma}(\alpha, \beta)$ and $\epsilon \sim \text{Gamma}(\alpha, \beta/\sqrt{\psi})$, we have Lévy measures

$$\nu_\tau(dx) = \frac{\alpha e^{-\beta x}}{x} dx \qquad \text{and} \qquad \nu_\epsilon(dx) = \frac{\alpha e^{-\beta x/\sqrt{\psi}}}{x} dx \tag{124}$$

Since $\tau \overset{d}{=} l + \epsilon$ with $l \perp \epsilon$, the Lévy measures subtract:

$$\nu_l(dx) = \nu_\tau(dx) - \nu_\epsilon(dx) = \frac{\alpha(e^{-\beta x} - e^{-\beta x/\sqrt{\psi}})}{x} dx \tag{125}$$

The total mass of $\nu_l$ is finite:

$$\Lambda = \int_0^\infty \nu_l(dx) = \alpha \int_0^\infty \frac{e^{-\beta x} - e^{-\beta x/\sqrt{\psi}}}{x} dx = -\frac{\alpha}{2} \log(\psi) \tag{126}$$

Thus, the distribution with Lévy measure $\nu_l(dx)$ is a compound Poisson distribution; *i.e.*, we can express $l$ as a sum of

$N \sim \text{Poisson}(\Lambda)$ jumps, where the jump sizes are distributed according to $\nu_l(dx)/(\Lambda dx)$. This jump size density is

$$f_{\text{jump}}(x) = \frac{\nu_l(dx)/dx}{\Lambda} = \frac{\alpha}{\Lambda} \cdot \frac{e^{-\beta x} - e^{-\beta x/\sqrt{\psi}}}{x} \tag{127}$$

$$= \frac{\alpha}{\Lambda} \int_{\beta}^{\beta/\sqrt{\psi}} e^{-ux} du \tag{128}$$

$$= \int_{\beta}^{\beta/\sqrt{\psi}} ue^{-ux} \cdot \frac{\alpha}{\Lambda u} du \tag{129}$$

$$= \int_{\beta}^{\beta/\sqrt{\psi}} ue^{-ux} \cdot \frac{\alpha}{-\frac{\alpha}{2}\log(\psi)u} du \tag{130}$$

$$= \int_{\beta}^{\beta/\sqrt{\psi}} ue^{-ux} \cdot \frac{1}{\log((\beta/\sqrt{\psi})/\beta)u} du \tag{131}$$

$$= \int_{\beta}^{\beta/\sqrt{\psi}} ue^{-ux} \cdot \frac{1}{u[\log((\beta/\sqrt{\psi})) - \log(\beta)]} du \tag{132}$$

This last integral represents a compound probability distribution $\text{Exp}(u)$, where $u$ follows a log-uniform distribution, *i.e.* $\log(u) \sim \text{Unif}(\log(\beta), \log(\beta/\sqrt{\psi}))$. This gives us a simple algorithm for generating draws from $f_l$:

- Draw $N \sim \text{Poisson}(\Lambda)$ jumps
- For each jump $j \in \{1, \ldots, N\}$, draw $\log(u_j) \sim \text{Unif}(\log(\beta), \log(\beta/\sqrt{\psi}))$
- For each jump, draw $X_j \sim \text{Exp}(u)$
- Assemble the draw: $l = \sum_{j=1}^{N} X_j$

When $N = 0$, $l = 0$, which happens with probability $e^{-\Lambda}$; thus, some fraction of infections have no latent period and begin with an immediate burst.

Simulations confirm that generating $l_i$ in this manner, and adding $\epsilon_j \sim \text{Gamma}(\alpha, \beta/\sqrt{\psi})$, yields the target $\tau_{ij} \sim \text{Gamma}(\alpha, \beta)$ (**Extended Data Fig. 1H–J**). As with the log-normal burst model, this type-II Gamma burst model recovers our central findings: bursty individual GI distributions produce coincidence superspreading (**Extended Data Fig. 4D–I**), inflate the variance of the epidemic latent period (**Extended Data Fig. 5G**), create sharp time-dependence in detect and isolate interventions (**Extended Data Fig. 6J–O**), and inflate the variance of daily case counts, making estimates of the epidemic growth rate less certain (**Extended Data Fig. 8G–I**).
